# Risk factors for atopic and non-atopic asthma in school-aged children from high-, and low-and-middle-income countries

**DOI:** 10.1101/2024.06.03.24308369

**Authors:** Charlotte E Rutter, Harriet Mpairwe, Camila A Figueiredo, Mary Njoroge, Steven Robertson, Hajar Ali, Collin Brooks, Jeroen Douwes, Philip J Cooper, Martha Chico, Natalia Romero-Sandoval, Alvaro A Cruz, Pius Tumwesige, Mauricio L Barreto, Neil Pearce, Lucy Pembrey, the CAMERA Study Group

## Abstract

**Background:** It is well established that there are different asthma phenotypes, but whereas determinants of atopic asthma are well studied, little is known about non-atopic asthma. We compared risk factors for atopy, atopic asthma (AA) in atopics, and non-atopic asthma (NAA) in non-atopics, in children in a wide variety of countries.

**Methods:** Using four studies, across 23 countries, we assessed asthma status and atopy (skin prick tests) for children aged 6-17, plus risk factors from housing, heating, pets, family, diet, and air-quality categories. Using mixed effects logistic regression models we assessed risk factors over 4 pathways: Pathway 1: non-atopic non-asthma to NAA; Pathway 2: non-atopic non-asthma to atopy (no asthma); Pathway 3: atopic non-asthma to AA; Pathway 4: non-atopic non-asthma to AA. We compared the log odds of risk factors between pathways using Pearson’s correlation coefficient.

**Results:** Our final sample of 32741 children comprised 67% with neither atopy nor asthma, 15% with atopy but without asthma, 8% with AA and 10% with NAA. Risk factors were similar between Pathway 1 and Pathway 3 (Pearson’s correlation = 0.81, 95% confidence interval = [0.68, 0.94]). In contrast, risk factors differed between Pathway 2 and Pathway 3 (−0.06, [−0.29, 0.17]).

**Discussion:** These findings indicate that although atopy increases the risk of asthma, the risk factors for subsequently developing asthma are generally the same in those with and without atopy. This raises important questions about the role of atopy in asthma, particularly whether it is an inherent part of the aetiological process or is coincidental.

**Key messages:** *What is already known on this topic:* It is well established that there are different phenotypes of asthma but little is known about risk factors for non-atopic asthma.

*What this study adds:* Using a novel approach, we found that lifestyle and environmental risk factors for developing asthma are generally similar in atopic children and non-atopic children but the risk factors for atopy are quite different.

*How this study might affect research, practice or policy:* Our findings suggest that atopy and asthma may be coincidental in a large proportion of children who are defined as having atopic asthma. This has important implications for our understanding of the causes and mechanisms of different asthma phenotypes, and therefore prevention and treatment of asthma.

## Introduction

Although the importance of intrinsic (or non-allergic or non-atopic) asthma has been recognised since the early 20th century,[1] the focus of asthma research in the past 60 years has been predominantly on allergic mechanisms despite compelling evidence that at most half of asthma cases are attributable to atopy.[2,3]

As a consequence, little is known about the causes and pathophysiological mechanisms of non-atopic asthma (NAA), hampering the development of effective prevention and treatment options for a large proportion of people with asthma.

Atopic asthma (AA) is associated with T helper type 2-mediated airways inflammation (Th2), with atopy often identified (with varying results) by skin prick test (SPT) positivity to common allergens or serum allergen-specific IgE measurement (commonly associated with airways eosinophilia).[4] The mechanisms underlying NAA remain unclear but may involve neutrophilic inflammation, neural mechanisms, infection-related responses, or other currently unknown pathophysiological pathways.[5–7] Different mechanisms may lead to different risk factors associated with disease. To make a valid comparison between NAA and AA we first need to tease apart the risk factors for asthma from the risk factors for atopy.

Figure 1 shows a framework for pathways potentially involved in the development of AA and NAA.[8] These are not often considered separately in studies of risk factors, instead most have done one of the following:[8] (i) compared all asthma cases with all non-asthmatic controls (a mix of all four pathways); (ii) adjusted for atopy (an average of the effects of pathway 1 and pathway 3); (iii) compared AA with all non-asthmatic controls (a mix of pathways 2-4); (iv) compared NAA with all non-asthmatic controls (none of the indicated pathways); or a few studies have (v) stratified on atopy (yielding separate estimates for pathways 1 and 3).

**Figure 1:**
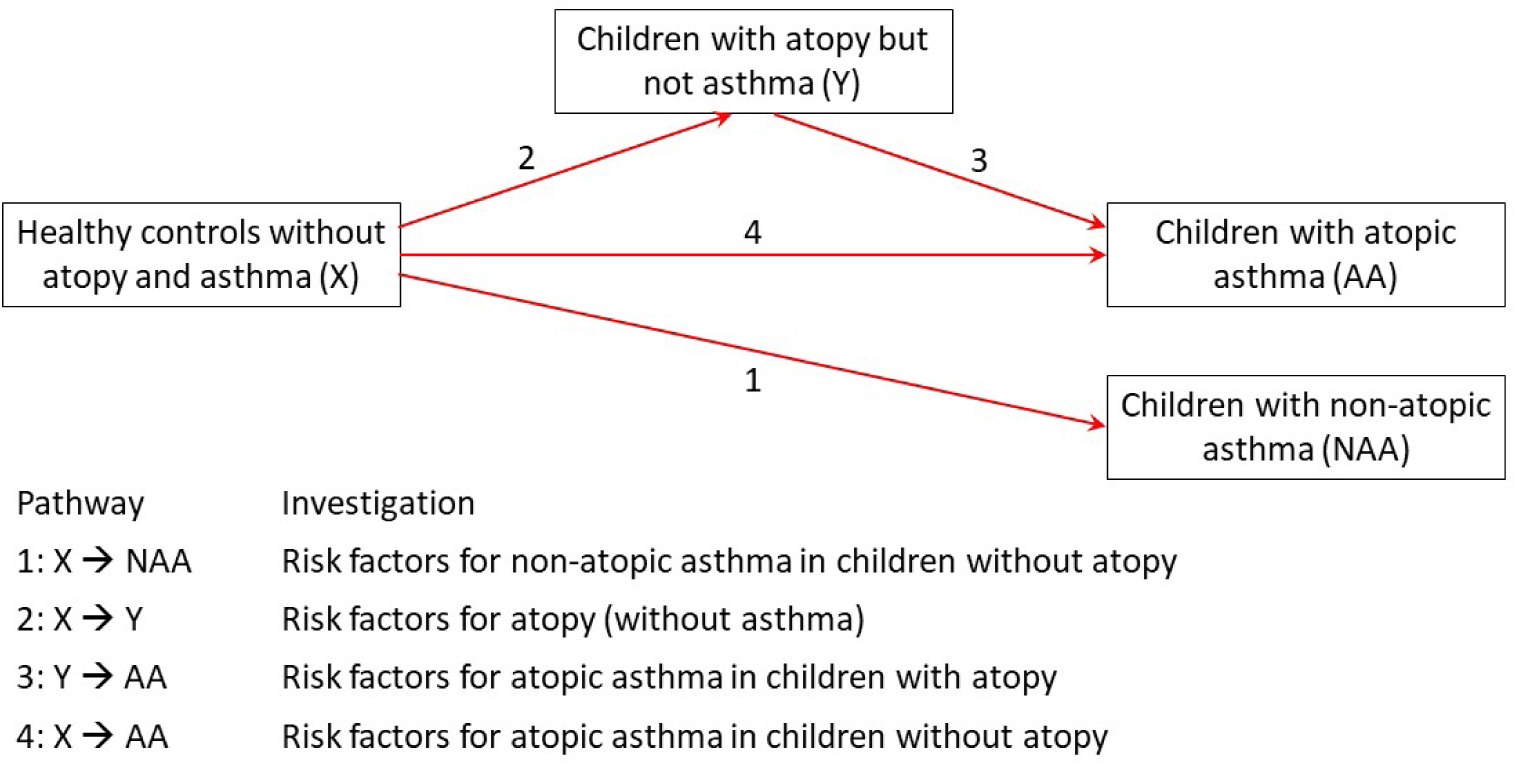
Pathways of possible asthma causation (Adapted from Barreto et al, 2010)

A 2014 review of the evidence for NAA risk factors found that, of 30 risk factors evaluated in 43 studies, only three (family history of allergic disease, household dampness/mould, and childhood lower respiratory tract infections) showed consistent associations with NAA. A lack of breastfeeding, or breastfeeding for a limited period, was less consistently associated with NAA. However, this review noted that most of these studies used inappropriate comparison groups.[9] There was also considerable methodological heterogeneity with different definitions of atopy and asthma, and different study designs, hampering valid comparisons.[9]

Here, using data from four studies: the International Study of Asthma and Allergies in Childhood (ISAAC) Phase II, the World Asthma Phenotypes Study (WASP), the Social Change, Asthma and Allergy in Latin America Study (SCAALA), and the Avon Longitudinal Study of Parents and Children (ALSPAC), we aimed to separately assess and compare risk factors for the four pathways (Figure 1), allowing the identification of modifiable factors that may increase/decrease the risk of NAA (pathway 1) and/or AA (pathways 2-4).

## Methods

### Data sources

The ISAAC Phase II cross-sectional questionnaire survey (with additional objective tests) commenced in 1998, targeting children aged 10-11, and included 30 centres from a mix of high-income and low-middle-income countries. Detailed methods have been published previously.[10]

The WASP cross-sectional case-control study was conducted in five centres in high-income (UK, NZ) and low-middle-income countries (Brazil, Ecuador, Uganda) between 2017 and 2020. Participants in Brazil, Ecuador and New Zealand were recruited from ongoing population-based cohort studies, with additional recruitment through the community (e.g., surveys in schools). In Uganda, participants were recruited from a larger case-control study of children with and without asthma identified through a cross-sectional survey in schools. Full details of the questionnaire and methods have been published previously.[4] The UK centre surveyed young adults (26-27 years) and has therefore been excluded from this analysis.

The SCAALA study involved a cross-sectional survey in Ecuador and a longitudinal study in Brazil, with detailed protocols published previously. In Ecuador, data were collected in 2006 on children aged 5-16 years in the Esmeraldas province.[11] In Brazil, data were collected in 2005 on children aged 4-11 years at baseline in the city of Salvador, Northeastern Brazil.[12]

For the ALSPAC birth cohort study, pregnant women resident in Avon, UK with expected dates of delivery between 1st April 1991 and 31st December 1992 were invited to take part in the study. The initial number of pregnancies enrolled was 14,541 from which 13,988 children were still alive at 1 year of age, consisting of 14,203 unique women (G0 mothers). Information was collected from 12,113 G0 partners (of the mothers) of which 3,807 are currently enrolled on the study. After 7 years, efforts were made to find eligible pregnancies that were missing from the study. The total sample size for analyses using any data collected after the age of seven is therefore 15,447 pregnancies, from which 14,901 children were still alive at 1 year of age. The study included regular surveys and tests throughout childhood, details of which have been reported previously.[13,14] Here we use SPT data at age 7 and surveys collected up to age 7. Please note that the study website contains details of all the data that is available through a fully searchable data dictionary and variable search tool.[15]

From all sources, individuals with missing data on symptoms, skin prick test results, age or sex were excluded, along with those outside the target age group of 6-17 years, and those who did not fit into a category of case or control (i.e. with previous asthma symptoms/diagnosis but not in the past year). Missingness was checked for associations with demographics.

### Variable definitions

Children were defined as having current asthma if they had wheeze symptoms and/or used asthma medications in the past 12 months. They were defined as without asthma if they never had asthma or asthma symptoms, determined by negative responses to questions on ever having wheeze symptoms or diagnosed asthma (ISAAC and SCAALA) or by negative responses to questions on recent wheeze symptoms across all surveys in ALSPAC. Participants qualifying for neither status (e.g., those who previously had asthma) were excluded from the analyses. More details on the exact definitions for each study are in supplementary Table S1.

We defined atopy as SPT positivity (≥3mm wheal in ISAAC, WASP and SCAALA, ≥2mm wheal in ALSPAC) to at least one of a panel of five or more common allergens. Individuals testing positive for saline (negative control), negative for histamine (positive control), or with fewer than five allergens tested, were excluded. Similar SPT methods were used in each study, and allergens appropriate for the local environment of each centre were used (supplementary Table S2).

Study participants were categorised into four outcome groups: healthy controls (no atopy and no asthma) (X), those with atopy but no asthma (Y), those with atopic asthma (AA) and those with non-atopic asthma (NAA).

We included potential risk factors if they were available in ISAAC as it was by far the largest study. These were all binary variables and included housing condition (heating, cooling, insulation, cooking fuel, feather/synthetic bedding, damp), animal contact (pet cats, dogs, birds and farm animals), diet (meat, fish, fruit, vegetables, fast food), body mass index (BMI) category, family (siblings, parental allergies/education/smoking) and early life factors (breastfeeding, birthweight, prematurity). (Supplementary Table S1)

### Statistical analyses

Four sets of models were used, representing each pathway in Figure 1. Pathway 1: comparing those with NAA to healthy controls (risk factors for NAA) Pathway 2: comparing those with atopy but without asthma to healthy controls (risk factors for atopy) Pathway 3: comparing those with AA to those with atopy but not asthma (risk factors for AA) Pathway 4: comparing those with AA to healthy controls (risk factors for AA and atopy together)

For the first three pathways we examined which risk factors were associated with NAA (Figure 1, pathway 1), atopy (pathway 2), and AA in atopic children (pathway 3). Pathway 4 was more difficult to address as it is in addition to the two existing pathways, 2 and 3. It was assessed by estimating the overall association of non-atopic non-asthma (X) with AA (not adjusted or stratified on atopy) and comparing it with that predicted assuming no direct path, i.e., the Odds Ratio (OR) for pathway 2 multiplied by the OR for pathway 3. A difference between the two figures was considered to indicate a possible direct path (assuming the difference was not due to misclassification or residual confounding). In addition, we directly compared AA to NAA to see which factors differentiated them and how these compared to the risk factors for atopy alone.

Mixed-effects logistic regression models were fitted on the binary outcome representing each pathway, with a random intercept for centre and adjusting for age, sex, study, and country income level (LMIC/HIC). In children, asthma prevalence is known to increase with age, peaking around 13-14 years, after which it decreases.[16] Thus, using the models with no risk factors we first tested for evidence against a linear trend for age by adding a quadratic term and then by adding a restricted cubic spline (RCS); the most appropriate form was then used for the models of each pathway.

Individual models were fitted for each risk factor in turn, and results compared between pathways. The sample size varies between models as the amount of missing data for each risk factor varies. In addition, some risk factors were not included for a particular study; as stated previously, all included risk factors were in the ISAAC study and at least one other study (Supplementary Table S1). To assess the relationship between risk factors for each pathway we plotted the OR estimates for one pathway against another (on the log scale as the log odds is usually normally distributed) and calculated Pearson’s correlation coefficient for the log odds and associated bootstrapped confidence intervals (using bootstrap samples of the individual level dataset to account for the uncertainty of the individual estimates as well as the uncertainty of the correlation) for 1,000 replications.

Models stratified by country income group were fitted, to test for interactions, and separate models for each study, to assess consistency.

Sensitivity analyses were conducted on the main non-stratified models as follows:

1. To exclude borderline atopics, two stricter atopy definitions were applied to data from ISAAC and ALSPAC (due to data availability): any wheal of at least 5mm in the core allergens; 2 or more wheals of at least 4mm in the core allergens. Those originally classed as atopic but not meeting the new definition were excluded from that analysis. The definition for non-atopics remained the same. Results were compared to those using the standard atopy definition but limited to the two studies;
2. To check if there was an imbalance between studies, four analyses were conducted leaving out one study each time;
3. Lastly, an analysis was conducted that used a more lenient definition for the controls, allowing for a small amount of missing symptom data (either one past questionnaire missing for ALSPAC or the question “Has the child ever been told they have asthma?” missing in ISAAC).

Data were analysed using Stata version 18.[17]

## Results

From a total sample of 71916 participants (1205 from WASP, 54271 from ISAAC, 8266 from SCAALA and 8174 from ALSPAC), 33149 were deemed ineligible for either being out of the age range, not being offered an SPT, or previously having asthma symptoms but not in the past year. This left a sample of 38,767 participants to be considered for analysis. Of these, 6026 (16%) were excluded due to missing data: demographics (21); SPT data (1705); and symptom data (4300) (Table 1). There were some relatively weak associations between missing data and demographics; being male was associated with missing SPT data (OR=1.15, 95% Confidence interval (CI) = [1.03, 1.29]), but not with missing symptom information. Older children were associated with a higher chance of missing SPT data (1.07, [1.02, 1.13]) but a much lower chance of missing symptom information (0.63, [0.60, 0.65]). Individuals with current asthma were more likely to be missing the SPT data than those who never had asthma (1.27, [1.09, 1.47]). (Supplementary Table S3)

**Table 1:**
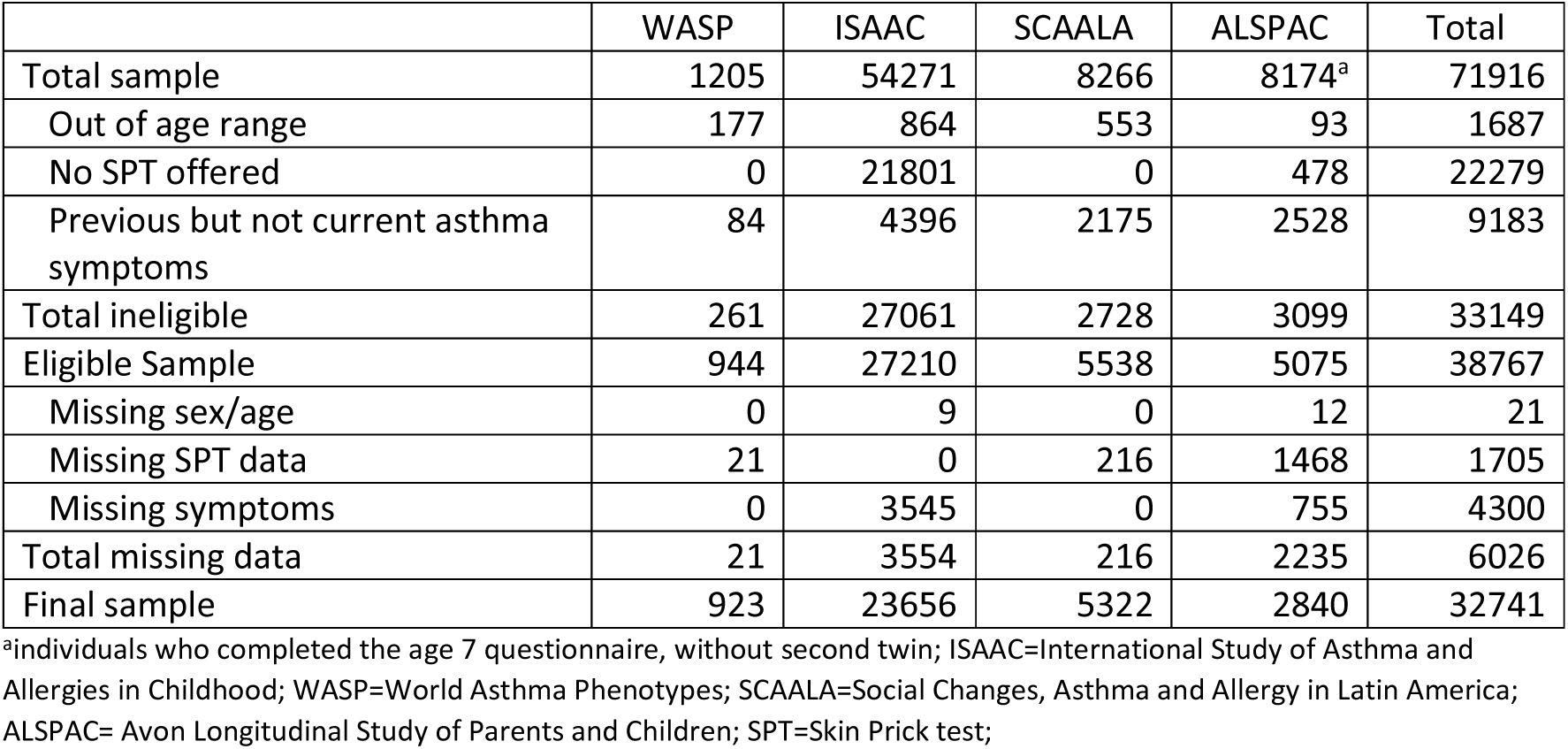
Exclusions.

|  | WASP | ISAAC | SCAALA | ALSPAC | Total |
| --- | --- | --- | --- | --- | --- |
| Total sample | 1205 | 54271 | 8266 | 8174<sup>a</sup> | 71916 |
| Out of age range | 177 | 864 | 553 | 93 | 1687 |
| No SPT offered | 0 | 21801 | 0 | 478 | 22279 |
| Previous but not current asthma symptoms | 84 | 4396 | 2175 | 2528 | 9183 |
| Total ineligible | 261 | 27061 | 2728 | 3099 | 33149 |
| Eligible Sample | 944 | 27210 | 5538 | 5075 | 38767 |
| Missing sex/age | 0 | 9 | 0 | 12 | 21 |
| Missing SPT data | 21 | 0 | 216 | 1468 | 1705 |
| Missing symptoms | 0 | 3545 | 0 | 755 | 4300 |
| Total missing data | 21 | 3554 | 216 | 2235 | 6026 |
| Final sample | 923 | 23656 | 5322 | 2840 | 32741 |

<sup>a</sup>individuals who completed the age 7 questionnaire, without second twin; ISAAC=International Study of Asthma and Allergies in Childhood; WASP=World Asthma Phenotypes; SCAALA=Social Changes, Asthma and Allergy in Latin America; ALSPAC= Avon Longitudinal Study of Parents and Children; SPT=Skin Prick test;

The final sample of 32741 comprised 66% healthy controls, 15% children with atopy but not asthma, 8% with AA and 10% NAA. The WASP sample had a higher proportion of children with asthma (75%) than the others (15-30%) due to its case-control design (Table 2).

**Table 2:** Distribution of main outcome by study.

| Study | Healthy controls, no atopy, no asthma (X) (%) | Atopy but no asthma (Y) (%) | Atopic Asthma (AA) (%) | Non-Atopic Asthma (NAA) (%) | Total |
| --- | --- | --- | --- | --- | --- |
| **WASP** | 171 (19%) | 66 (7%) | 422 (46%) | 264 (29%) | 923 |
| **ISAAC** | 16239 (69%) | 3922 (17%) | 1584 (7%) | 1911 (8%) | 23656 |
| **SCAALA** | 3860 (73%) | 560 (11%) | 203 (4%) | 699 (13%) | 5322 |
| **ALSPAC** | 1650 (58%) | 322 (11%) | 433 (15%) | 435 (15%) | 2840 |
| **TOTAL** | 21920 (67%) | 4870 (15%) | 2642 (8%) | 3309 (10%) | 32741 |

ISAAC=International Study of Asthma and Allergies in Childhood; WASP=World Asthma Phenotypes; SCAALA=Social Changes, Asthma and Allergy in Latin America; ALSPAC= Avon Longitudinal Study of Parents and Children

Participants in WASP were on average the oldest but with a wide range (9-18 years). Those in ALSPAC were 7-8 years, ISAAC were mainly 10-11 years and SCAALA ranged from 6-16 years (Supplementary Figures S1-S3). The WASP dataset was 55% female, SCAALA was 48% and ISAAC and ALSPAC were both 51%. In WASP 64% of the individuals were from LMIC and in ISAAC this was 32%. SCAALA participants were all from LMIC (Brazil and Ecuador) and ALSPAC were all from HIC (UK).

Details of the final sample by country are in Supplementary Table S4.

The prevalence of each risk factor by study is presented in Table 3 (expanded to include individual centres in Supplementary Table S5). Each risk factor was available for a different subset of individuals, ranging from 14060 for synthetic pillows to 31175 for parental allergic diseases. There was considerable variation between centres for some variables, particularly indoor heating questions as some centres in hot countries do not use heating at all.

**Table 3:**
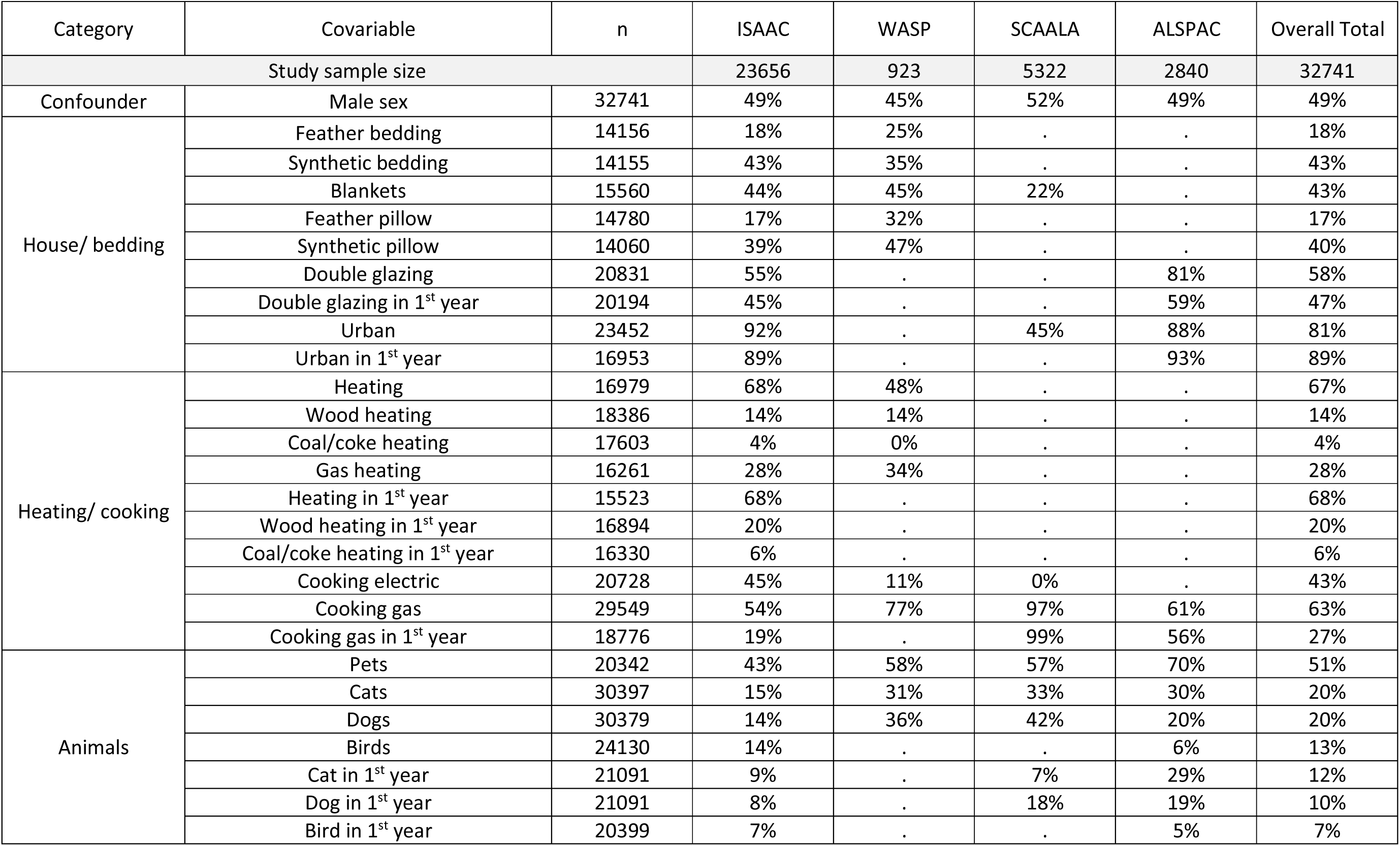
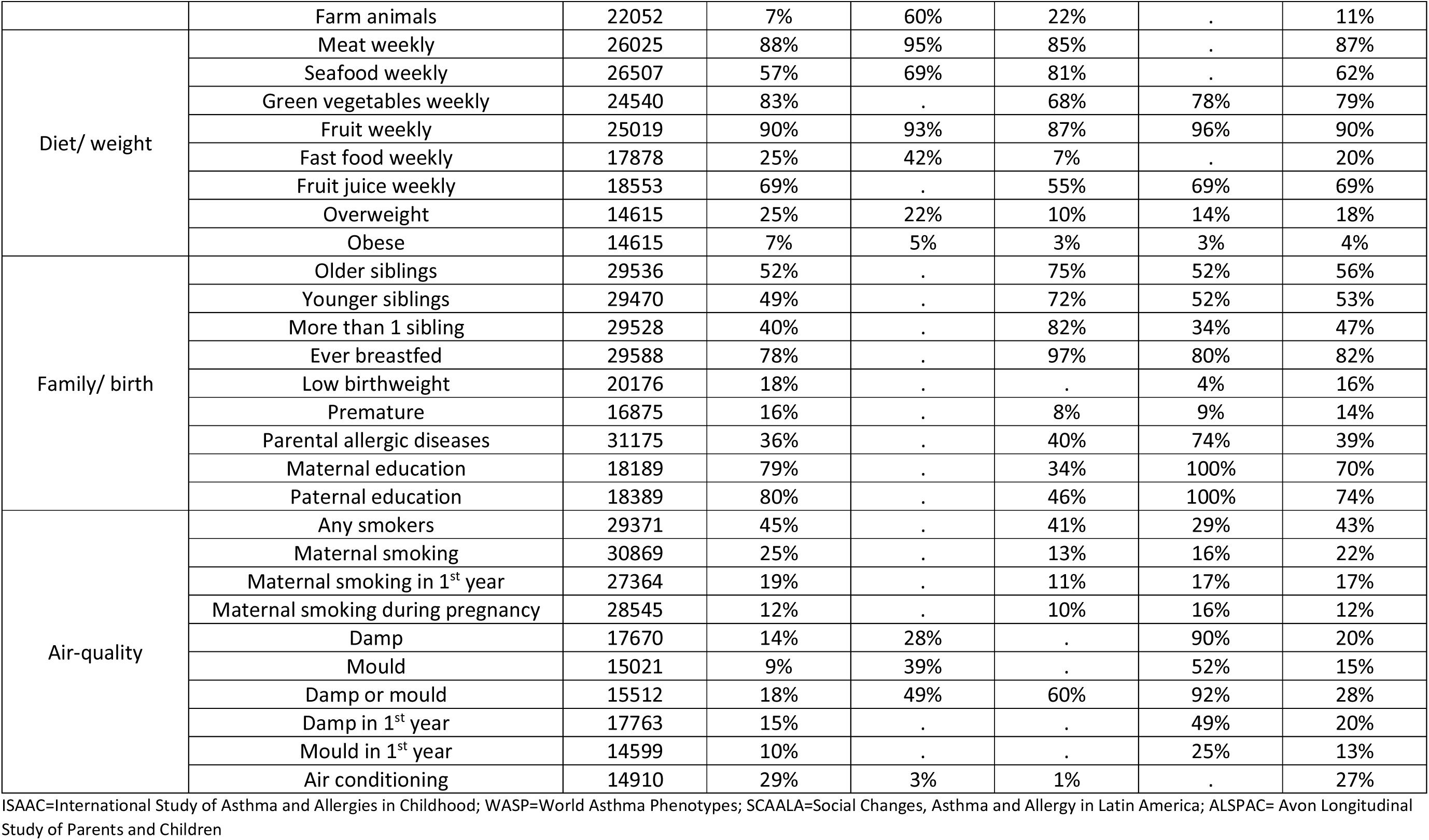
Prevalence of potential risk factors by study.

Results from the mixed effects logistic regression, with random intercepts for centre and adjusted for study, sex, age and country income level (LMIC/HIC), but without including risk factors, showed evidence that the effect of age was not linear for pathway 1 (quadratic *P*=0.046, RCS *P*=0.17) and for the AA/NAA comparison (quadratic *P*=0.016, RCS *P*=0.23), but there was no evidence against linearity for pathways 2-4 (quadratic *P*=0.27, *P*=0.18, *P*=0.45 and RCS *P*=0.43, *P*=0.87, *P*=0.87 respectively). Thus, a quadratic age term was included in all models for pathway 1 and the AA/NAA comparison.

Figure 2 compares the estimated ORs of each risk factor between AA in already atopic individuals (pathway 3), and the risk of NAA in non-atopic individuals (pathway 1). The associations for the included risk factors were generally similar for AA and NAA, with a high positive correlation between the estimated log odds (Pearson correlation coefficient=0.81; bootstrapped 95% CI=[0.68, 0.94]).

**Figure 2:**
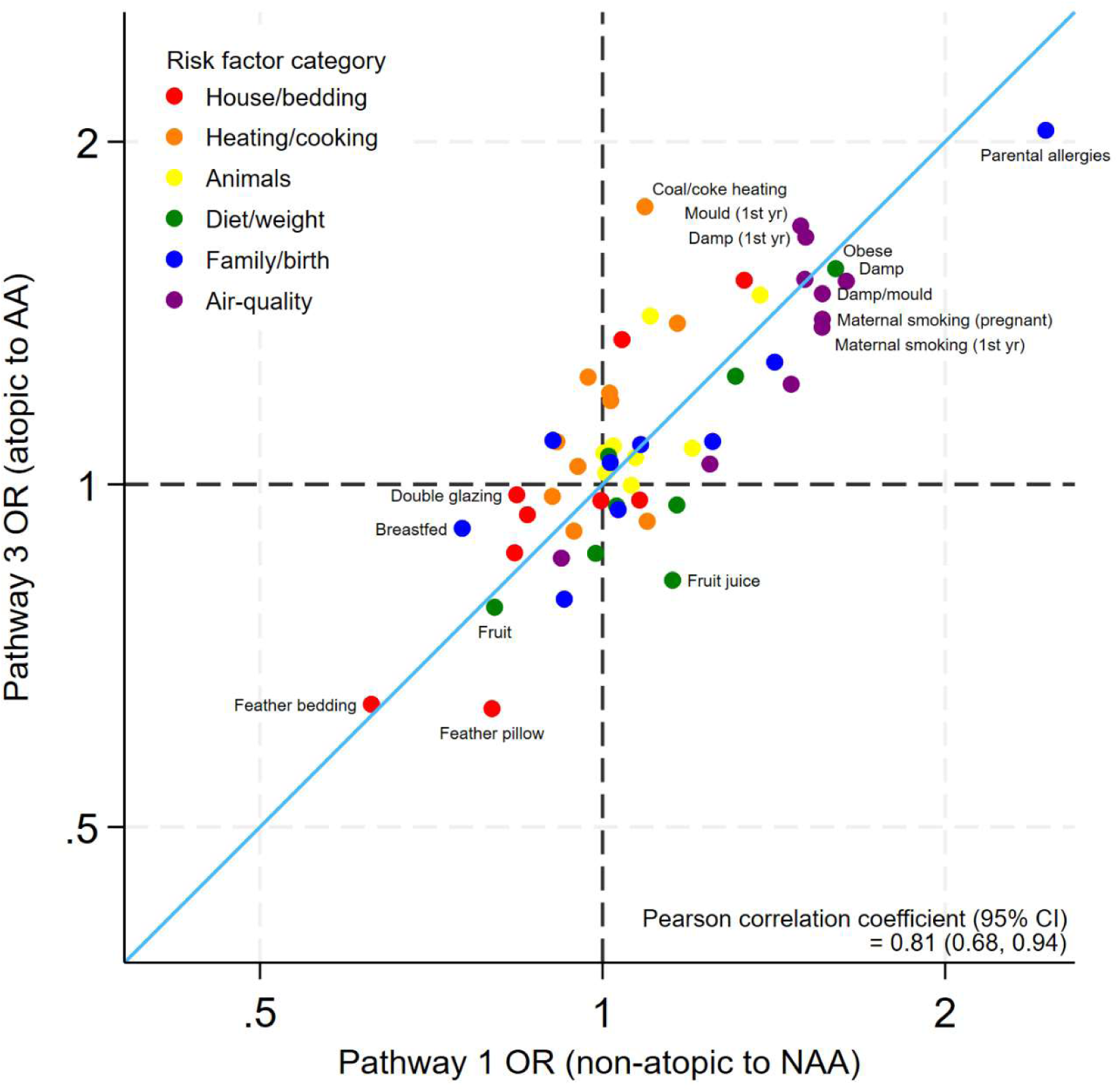
**Estimated Odds ratios (ORs) of risk factors for atopic asthma compared to non-atopic asthma, excluding atopy risk (Pathways 3 v 1)** Axes are shown on the natural log scale.

Figure 3 compares risk factors for atopy (pathway 2) with risk factors for AA in atopic individuals (pathway 3) with no correlation between the two groups (−0.06; [−0.28, 0.16]).

**Figure 3:**
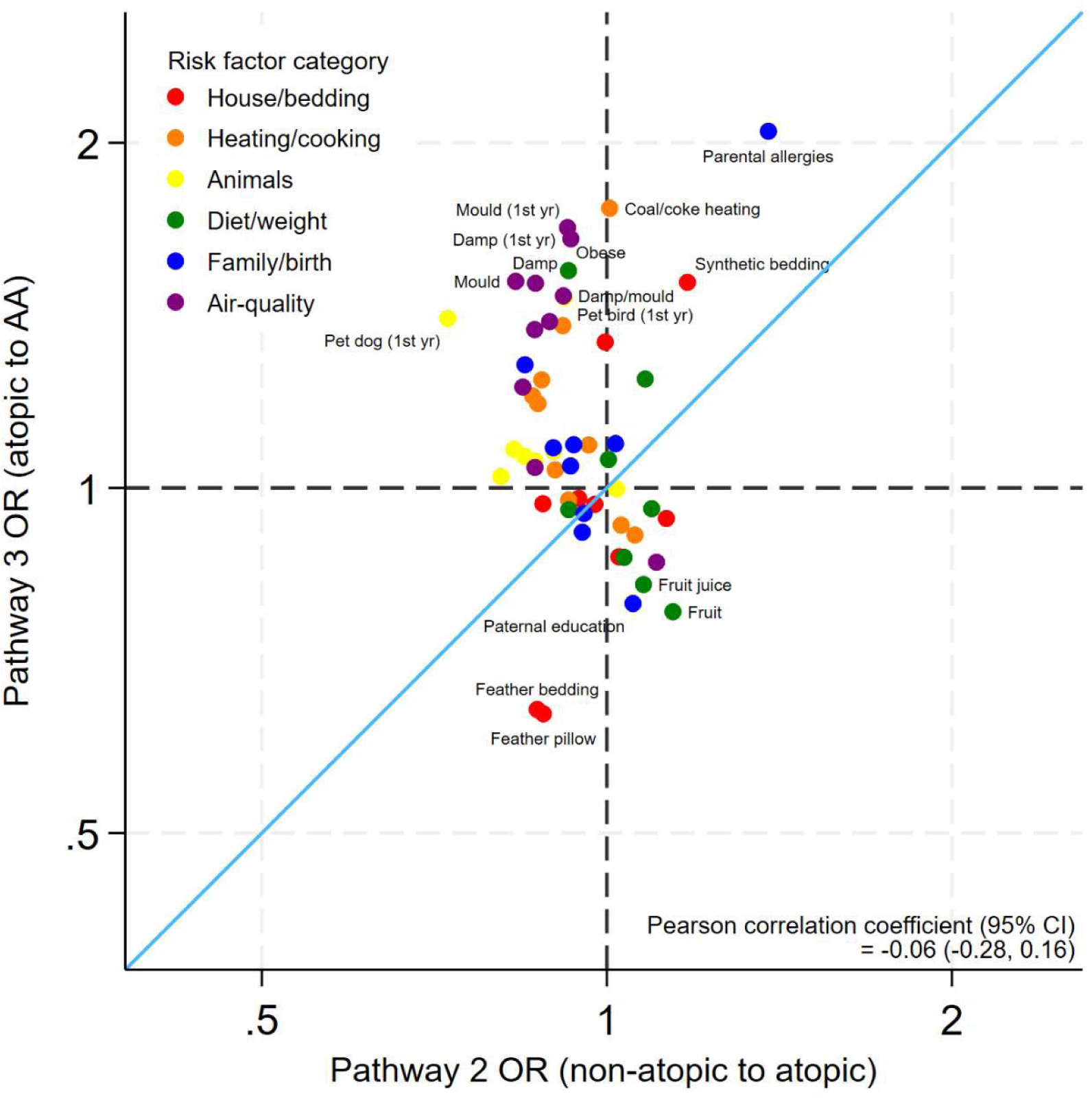
**Estimated Odds ratios (ORs) of risk factors for asthma (in atopic individuals) compared to atopy (Pathways 3 v 2)** Axes are shown on the natural log scale.

We also compared the risk factors for atopy (pathway 2) with the risk factors for AA compared to NAA, as in both of these groups the difference is atopy. We found a moderate positive correlation (0.54; [0.34, 0.74]) between the risk factor log odds of these groups (Supplementary Figure S4).

The individual results for each risk factor and outcome are shown in Supplementary Table S6, along with predictions of pathway 4, based on the results for pathway 2 and 3. The observed and the predicted values are similar so there is likely no direct pathway 4 (non-atopic to AA) over and above the two separate pathways. Results comparing pathways 1-3 by category of risk factor are shown in Supplementary Figures S5-S10.

The strongest risk factor for asthma was parental history of allergic disease (asthma, eczema or hay fever) (NAA OR=2.45 [95% CI=2.24-2.69]; AA 2.05 [1.81-2.31]). Breastfeeding (ever) was associated with a lower risk of NAA (0.75 [0.67-0.85]). Premature birth (1.42 [1.19-1.69]) and low birthweight (1.25 [1.05-1.49]) were associated with an increased NAA risk. (Supplementary Figure S9)

Bedding-related risk factors included feather bedding (NAA 0.63 [0.52-0.76]; Atopy 0.87 [0.76-0.99]; AA 0.64 [0.52-0.79]) and feather pillows (NAA 0.80 [0.66-0.96]; Atopy 0.88 [0.77-1.01]; AA 0.64 [0.50-0.81]) which were both associated with a lower risk of asthma and atopy across all pathways. Conversely synthetic bedding was associated with a higher risk (NAA 1.33 [1.16-1.53]; Atopy 1.18 [1.05-1.31]; AA 1.51 [1.28-1.79]). (Supplementary Figure S5)

Current use of coal/coke heating was associated with an increased risk of developing AA among atopic individuals (1.75 [1.11-2.78]) but other heating factors did not show a strong association with asthma or atopy. (Supplementary Figure S6)

There was evidence that having pet birds in the first year of life was associated with higher risk of asthma (NAA 1.38 [1.13-1.68]; AA 1.47 [1.15-1.87]), and pet dogs in the first year of life a higher risk of AA 1.41 [1.15-1.72]. Many types of pets (both current and in first year of life) were associated with a lower risk of atopy (any pets 0.87 [0.79-0.95]; cats 0.81 [0.74-0.89]; dogs 0.90 [0.82-0.99]; cats in first year 0.83 [0.72-0.95]; dogs in first year 0.73 [0.63-0.84]). Farm animal contact showed no association with either asthma or atopy. (Supplementary Figure S7)

Obesity (NAA 1.60 [1.28-2.01]; AA 1.55 [1.10-2.16]) and being overweight (NAA 1.31 [1.14-1.50]; AA 1.24 [1.03-1.51]) were associated with higher risk of both forms of asthma but were not related to risk of atopy alone. Regular fruit consumption was associated with a lower risk of asthma and weakly with an increased risk of atopy (NAA 0.80 [0.70-0.93]; Atopy 1.14 [0.99-1.31]; AA 0.78 [0.63-0.97]). (Supplementary Figure S8)

Almost all indoor air-related factors including damp (NAA 1.64 [1.41-1.90]; Atopy 0.87 [0.75-1.00]; AA 1.51 [1.22-1.86]), mould (NAA 1.51 [1.29-1.75]; Atopy 0.83 [0.69-1.00]; AA 1.51 [1.20-1.91]) and maternal smoking (current: NAA 1.46 [1.32-1.62]; Atopy 0.84 [0.78-0.92]; AA 1.22 [1.07-1.40]; in first year: NAA 1.56 [1.40-1.74]; Atopy 0.87 [0.78-0.96]; AA 1.37 [1.18-1.60]) were associated with an increased risk of asthma (NAA and AA) and a slight protective effect on atopy. (Supplementary Figure S10)

Stratified by country affluence, there was still a positive correlation between risk factors for AA and NAA in both settings but some of the strongest risk factors were quite different by setting (Figure 4). The risk factors for atopy were unrelated to the risk factors of asthma in HIC but in LMIC there was evidence of a negative association (Figure 5).

**Figure 4:**
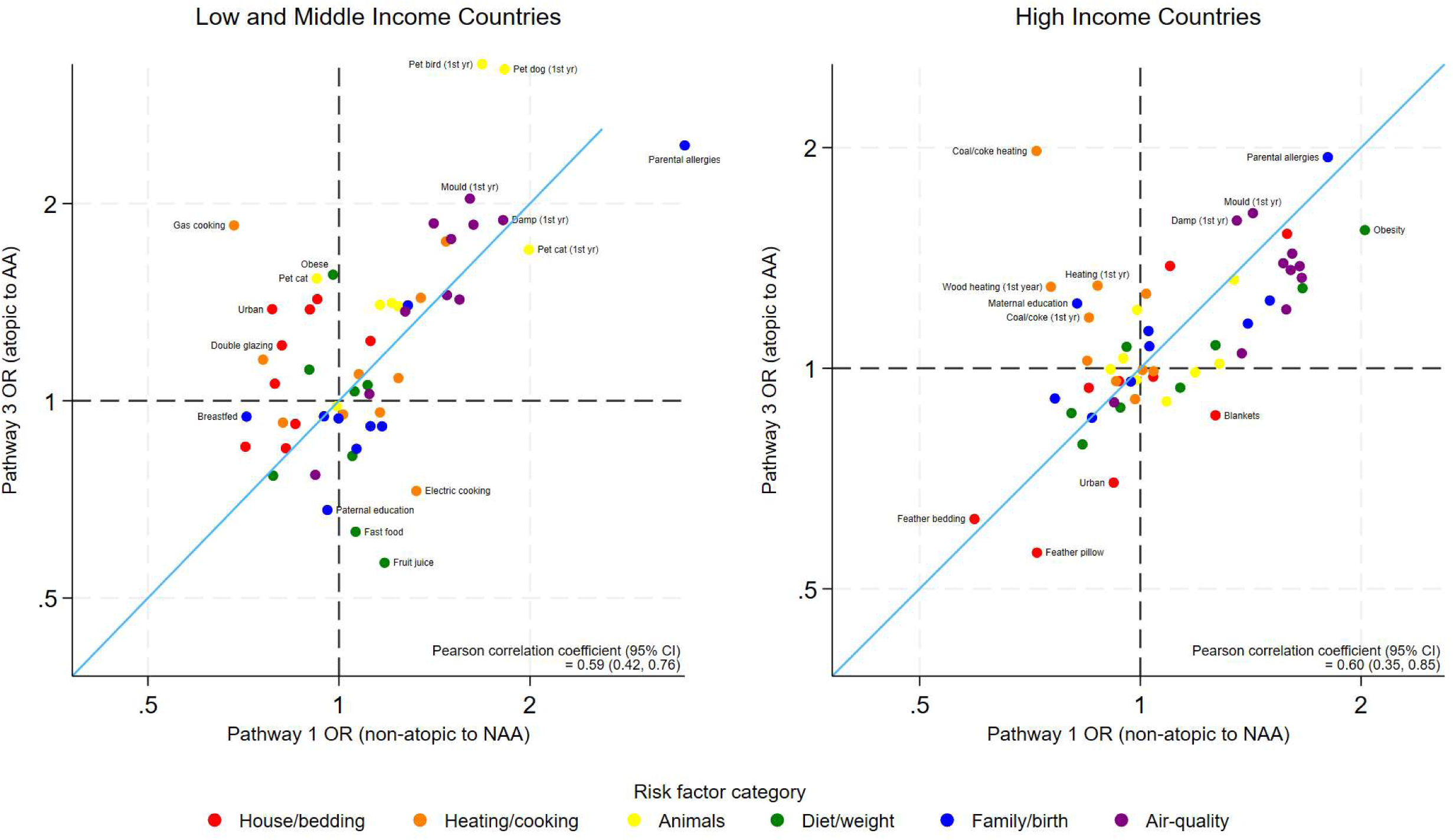
**Estimated Odds ratios (ORs) of risk factors for atopic asthma compared to non-atopic asthma, excluding atopy risk (Pathways 3 v 1) stratified by country affluence** Axes are shown on the natural log scale.

**Figure 5:**
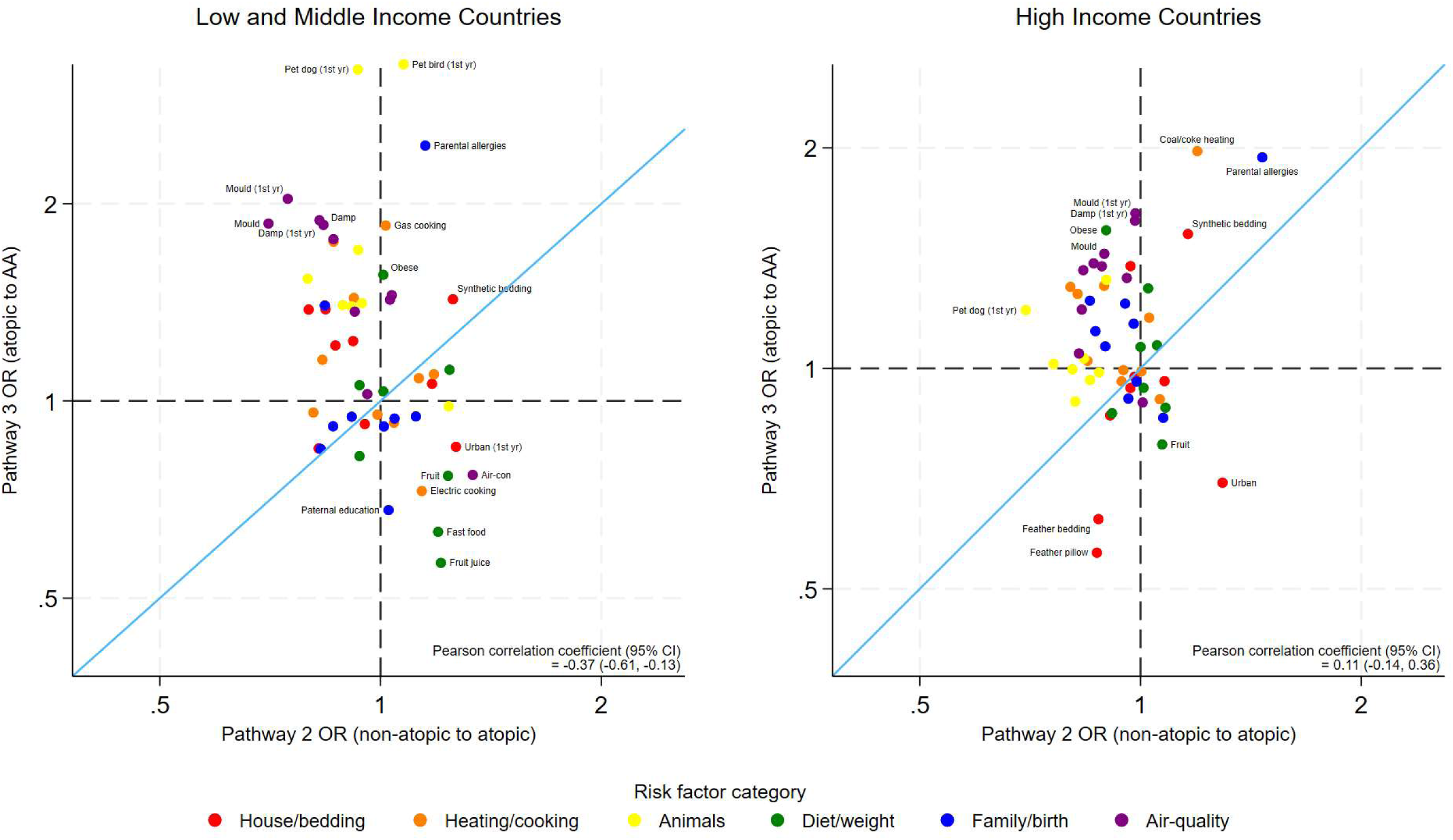
**Estimated Odds ratios (ORs) of risk factors for asthma (in atopic individuals) compared to atopy (Pathways 3 v 2) stratified by country affluence** Axes are shown on the natural log scale.

Notably, pet birds and dogs were strong risk factors for asthma in LMIC but not in HIC. Coal/coke heating was a risk factor for AA in HIC but not LMIC. The protective effect of feather bedding and pillows was only found in HIC. Regular fast food was protective for AA (but not NAA) in LMIC only. Detailed results of these stratified models are in Supplementary Table S7.

Results for the separate studies (Supplementary Tables S8 to S11) showed some differences, though the WASP study alone was relatively small, resulting in wide confidence intervals.

The sensitivity analyses with different atopy definitions showed similar outcomes. Using only ISAAC and ALSPAC but with the standard atopy definition (n=26496), the correlation between risk factors for AA in already atopic individuals (pathway 3), and NAA in non-atopic individuals (pathway 1) was lower but still very strongly positive (Pearson correlation coefficient=0.80, bootstrapped 95% CI=[0.67, 0.93]) and the correlation between risk factors for atopy (pathway 2) and AA in atopic individuals (pathway 3) was still very low (−0.12 [−0.33, 0.09]). Using wheal size >=5mm for any core allergen (n=23188), the correlation between risk factors for AA and NAA was slightly less strong (0.63 [0.45, 0.81]) and there was no correlation between risk factors for AA and atopy (−0.13 [−0.36, 0.10]). Using wheal size >=4mm for 2 or more core allergens (n=22243) resulted in similar findings (0.63 [0.45, 0.81] and −0.17 [−0.41, 0.07]). (Supplementary Figures S11-S12 and Table S12)

The second sensitivity analysis leaving out one study at a time also showed similar findings to the main analyses. The correlation between risk factors for AA and NAA was slightly lower when leaving out the ISAAC data (0.62 [0.36, 0.88]) but the confidence interval on the correlation coefficient was wider due to the lower sample size (n=9085). (Supplementary Figures S13-S16 and Table S13)

The last sensitivity analysis, allowing for a small amount of missing symptom data, increased the sample size by 3668 (n=36409) and produced similar findings to the main analyses. (Supplementary Figure S17 and Table S13)

## Discussion

We used a novel approach to assess risk factors for the different pathways to atopic and non-atopic asthma, in a large and diverse population representing both HIC and LMIC.

Our findings from the available variables indicate that lifestyle and environmental risk factors are similar for NAA (in non-atopic children) and AA (in atopic children) but not for atopy. For example, dog contact in the first year of life showed a strong association for AA in atopics (OR=1.41), weaker evidence of effect for NAA in non-atopics (1.10) and a protective association with atopy (0.73); frequent fruit intake showed strong and consistent protective associations for asthma (NAA 0.80; AA 0.78) but a positive association with atopy (1.09); maternal smoking at various times in the child’s life was strongly associated with increased risk of asthma (NAA and AA) but reduced risk of atopy. The exception was parental history of allergic diseases which was a strong risk factor for both forms of asthma (NAA 2.45; AA 2.05) and atopy (1.38).

Although atopy (defined using SPT) is associated with increased risk of asthma, other risk factors are generally the same for AA and NAA. There may be important risk factors that were not included in this study, particularly for NAA which has not been as extensively studied, and the results may have been different if these “other” risk factors (e.g., air pollution, viral infections, stress) had been examined. Nevertheless, we have included a large number of commonly recognised and studied risk factors in our analyses, including most of the risk factors that have been identified as being associated with asthma in major studies such as ISAAC.[18,19] The fact that we have found these risk factors to have similar associations with both asthma in atopics (AA) and asthma in non-atopics (NAA) raises important questions about the role of atopy in asthma, particularly whether it is an inherent part of the aetiological process, or an exacerbating factor (or a mix of both), or is coincidental. Alternatively, NAA and AA could be separate phenotypes caused by different mechanisms for the initial onset of asthma but with similar exacerbating factors. However, this cannot be established from our cross-sectional studies as it is not possible to differentiate risk factors for initial onset from those for exacerbation of pre-existing asthma.

Risk factors in the animal and air-quality categories showed a particularly striking pattern of negative associations with atopy but positive associations with asthma (both AA and NAA) (Supplementary Figures S7 and S9). This is apparent for risk factors from early life as well as current, so is unlikely to be due to reverse causation or specific to exacerbation of symptoms.

In comparing our method of investigating different pathways to the other methods listed in the introduction, we found, as an example, that fruit intake (which was protective for NAA and AA but a risk factor for atopy) was protective when comparing all asthma to all non-asthma, also when adjusting for atopy, and when comparing NAA to all people without asthma. However this effect was not seen when comparing AA to all people without asthma (as the effect was confounded by atopy). If the differences between risk factors for NAA and AA were greater than we observed, consideration of the separate pathways is even more important.

A strength of this study is the inclusion of data from 23 different countries, including both HICs and LMICs, as well as the resulting large sample size. Previous analyses of ISAAC data have generally found the risk factors for asthma to be similar in HICs and LMICs, providing for triangulation of findings,[20] and increasing the likelihood that the associations are causal. However, there are difficulties in ensuring the relevance of questions in varying settings e.g., heating questions in different climates. In addition, some risk factors may be differentially associated with socio-economic position (SEP), which we partially assessed by stratifying the results by country income level, but we do not have the information to assess differences within country.

The strongest risk factor for both types of asthma, and for atopy alone, was parental history of allergic disease, consistent with previously published findings.[9,21] Other strong risk factors for asthma (AA and NAA), but not atopy, were damp and mould in the home, which are already established risk factors for childhood asthma (both aetiology and exacerbation).[9] Obesity was a risk factor for both AA and NAA in HIC, which is consistent with literature,[9] but we did not see this in LMICs; whereas pet birds and dogs were risk factors for AA and NAA only in LMIC. These differences could indicate that associations were due to confounding, or that there were other uncaptured differences between settings e.g., bird contact could mean one pet budgie or a brood of chickens.

Other limitations of these analyses should be considered. The associations reported here will likely be subject to residual confounding as not all asthma risk factors are known or were recorded in all the included studies. Furthermore, the questionnaire was relatively simple, and most questions were binary. For some risk factors, reverse causation is a possibility, e.g., those with asthma (or family members) may have changed to synthetic bedding if feathers exacerbate symptoms. Selection bias could be a problem with males and younger people being more likely to have missing data, along with a higher chance of missing the skin prick test for those who never had asthma. Recall bias could also be an issue, with parents of children with asthma more likely to recall information on known risk factors than parents of children without asthma. There is evidence of a disassociation between SPT and IgE in LMIC where some individuals with negative SPT are IgE-positive to the same allergen.[22] In addition, the European Academy of Allergy and Clinical Immunology, in 2023, proposed a broader classification of allergy, including hypersensitivity reactions far beyond IgE-mediated phenomena (atopy).[23] Both these factors could indicate a higher proportion of asthma cases are in fact allergic (using the above definition) than that which we find here using SPT alone.

Our findings are important as they point towards similar mechanisms in AA and NAA, once the risk factors for atopy itself are removed. With SPT positivity defining atopy, there is no evidence that the risk factors differ between asthma phenotypes, even after using stricter definitions in sensitivity analyses. This suggests that atopy and asthma may be coincidental in a proportion of children who are defined as having AA. To confirm or refute these findings, further work on identifying different phenotypes of asthma should be undertaken in a variety of populations, including using other markers and data sources (e.g., microbiome and epigenetics). IgE analysis could identify patterns in potential allergens and clinical tests could investigate the role of neurological triggers.

## Contributorship

N.P. conceived of and designed the study with support from C.B., H.M., M.L.B., P.J.C., J.D., C.A.F., A.A.C. and L.P.; C.E.R and S.R. were responsible for data cleaning and management; C.E.R. analysed the data; C.E.R, N.P. and L.P. wrote the first draft of the manuscript; all other authors contributed to revisions of the manuscript; C.E.R. is the guarantor; Others also contributed to the planning and conduct of the study, and are listed below as part of the CAMERA Study Group.

## Ethics approval

Ethical approval for the study was obtained from the London School of Hygiene & Tropical Medicine (LSHTM) ethics committee (ref: 26308), the ALSPAC Ethics and Law Committee and the Local Research Ethics Committees.

## Data sharing

ISAAC data are publicly available at the UK Data Service archive http://discover.ukdataservice.ac.uk/catalogue?sn=8131.[24] ALSPAC data can be accessed after application to the ALSPAC Executive Team; application instructions and data use agreements are available at http://www.bristol.ac.uk/alspac/researchers/access/. The relevant data from the WASP study and SCAALA study will be shared on reasonable request to the corresponding author.

## Data Availability

ISAAC data are publicly available at the UK Data Service archive. ALSPAC data can be accessed after application to the ALSPAC Executive Team; application instructions and data use agreements are available. The relevant data from the WASP study and SCAALA study will be shared on reasonable request to the corresponding author.

http://discover.ukdataservice.ac.uk/catalogue?sn=8131

http://www.bristol.ac.uk/alspac/researchers/access/

## Acknowledgements

We thank all the participants, their parents, and the researchers and health workers involved in the WASP, ISAAC, SCAALA and ALSPAC studies for providing the data for this work to be completed.

## AI Use Statement

No AI was used in the creation of this manuscript.

## CAMERA Study Group

Jessica Alchundia, Hospital de Especialidades, Portoviejo, Manabi Province, Ecuador; Hajar Ali, Centre for Public Health Research, Massey University, Wellington, New Zealand; Lucia Alonzo, School of Medicine, Universidad Internacional del Ecuador (UIDE), Quito, Ecuador; Candace Andrade, Federal University of Bahia (UFBA), Salvador, Brazil; Mauricio L Barreto, The Centre for Data and Knowledge Integration for Health (CIDACS), Fiocruz, Bahia, Brazil; Collin Brooks, Centre for Public Health Research, Massey University, Wellington, New Zealand; Jeroen Burmanje, Centre for Public Health Research, Massey University, Wellington, New Zealand; Martha Chico, Fundacion Ecuatoriana Para Investigacion en Salud (FEPIS), Quito, Ecuador; Philip J Cooper, FEPIS, Quito, Ecuador, School of Medicine, UIDE, Quito, Ecuador, and Institute of Infection and Immunity, St George’s University of London, London, UK; Alvaro A Cruz, Program for Asthma Control in Bahia (ProAR), UFBA, Salvador, Brazil; Auxiliadora Damiane Pereira Vieira da Costa, Universidade Federal de Alagoas, Maceió, Brazil; Donna Davoren, Department of Medical Statistics, London School of Hygiene and Tropical Medicine (LSHTM), UK; Jeroen Douwes, Centre for Public Health Research, Massey University, Wellington, New Zealand; Maria Fernanda Arias, School of Medicine, UIDE, Quito, Ecuador; Camila A Figueiredo, Institute of Health Sciences, UFBA, Salvador, Brazil; Luciano Gama da Silva Gomes, UFBA, Salvador, Brazil; Givaneide Lima, UFBA, Salvador, Brazil; Letícia Marques dos Santos, UFBA, Salvador, Brazil; Santiago Mena, School of Medicine, UIDE, Quito, Ecuador; Harriet Mpairwe, MRC/UVRI and LSHTM Uganda Research Unit, Entebbe, Uganda; Irene Nambuya, MRC/UVRI and LSHTM Uganda Research Unit, Entebbe, Uganda; Milly Namutebi, MRC/UVRI and LSHTM Uganda Research Unit, Entebbe, Uganda; Karla Nicole, UFBA, Salvador, Brazil; Mary Njoroge, Department of Medical Statistics, LSHTM, UK; Marble Nnaluwooza, MRC/UVRI and LSHTM Uganda Research Unit, Entebbe, Uganda; Genesis Palma, School of Medicine, UIDE, Quito, Ecuador; Neil Pearce, Department of Medical Statistics, LSHTM, UK; Lucy Pembrey, Department of Medical Statistics, LSHTM, UK; Gabriela Pimentel Pinheiro, Program for Asthma Control in Bahia (ProAR), UFBA, Salvador, Brazil; David E Rebellon Sanchez, Department of Medical Statistics, LSHTM, UK; Steven Robertson, Clinical Trials Unit, LSHTM, UK; Natalia Romero-Sandoval, School of Medicine, UIDE, Quito, Ecuador, and Grups de Recerca d’Amèrica i Àfrica Llatines (GRAAL), Barcelona, Spain; Charlotte E Rutter, Department of Medical Statistics, LSHTM, UK; Bianca Sampaio Dotto Fiuza, UFBA, Salvador, Brazil; Karla Solorzano, Hospital de Especialidades, Portoviejo, Manabi Province, Ecuador; Josephine Tumusiime, MRC/UVRI and LSHTM Uganda Research Unit, Entebbe, Uganda; Pius Tumwesige, MRC/UVRI and LSHTM Uganda Research Unit, Entebbe, Uganda; Cinthia Vila Nova Santana, Program for Asthma Control in Bahia (ProAR), UFBA, Salvador, Brazil;

