## Supplementary for "Risk factors for atopic and non-atopic asthma in school-aged children from high-, and low-and-middle-income countries"

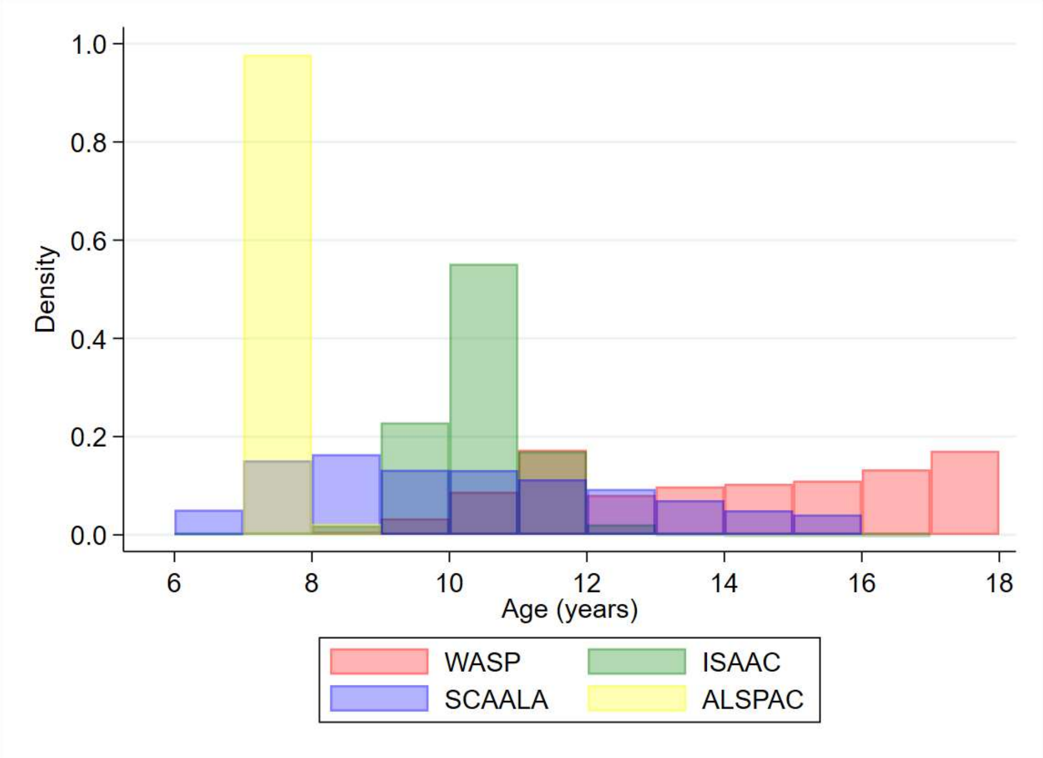

**Figure S1:** **Age distribution by study**

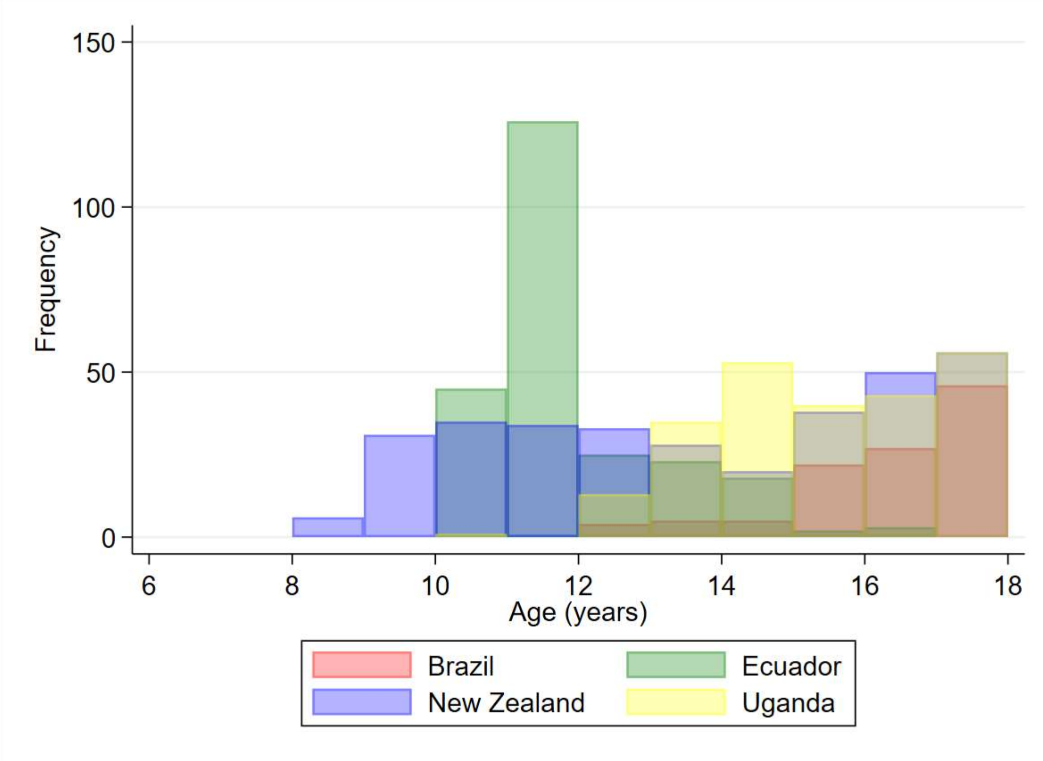

**Figure S2:** **Age distribution within World Asthma Phenotypes (WASP) study**

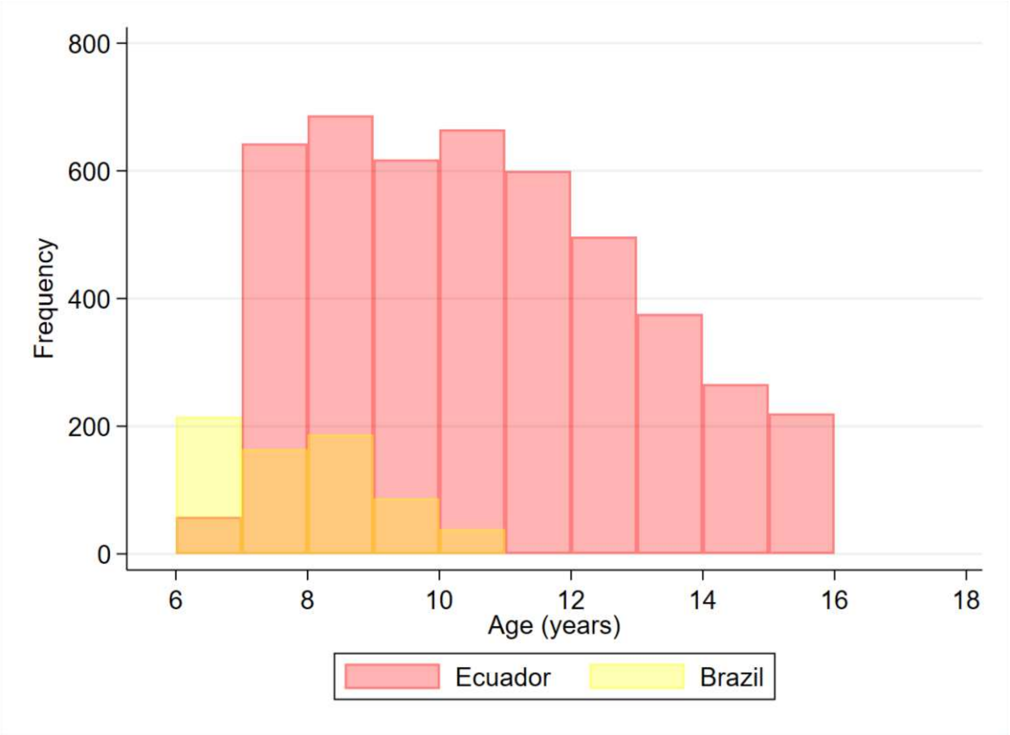

**Figure S3: Age distribution within Social Changes Asthma and Allergy in Latin America (SCAALA) study**

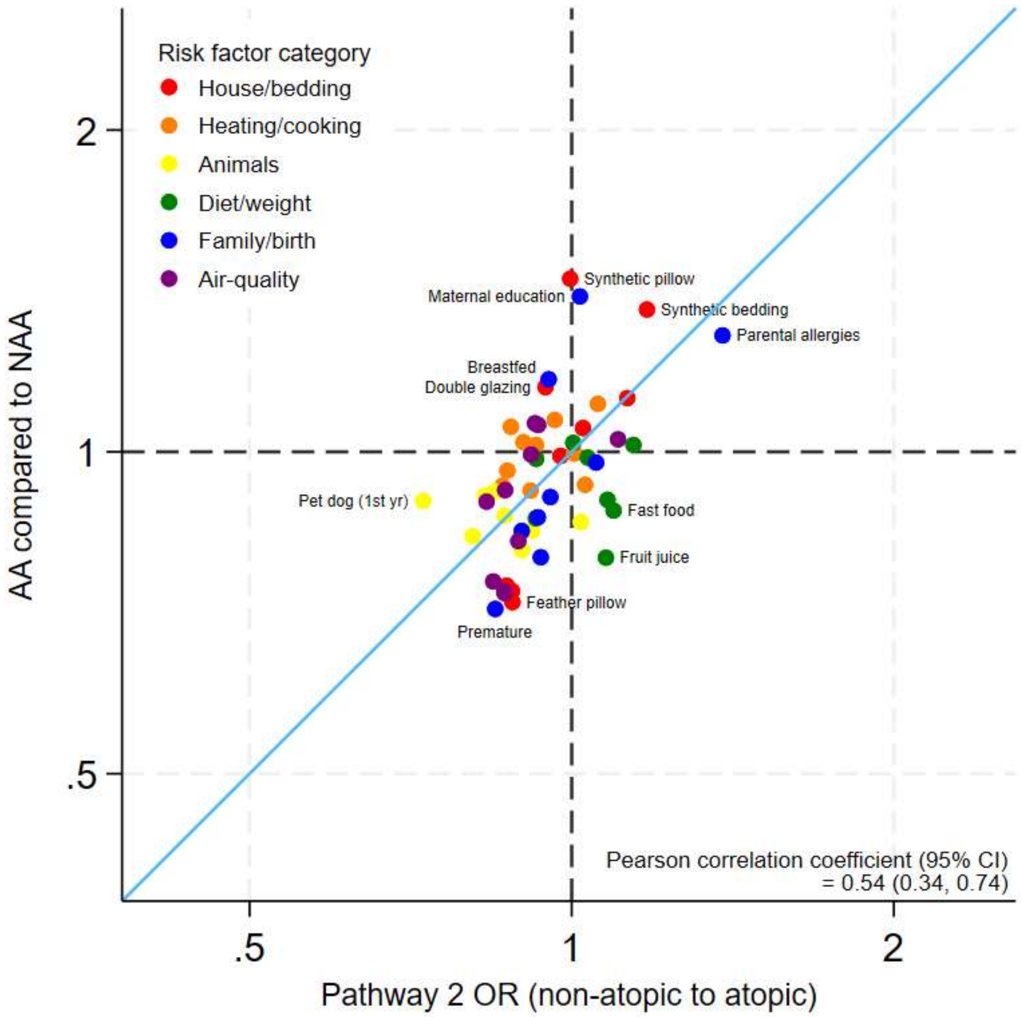

**Figure S4: Comparison of risk factors for atopy to difference between atopic asthma (AA) and non-atopic asthma (NAA)**

Axes are shown on the natural log scale.

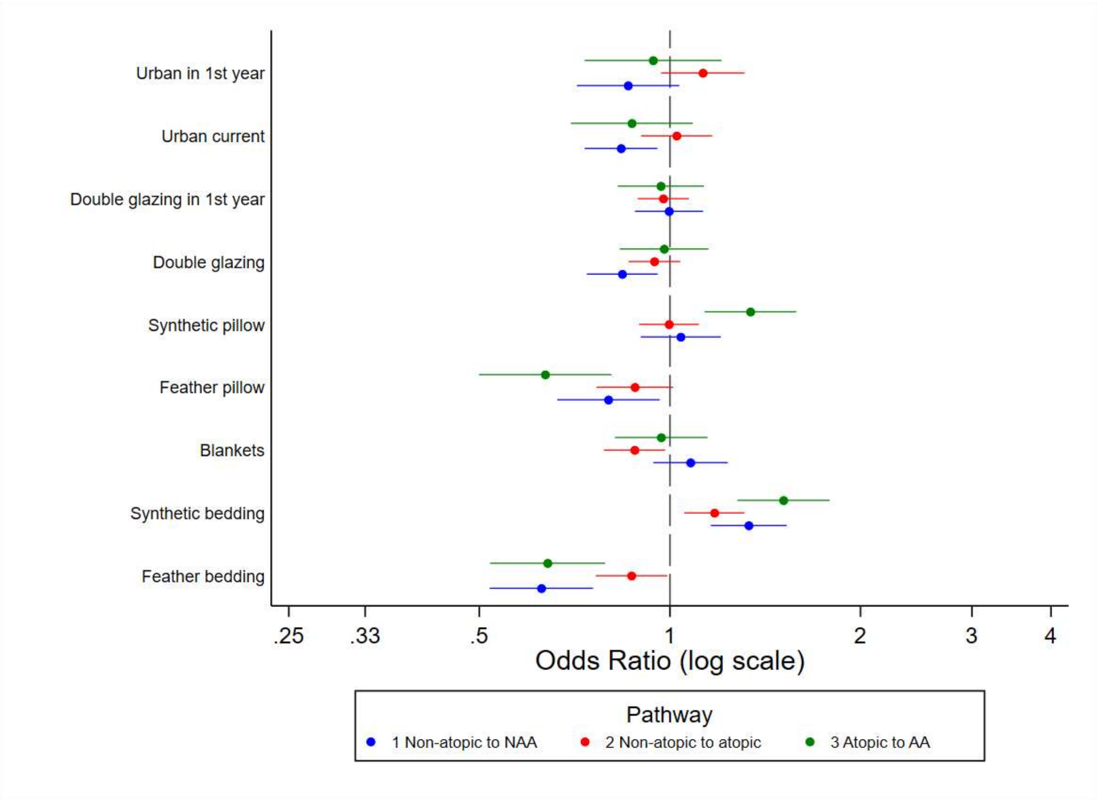

**Figure S5: Comparing pathways to asthma for house / bedding related risk factors**

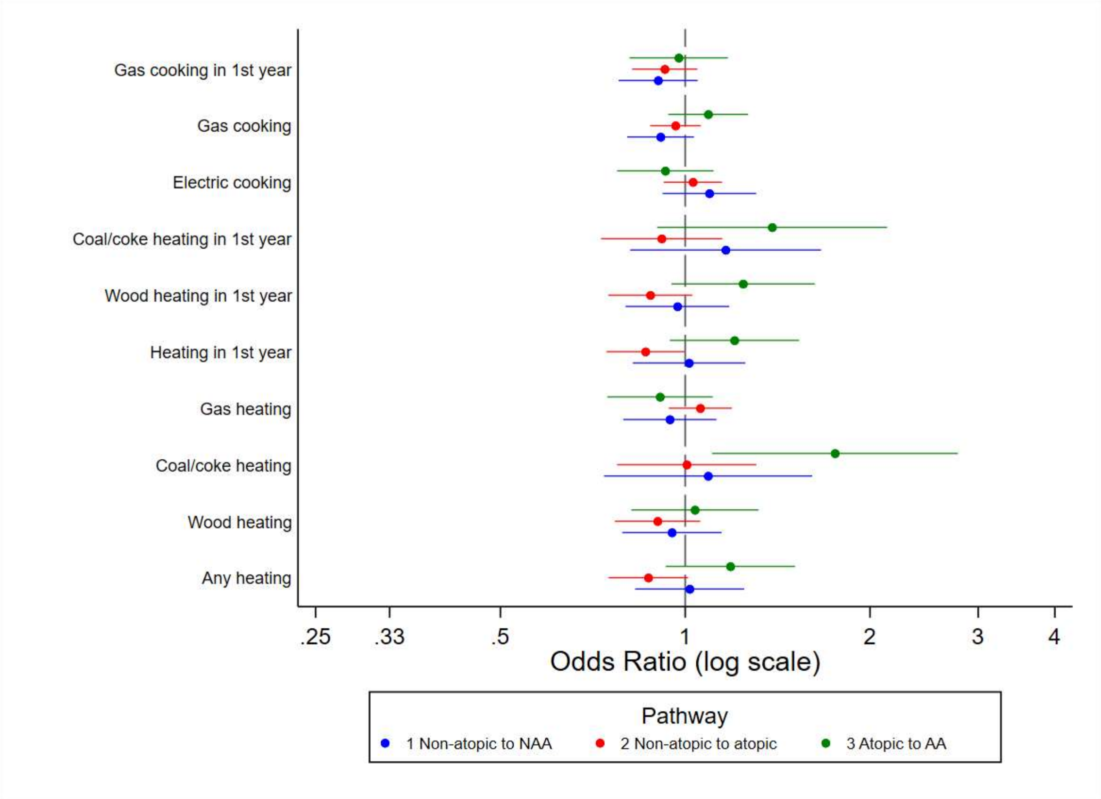

**Figure S6: Comparing pathways to asthma for heating / cooking risk factors**

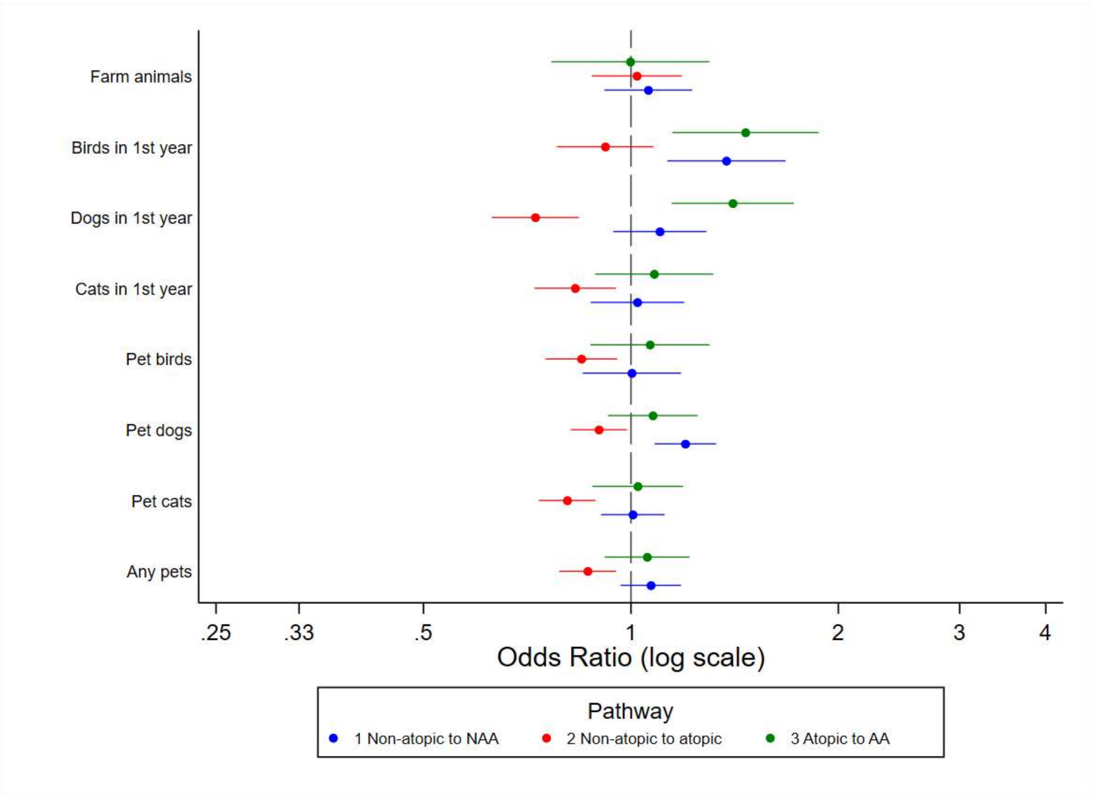

**Figure S7: Comparing pathways to asthma for animal risk factors**

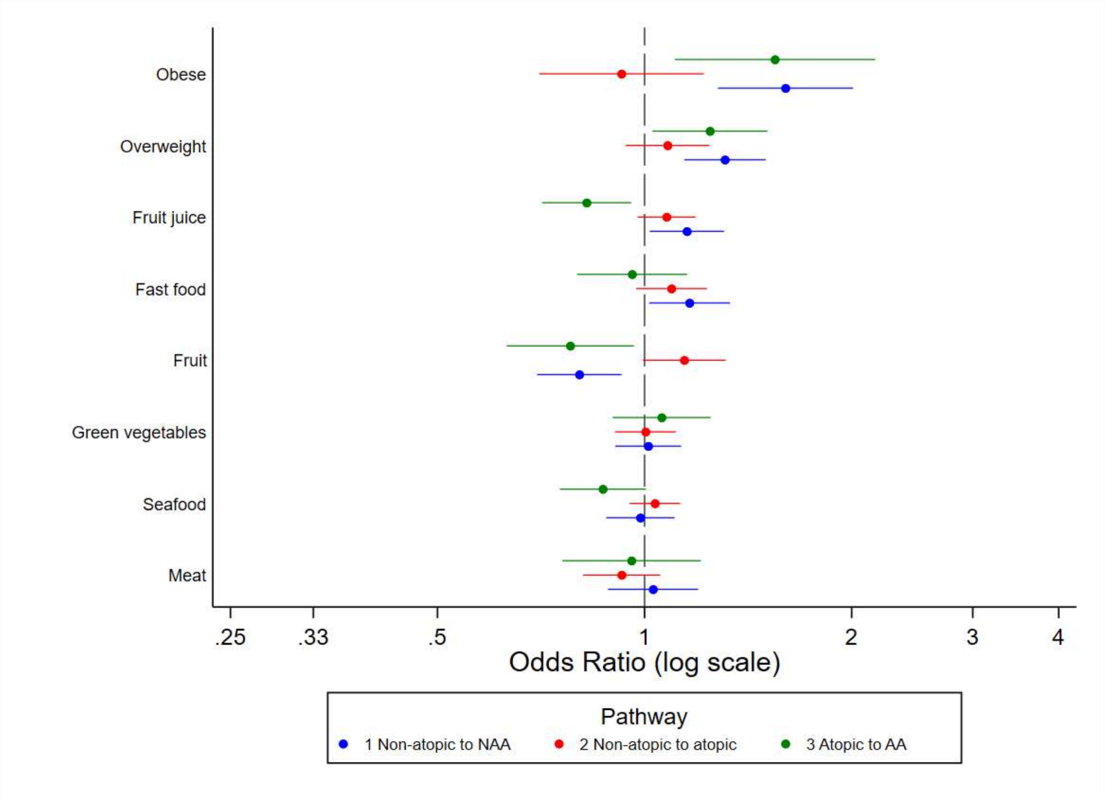

**Figure S8: Comparing pathways to asthma for dietary / weight related risk factors**

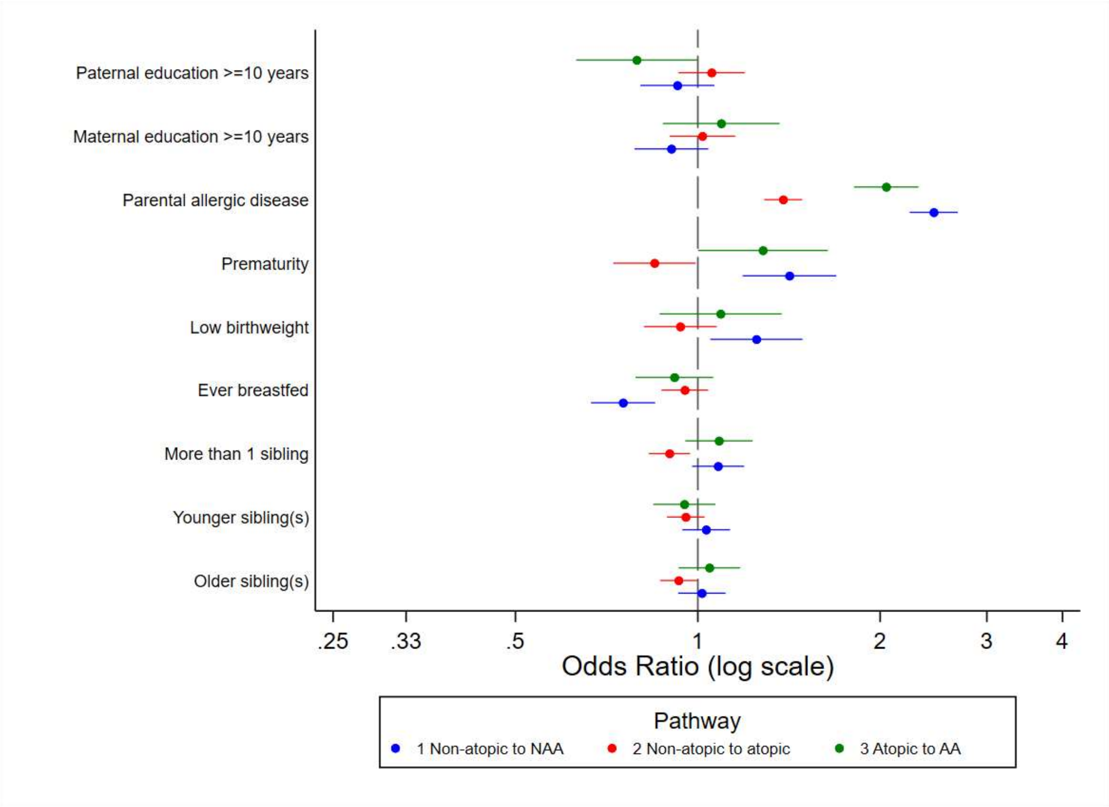

**Figure S9: Comparing pathways to asthma for family / birth related risk factors**

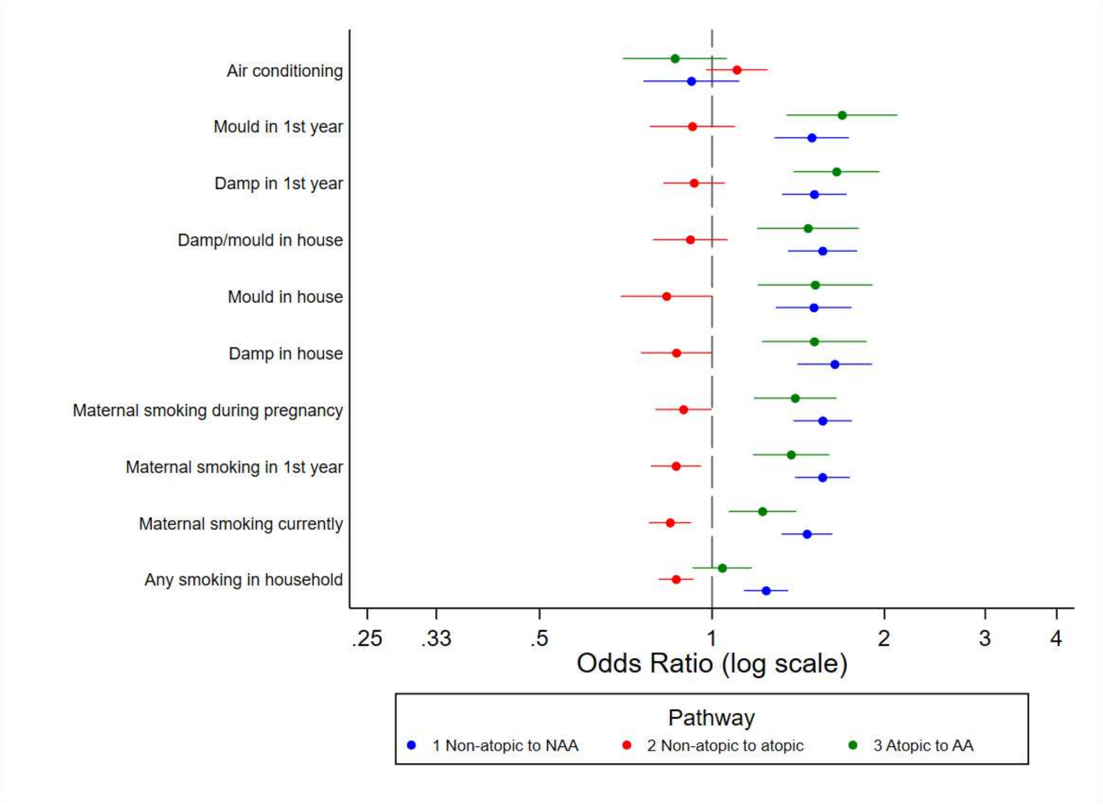

**Figure S10: Comparing pathways to asthma for air-quality related risk factors**

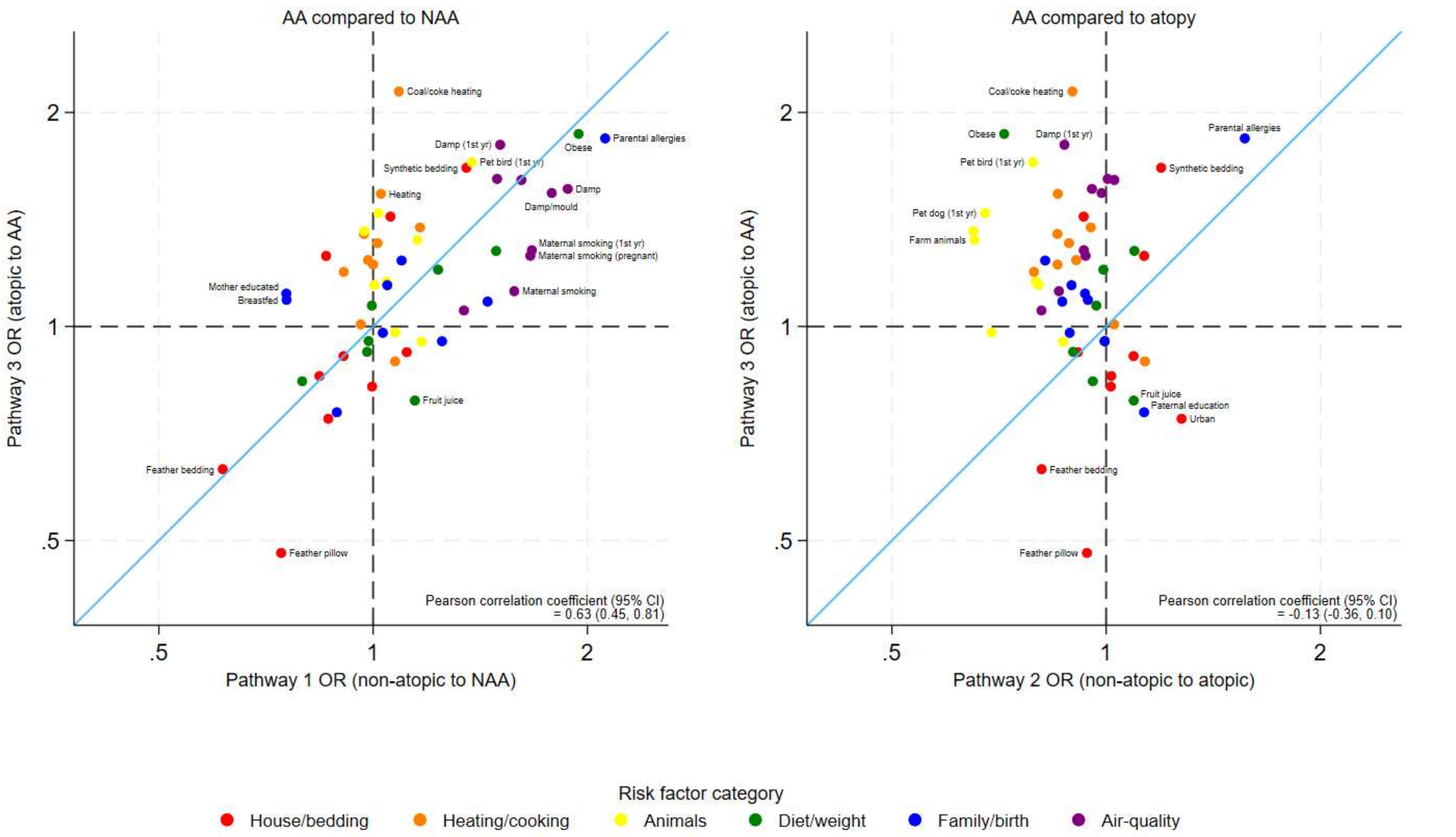

**Figure S11: Sensitivity analysis results using any wheal >=5mm in core allergens**

Axes are shown on the natural log scale. Only includes ALSPAC and ISAAC data.

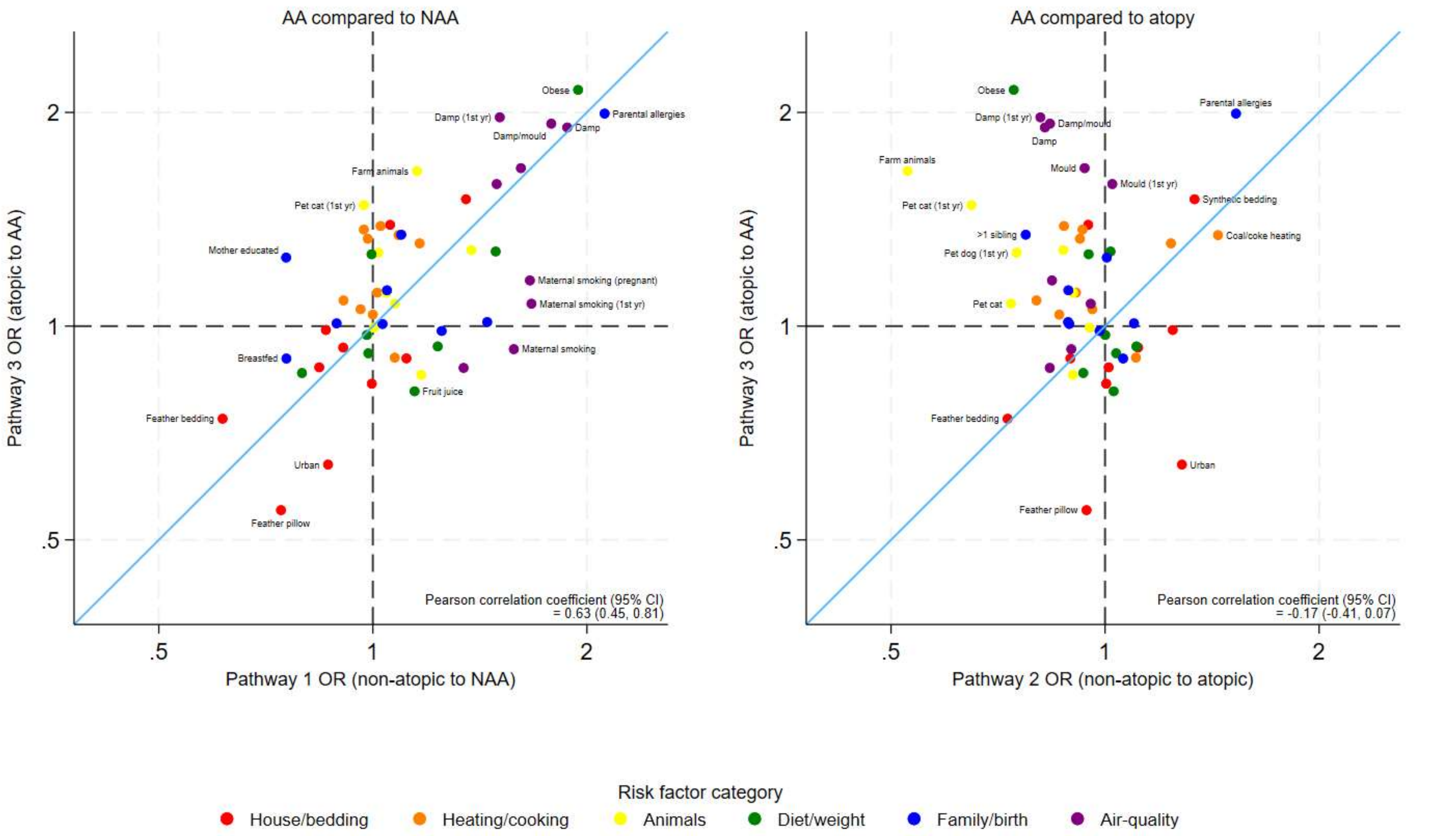

**Figure S12: Sensitivity analysis results using wheal >=4mm for 2+ core allergens**

Axes are shown on the natural log scale. Only includes ALSPAC and ISAAC data.

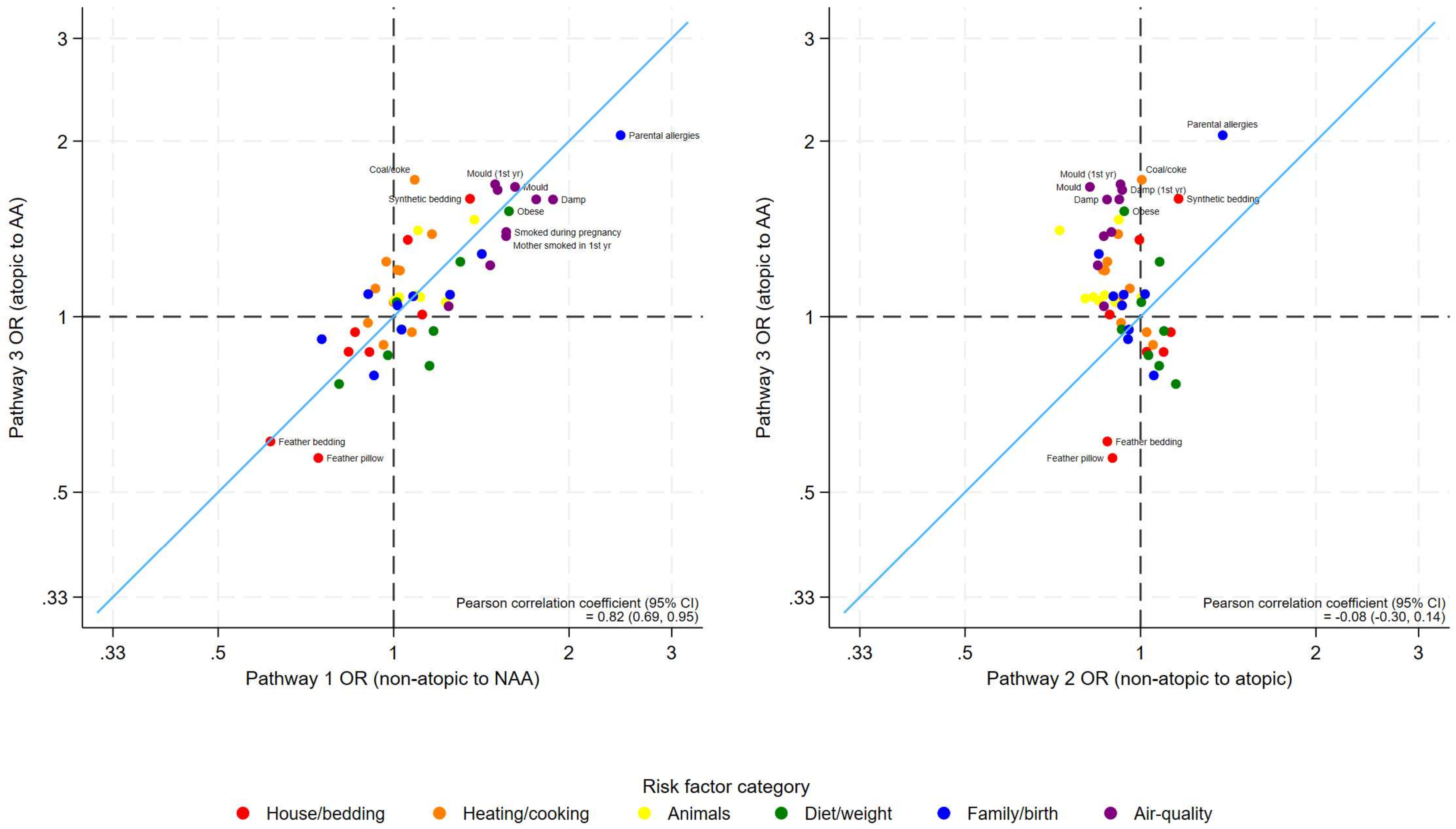

**Figure S13: Sensitivity analysis leaving out the WASP study data**

Axes are shown on the natural log scale.

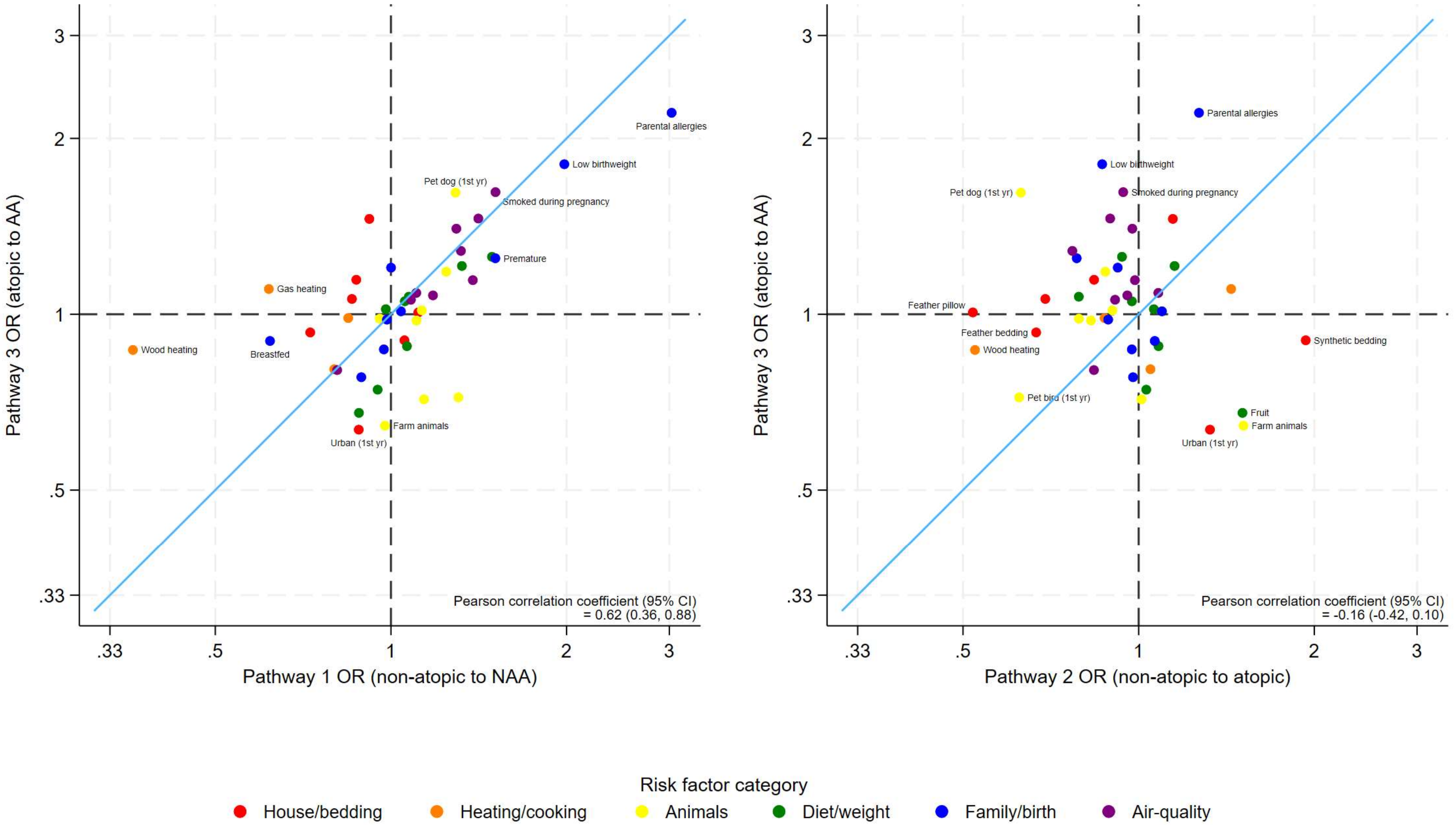

**Figure S14: Sensitivity analysis leaving out the ISAAC study data**

Axes are shown on the natural log scale.

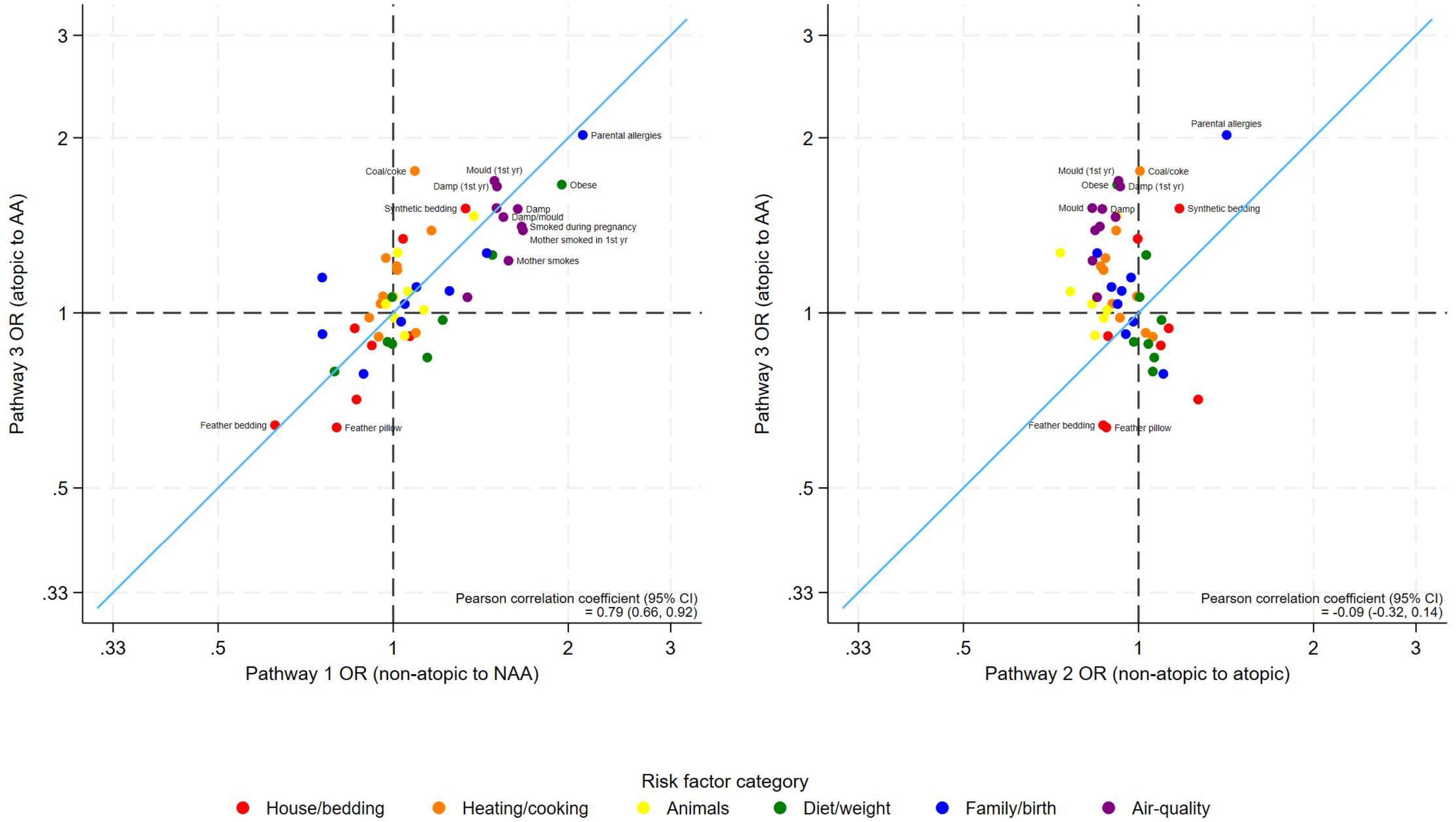

**Figure S15: Sensitivity analysis leaving out the SCAALA study data**

Axes are shown on the natural log scale.

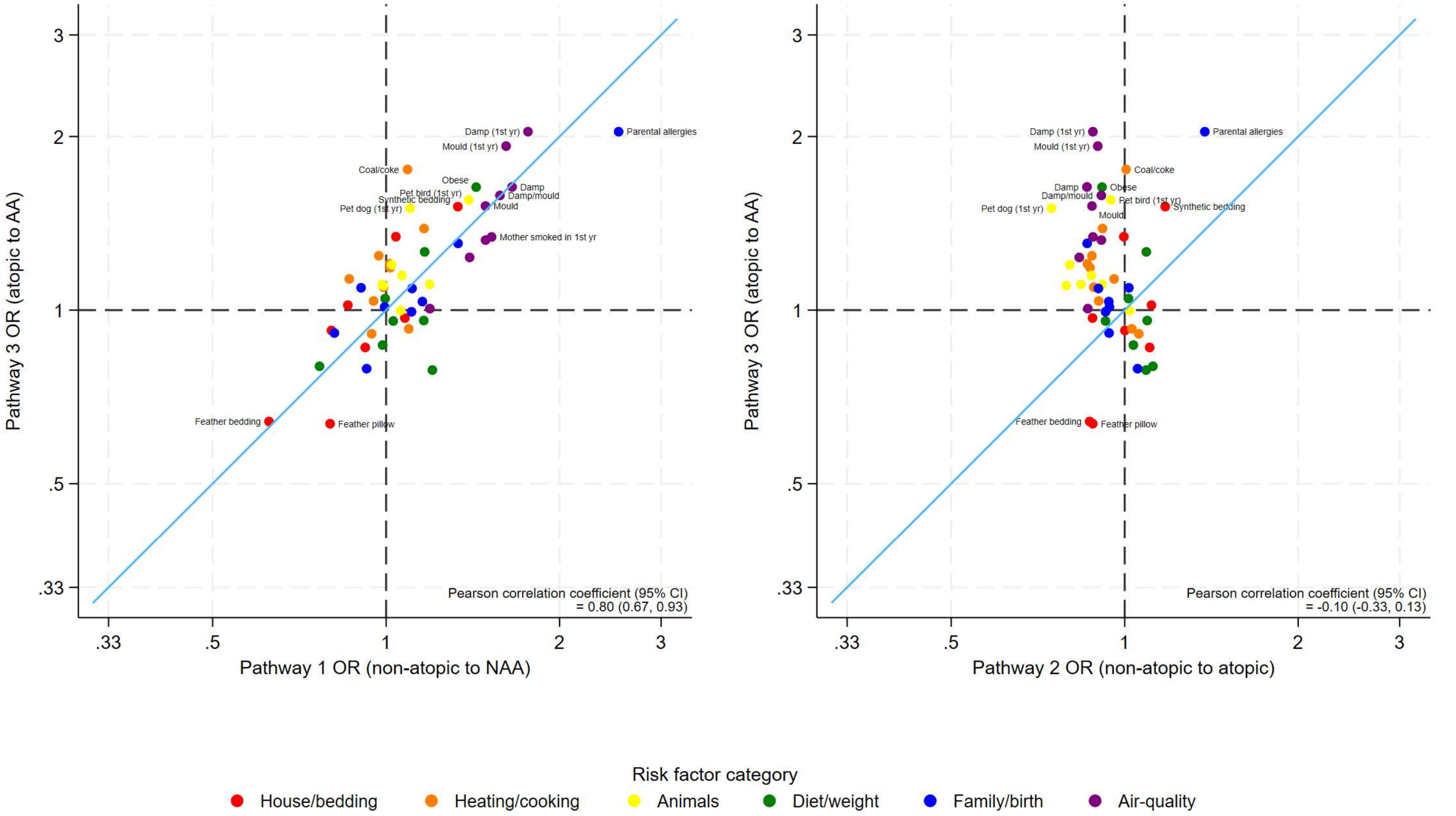

**Figure S16: Sensitivity analysis leaving out the ALSPAC study data**

Axes are shown on the natural log scale.

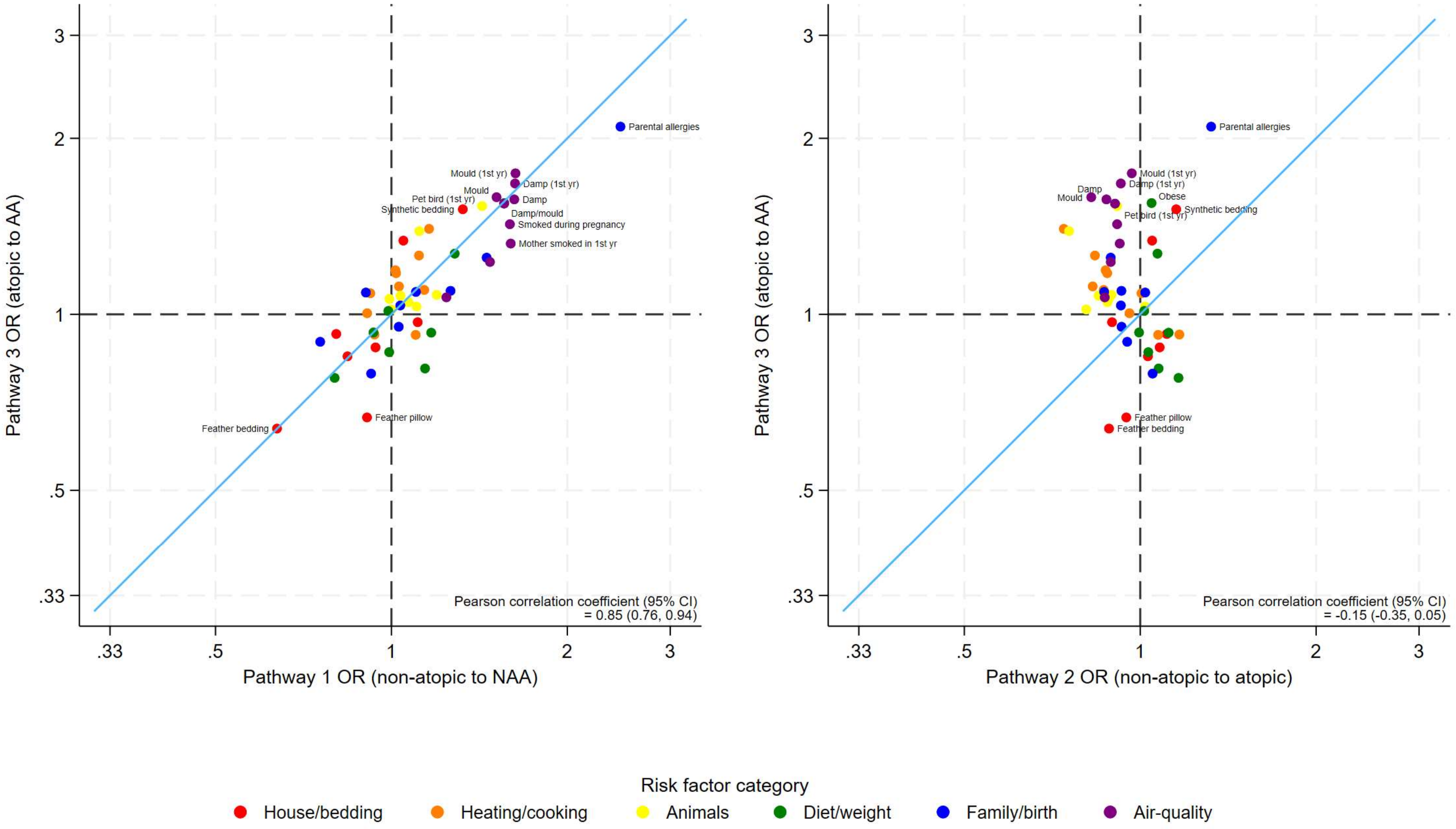

**Figure S17: Sensitivity analysis using lenient control definition (allowing some missing data)**

Axes are shown on the natural log scale.

**Table S1: Comparison of questions used to define variables across studies**

| Study: | WASP | | ISAAC | | SCAALA | | ALSPAC[a] | |
|---|---|---|---|---|---|---|---|---|
| Variable | Question | Answer | Question | Answer | | | Question | Answer |
| Current asthma | 1) Has your child ever had wheeze (a whistling sound from the chest) at any time in the past?<br>2) Has he or she had wheezing or whistling in his or her chest at any time in the past 12 months?<br>In the past 12 months, has he or she taken any medication, pills, inhalers, or other medications for asthma or wheeze? | 1) yes<br>and<br>either<br>2) yes or 3) yes | 1) Has your child ever had wheezing or whistling in the chest at any time in the past?<br>2) Has your child had wheezing or whistling in the chest in the last 12 months?<br>3) In the past 12 months, has your child used any medicines, pills, puffers or other medication for wheezing or asthma? | 1) yes or missing<br>and<br>either<br>2) yes or 3) yes | 1) Has your child ever had wheezing or whistling in the chest at any time in the past?<br>2) Has your child had wheezing or whistling in the chest in the last 12 months?<br>3) In the past 12 months, has your child used any medicines, pills, puffers or other medication for wheezing or asthma? | 1) yes or missing<br>and<br>either<br>2) yes or 3) yes | 1) Has your child had any wheezing in the past 12 months?<br>2) Has your child had asthma medication in the past 12 months? | 1) Yes @7y 7m<br>or<br>2) Yes @7y 7m |
| Never asthma | As above plus:<br>4) Has he or she ever had asthma? | 1) no and<br>2) no and<br>3) no and<br>4) no | As above plus:<br>4) Has your child ever had asthma? | 1) no and<br>2) no and<br>3) no or missing<br>and<br>4) no | As above plus:<br>4) Has your child ever had asthma? | 1) no and<br>2) no and<br>3) no or missing<br>and<br>4) no or missing | 1) Has your child had any wheezing in the past X months [since last survey]?<br>2) Has your child had asthma in the past 12 months?<br>3) In the past year has your child had any periods when there was wheezing with whistling on their chest when they breathed?<br>4) Has a doctor ever actually said that your child has asthma? | 1) No @7y 7m & @6y 9m & @5y 9m & @4y 9m & @3y 6m & @2y 6m & @1y 6m & @6m<br>and<br>2) no @7y 7m & @ 6y 9m<br>and<br>3) no @ 6y 9m<br>and<br>4) no @7y 7m |
| Atopy | Skin prick test[b] | >= 1 reaction to allergens, correct +ve and -ve controls and >=5 allergens tested | Skin prick test[b] | >= 1 reaction to allergens, correct +ve and -ve controls<br>and >=5 allergens tested | Skin prick test[b] | >= 1 reaction to allergens, correct +ve and -ve controls<br>and >=5 allergens tested | Skin prick test[b] from clinic at age 7y 6m | >= 1 reaction to allergens, correct +ve and -ve controls<br>and >=5 allergens tested |
| Non-atopic | Skin prick test[b] | No reaction to any | Skin prick test[b] | No reaction to any allergens, | Skin prick test[b] | No reaction to any allergens, | Skin prick test[b] from clinic at age 7y 6m | No reaction to any allergens, |

| | | | | | | | | |
|---|---|---|---|---|---|---|---|---|
| | | allergens, correct +ve and -ve controls and >=5 allergens tested | | correct +ve and -ve controls and >=5 allergens tested | | correct +ve and -ve controls and >=5 allergens tested | | correct +ve and -ve controls and >=5 allergens tested |
| House / bedding | | | | | | | | |
| Feather bedding | Does your child use a feather/ down duvet or quilt on *his or her* bed? | Yes/No | What kind of bedding does or did your child use? Feather quilt at present? | Yes/No | | | | |
| Synthetic bedding | Does your child use a synthetic duvet or quilt on *his or her* bed? | Yes/No | What kind of bedding does or did your child use? Synthetic quilt at present? | Yes/No | | | | |
| Blankets | Does your child use blankets on *his or her* bed? | Yes/No | What kind of bedding does or did your child use? Blankets at present? | Yes/No | Does your child use blankets at present? | Yes/No | | |
| Feather pillow | Does your child use a feather/down pillow on *his or her* bed? | Yes/No | What kind of pillow does or did your child use? Feather pillow at present? | Yes/No | | | | |
| Synthetic pillow | Does your child use a synthetic pillow on *his or her* bed? | Yes/No | What kind of bedding does or did your child use? Synthetic pillow at present? | Yes/No | | | | |
| Double glazing | | | What kind of windows are there in your child's bedroom? | Yes = Yes to sealed unit/double glazing or secondary window No = no to both | | | Are any of your windows double glazed? (including secondary double glazing) | Yes = All of them, OR Some of them No = None of them |
| Double glazing in 1st year | | | What kind of windows were there in your child's bedroom? | Yes = Yes to sealed unit/double glazing or secondary window No = no to both | | | Are any of your windows double glazed? (including secondary double glazing) | Yes = All of them, OR Some of them No = None of them |
| Urban | | | How would you describe the surroundings of your child's home at present? | Urban = Suburban, with many parks or gardens OR Suburban, with few parks or gardens OR Urban | | | Postcode matched to 2001 census urban/rural indicator | Urban = "urban (pop>10k)" OR "town and fringe" Rural = "village" OR "hamlet and isolated dwelling" |

| | | | | | | | | |
|---|---|---|---|---|---|---|---|---|
| | | | | with no parks or gardens<br>Rural = Rural, open spaces or fields nearby | | | | |
| Urban in first year | | | How would you describe the surroundings of your child's home during the child's first year of life? | Urban = Suburban, with many parks or gardens OR Suburban, with few parks or gardens OR Urban with no parks or gardens<br>Rural = Rural, open spaces or fields nearby | | | Postcode matched to 2001 census urban/rural indicator | Urban = "urban (pop>10k)" OR "town and fringe"<br>Rural = "village" OR "hamlet and isolated dwelling" |
| Heating / cooking | | | | | | | | |
| Heating | In the past 12 months have you used home heating? | Yes/No | Is your child's home heated at present? | Yes (one or more fire, stove or boiler inside or outside the home)/<br>No (not heated) | | | | |
| Heating in the first year | | | Was your child's home heated during the child's first year of life? | Yes (one or more fire, stove or boiler inside or outside the home)/<br>No (not heated) | | | | |
| Wood for heating | In the past 12 months have you used wood for heating? | Yes/No | Which fuel do you use for heating?<br>Wood (at present)? | Yes/No | | | | |
| Wood for heating in the first year | | | Which fuel did you use for heating?<br>Wood (during the child's first year of life)? | Yes/No | | | | |
| Coal or coke for heating | In the past 12 months have you used coal or coke fire? | Yes/No | Which fuel did you use for heating?<br>Coal or coke (at present)? | Yes/No | | | | |
| Coal or coke for heating in the first year | | | Which fuel did you use for heating?<br>Coal or coke (during the child's first year of life)? | Yes/No | | | | |

| | | | | | | | | |
|---|---|---|---|---|---|---|---|---|
| Gas for heating | In the past 12 months have you used any gas heating in your home (or homes)? | Yes/No | Is gas used for heating at present? | Yes/No | | | | |
| Electric cooking | Is electricity usually used for cooking in your house? | Yes/No | Do you use electricity for cooking at present? | Yes/No | Do you use electricity for cooking at present? | Yes/No | | |
| Gas cooking | In past 12 months have you used gas for any cooking in your home? (not gas BBQ) | Yes/No | Which fuel do you use for cooking?<br>Gas (at present)? | Yes/No | Do you use gas for cooking at present? | Yes/No | Do you use gas for cooking? | Yes = “Yes, ring(s) only” OR “yes, oven only” OR “yes, rings and oven”<br>No = “no, not at all” |
| Gas cooking in first year | | | Which fuel did you use for cooking?<br>Gas (during the child’s first year of life)? | Yes/No | Did you use gas for cooking in your child's first year of life? | Yes/No | Do you use gas for cooking? | Yes = “Yes, ring(s) only” OR “yes, oven only” OR “yes, rings and oven”<br>No = “no, not at all” |
| Animals | | | | | | | | |
| Any pets | Are there any pets present in the household? | Yes/No | Which of the following pets do you keep inside your child’s home? Do you keep a Dog / Cat / Other furry pet / Bird / Other pet inside your child’s home (at present)? | Yes=Yes to one of Dog / Cat / Other furry pet / Bird / Other pet<br>No= No to Dog & Cat & Other furry pet & Bird & Other pet | Derived from 6 questions. Do you keep a dog / cat / chicken / pigeon / parrot / other pet inside your home? | Yes if yes to any / No if no to all | Do you have any pets? | Yes/No |
| Cats | Cat (listed) | Yes/No | Which of the following pets do you keep inside your child’s home? Cats (at present)? | Yes/No | Do you keep a cat inside your home? | Yes/No | How many of the following pets do you have? Cats | Yes = 1+<br>No = 0 OR No to any pets |
| Cats in first year | | | Which of the following pets did you keep inside your child’s home? Cats (during the child’s first year of life)? | Yes/No | Did you keep a cat inside your home during your child’s 1st year of life? | Yes/No | How many of the following pets do you have? Cats | Yes = 1+<br>No = 0 OR No to any pets in first year |
| Dogs | Dog (listed) | Yes/No | Which of the following pets do you keep inside your child’s home? Dogs (at present)? | Yes/No | Do you keep a dog inside your home? | Yes/No | How many of the following pets do you have? Dogs | Yes = 1+<br>No = 0 OR No to any pets |
| Dogs in first year | | | Which of the following pets did you keep inside your child’s home? Dogs (during the child’s first year of life)? | Yes/No | Did you keep a dog inside your home during your child’s 1st year of life? | Yes/No | How many of the following pets do you have? Dogs | Yes = 1+<br>No = 0 OR No to any pets in first year |

| | | | | | | | | |
|---|---|---|---|---|---|---|---|---|
| Birds | | | Which of the following pets do you keep inside your child's home? Birds (at present)? | Yes/No | | | How many of the following pets do you have? Birds (budgerigar, parrot, etc.) | Yes = 1+<br>No = 0 OR No to any pets |
| Birds in first year | | | Which of the following pets did you keep inside your child's home? Birds (during the child's first year of life)? | Yes/No | | | How many of the following pets do you have? Birds (budgerigar, parrot, etc.) | Yes = 1+<br>No = 0 OR No to any pets in first year |
| Farm animal contact | Does your child currently have regular contact with cattle, sheep, horses, pigs, poultry, goats or other farm animals? | Yes/No derived from separate yes/no questions on each animal | Does your child have at least once a week contact with farm animals outside your child's home? | Yes/No | Does your child have any contact with farm animals? (Ecuador only) | Yes/No | | |
| Diet / weight | | | | | | | | |
| Meat | In the past 12 months, how often on average, did you eat meat (e.g. beef, lamb, chicken, pork) | Less than once per week = "Never or only occasionally"<br>At least once a week = "Once or twice per week" OR "Most or all days" | How often, on average, does your child eat meat nowadays? | Less than once per week = "Never" OR "Less than once per week"<br>At least once a week = "1-2 times per week" OR "3-6 times per week" OR "Once per day or more often" | How often, on average, does your child eat meat, nowadays? | Less than once per week = "Never" OR "Less than once per week"<br>At least once a week = "1-2 times per week" OR "3-6 times per week" OR "Once per day or more often" | | |
| Fish | In the past 12 months, how often on average, did you eat seafood (including fish) | Less than once per week = "Never or only occasionally"<br>At least once a week = "Once or twice per week" OR | How often, on average, does your child eat fish nowadays? | Less than once per week = "Never" OR "Less than once per week"<br>At least once a week = "1-2 times per week" OR "3-6 times per week" OR "Once per day or more often" | How often, on average, does your child eat fish, nowadays? | Less than once per week = "Never" OR "Less than once per week"<br>At least once a week = "1-2 times per week" OR "3-6 times per week" OR "Once per day or more often" | | |

| | | | | | | | | |
|---|---|---|---|---|---|---|---|---|
| | | "Most or all days" | | | | | | |
| Fruit | In the past 12 months, how often on average, did you eat fruit | Less than once per week = "Never or only occasionally" At least once a week = "Once or twice per week" OR "Most or all days" | How often, on average, does your child eat fresh fruit nowadays? | Less than once per week = "Never" OR "Less than once per week" At least once a week = "1-2 times per week" OR "3-6 times per week" OR "Once per day or more often" | How often, on average, does your child eat fresh fruit, nowadays? | Less than once per week = "Never" OR "Less than once per week" At least once a week = "1-2 times per week" OR "3-6 times per week" OR "Once per day or more often" | Thinking about all the food **that you provide** (**Do not include meals provided by school).** How often does your child eat 1. Fresh citrus fruit (e.g. oranges, grapefruit , satsumas, tangerines etc.) 2. Other fresh fruit (e.g. apple, banana, pear, bunch of grapes, peach etc.) | Less than once a week = Citrus fruit AND other fruits both "Never Or Rarely" OR "Once In 2 Weeks" At keast once a week = Citrus fruit AND/OR other fruits "1-3 Times A Week" OR "4-7 Times A Week" OR "> Once A Day" |
| Green vegetables | | | How often, on average, does your child eat or drink the following nowadays? 1. Raw green vegetables 2. Cooked green vegetables | Less than once a week = Raw green vegetables AND Cooked green vegetables both "Never" OR "Less than once per week" At least once a week = Raw green vegetables AND/OR Cooked green vegetables "1-2 times per week" OR "3-6 times per week" OR "Once per day or more often" | How often, on average, does your child eat any green vegetables, nowadays? | Less than once per week = "Never" OR "Less than once per week" At least once a week = "1-2 times per week" OR "3-6 times per week" OR "Once per day or more often" | Thinking about all the food **that you provide** (**Do not include meals provided by school).** How often does your child eat 1. Cabbage, brussel sprouts, spinach, broccoli and other dark green leafy vegetables 2. Other green vegetables (cauliflower, runner beans, leeks, courgettes etc.) | Less than once a week = Dark green leafy vegetables AND other green vegetables both "Never or Rarely" OR "Once In 2 Weeks" At keast once a week = Dark green leafy vegetables AND/OR other green vegetables "1-3 Times A Week" OR "4-7 Times A Week" OR "> Once A Day" |
| Fast food | In the past 12 months, how often on average, did you eat fast food/burgers | Less than once per week = "Never or only occasionally" | How often, on average, does your child eat burgers nowadays? | Less than once per week = "Never" OR "Less than once per week" At least once a week = "1-2 times | How often, on average, does your child eat burgers, nowadays? | Less than once per week = "Never" OR "Less than once per week" At least once a week = "1-2 times | | |

| | | | | | | | | |
|---|---|---|---|---|---|---|---|---|
| | | At least once a week = "Once or twice per week" OR "Most or all days" | | per week" OR "3-6 times per week" OR "Once per day or more often" | | per week" OR "3-6 times per week" OR "Once per day or more often" | | |
| Juice | | | How often, on average, does your child drink fruit juice nowadays? | Less than once per week = "Never" OR "Less than once per week"<br>At least once a week = "1-2 times per week" OR "3-6 times per week" OR "Once per day or more often" | How often, on average, does your child drink fruit juice, nowadays? | Less than once per week = "Never" OR "Less than once per week"<br>At least once a week = "1-2 times per week" OR "3-6 times per week" OR "Once per day or more often" | Thinking about all the food **that you provide** which she eats during the day, how often does she eat the following foods? **Do not include meals provided by school.**<br>Fruit juice from a tin (including tomato juice) and Pure fruit juice from a carton or freshly squeezed | Less than once a week=<br>Fruit juice from a tin AND pure fruit juice both "Never Or Rarely" OR "Once In 2 Weeks"<br>At keast once a week =<br>Fruit juice from a tin AND/OR pure fruit juice "1-3 Times A Week" OR "4-7 Times a Week" OR "> Once A Day" |
| Overweight | Based on Body mass index (kg/m²) compared to table by age | Overweight (BMI≥table) / Not overweight (BMI<table) | Based on Body mass index (kg/m²) compared to table by age | Overweight (BMI≥table) / Not overweight (BMI<table) | Based on Body mass index (kg/m²) compared to table by age | Overweight (BMI≥table) / Not overweight (BMI<table) | Based on Body mass index (kg/m²) compared to table by age | Overweight (BMI≥table) / Not overweight (BMI<table) |
| Obese | Based on Body mass index (kg/m²) compared to reference table by age | Obese (BMI≥table) / Not obese (BMI<table) | Based on Body mass index (kg/m²) compared to reference table by age | Obese (BMI≥table) / Not obese (BMI<table) | Based on Body mass index (kg/m²) compared to table by age | Obese (BMI≥table) / Not obese (BMI<table) | Based on Body mass index (kg/m²) compared to table by age | Obese (BMI≥table) / Not obese (BMI<table) |
| Family / birth | | | | | | | | |
| Any older siblings | | | Derived from number of older siblings | Yes/No | Does your child have older siblings? | Yes/No | Derived from number of older brothers and number of older sisters | Yes/No |
| Any younger siblings | | | Derived from number of younger siblings | Yes/No | Does your child have younger siblings? | Yes/No | Derived from number of younger brothers and number of younger sisters | Yes/No |
| More than 1 sibling | | | Derived from number of older siblings and number of younger siblings | Yes/No | Derived from number of older brothers, number of older sisters, number of | Yes/No | Derived from number of older brothers, number of older sisters, number of | Yes/No |

| | | | | | | | | |
|---|---|---|---|---|---|---|---|---|
| | | | | | younger brothers and number of younger sisters | | younger brothers and number of younger sisters | |
| Breastfed ever | | | Was your child ever breast fed? | Yes/No | Was your child ever breastfed? | Yes/No | Based on questions on breastfeeding throughout first year | Yes/No |
| Low birthweight | | | How much did your child weigh at birth? | Yes = less than 2500g<br>No = 2500g or more | | | Birthweight in grams | Yes = less than 2500g<br>No = 2500g or more |
| Premature | | | Was your child born within 3 weeks of the calculated date? | Premature = “No, more than 3 weeks early”<br>Term = “Yes” OR “No, more than 3 weeks late” | How many months of pregnancy when your child was born? | Yes = 8 months or less / No = 9 months or more | Weeks gestation | Yes if weeks gestation <=37<br>No if weeks gestation>=38 |
| Parental allergic diseases | | | Has the child’s mother ever had any of the following diseases?<br>Asthma, hayfever, eczema<br>Has the child’s father ever had any of the following diseases?<br>Asthma, hayfever, eczema | Yes = answer yes to any of the questions<br>No = answer no to all of the questions | Dervied from did mother have any allergic diseases, and did father have any allergic diseases? | Yes = answer yes to any of the questions<br>No = answer no to all of the questions | Has the child’s mother ever had any of the following problems: asthma, hayfever, eczema?<br>Has the mother’s partner (if is child’s father) ever had any of the following problems: asthma, hayfever, eczema? | Yes = answer yes to any of the questions<br>No = answer no to all of the questions |
| Maternal education | | | For how long did the child’s mother attend school or professional training?<br>School and college | Yes = at least 10 years education<br>No = less than 10 years | For how long did the child’s mother attend school and professional training? | Yes = complete primary education or more<br>No = incomplete primary education OR no education | What is the mother’s highest education qualification? | Yes = any secondary school qualification<br>No = no qualifications |
| Paternal education | | | For how long did the child’s father attend school or professional training?<br>School and college | Yes = at least 10 years education<br>No = less than 10 years | | Yes = complete primary education or more<br>No = incomplete primary education OR no education | What is the mum’s partner’s highest education qualification? | Yes = any secondary school qualification<br>No = no qualifications |
| Air-quality | | | | | | | | |
| Any smokers in household | | | Does anybody, at present, smoke inside your child’s home? | Yes/No | Does anybody, at present, smoke inside your child's home? | Yes/No | Based on how many people living in your household | Yes/No |

| | | | | | | | | |
|---|---|---|---|---|---|---|---|---|
| | | | | | | | (including yourself) are smokers ? | |
| Maternal smoking | | | Does your child's mother smoke at present? | Yes/No | Does your child's mother smoke at present? | Yes/No | Based on number of cigarettes smoked | Yes/No |
| Maternal smoking in first year | | | Did your child's mother smoke during the child's first year of life | Yes/No | Did your child's mother smoke in the child's first year of life? | Yes/No | Based on number of cigarettes smoked | Yes/No |
| Maternal smoking while pregnant | | | Does or did your child's mother smoke during pregnancy with your child? | Yes/No | Did your child's mother smoke during the pregnancy of your child? | Yes/No | Did you smoke regularly at any of the following times during pregnancy:<br>First trimester<br>Second trimester<br>Third trimester | Yes if yes to any<br>No if no to all |
| Damp | In the past 12 months, have you noticed dampness on walls or ceilings? | Yes/No | Does the child's home have damp spots on the walls or ceiling at present? | Yes/No | | | How much of a problem is damp or condensation? | 1 = no damp or condensation<br>2= not serious<br>3 = fairly serious<br>4 = very serious |
| Mould | In the past 12 months, have you noticed patches of mould/mildew on any surface of your home? | Yes/No | Does the child's home have visible moulds or fungus at present? | Yes/No | | | How much of a problem is mould? | 1 = no mould<br>2= not serious<br>3 = fairly serious<br>4 = very serious |
| Damp/ mould | Combine Damp and Mould question | Yes/No | Combine Damp and Mould question | Yes/No | Does the child's home have damp spots on the walls or ceiling, moulds or fungus? | Yes/No | Is there ever any damp, condensation or mould in your home? | Yes/No |
| Damp in first year | | | Did the child's home have damp spots on the walls or ceiling in the first year if their life? | Yes/No | | | How much of a problem is damp or condensation? | 1 = no damp or condensation<br>2= not serious<br>3 = fairly serious<br>4 = very serious |
| Mould in first year | | | Did the child's home have visible moulds or fungus in the first year if their life? | Yes/No | | | How much of a problem is mould? | 1 = no mould<br>2= not serious<br>3 = fairly serious<br>4 = very serious |
| Air conditioning | In the past 12 months have you used air conditioning? | Yes/No | Does your child's home have air conditioning at present? | Yes/No | Do you have air conditioning at home? | Yes/No | | |

ISAAC=International Study of Asthma and Allergies in Childhood; WASP=World Asthma Phenotypes; SCAALA=Social Changes, Asthma and Allergy in Latin America; ALSPAC= Avon Longitudinal Study of Parents and Children; [a] cohort study with multiple surveys at different ages, y=years, m=months, survey at 7y 7m used unless specified; [b] method and allergens from skin prick test detailed in table S2

**Table S2: Comparison of skin prick tests between studies**

| Allergen | ISAAC (>=3mm weal[a], 15 mins) | WASP (>=3mm weal[a], 15 Mins) | SCAALA (>=3mm weal[a], 15 Mins) | ALSPAC (>=2mm weal[a], 10 Mins) |
|---|---|---|---|---|
| negative control (diluent/saline) | core | core | core | core |
| positive control (histamine) | core | core | core | core |
| Mixed grass | core | NZ, Ecuador, Uganda | Ecuador | core |
| Dermatophagoides pteronyssinus (Der p) | core | Brazil, Ecuador, NZ | Brazil | core |
| Der p/Der f mix | | Uganda | Ecuador | |
| Cat | core | core | core | core |
| Egg white | | | | core |
| Peanut | | Uganda | | core |
| Mixed nuts | | | | core |
| Dog | optional | core | core | group A (1/3) |
| Horse | optional | | | group A (1/3) |
| Mouse | | | | group A (1/3) |
| Rabbit | | | | group A (1/3) |
| Guinea pig | | | | group A (1/3) |
| Hamster | | | | group A (1/3) |
| Soya | | | | group A (1/3) |
| Mixed tree pollen | core | NZ | | group C (1/3) |
| Dermatophagoides farinae (Der f) | core | Ecuador, Brazil | | group C (1/3) |
| Alternaria tenuis (fungus) | core | NZ, Ecuador | Ecuador | group C (1/3) |
| Cladosporium (mould) | optional | NZ | | group C (1/3) |
| Aspergillus fumigatus (mould) | | | | group C (1/3) |
| Mixed cockroach | optional | | | removed[b] |
| Latex | | | | group C (1/3) |
| Milk | | | | group C (1/3) |
| Olive | optional | | | |
| Parietaria officinalis | optional | | | |
| Mixed weeds | optional | | | |
| Bird epithelium | optional | | | |
| Turkish tree mix | optional | | | |
| Penicillium mix | | Ecuador, NZ, Uganda | | |
| Blomia Tropicalis | | Brazil, Uganda, Ecuador | Brazil | |
| Mould mix | optional | Uganda | | |
| Mixed fungi | | | core | |
| Blatela germanica (cockroach) | | Uganda, Brazil | Brazil | |
| Periplaneta Americana (cockroach) | | Ecuador, Brazil | core | |
| Anisakis simplex (nematode) | | Uganda | | |
| Fish | | | | group B (1/3) |
| Sesame | | | | group B (1/3) |
| Cashew | | | | group B (1/3) |
| Almond | | | | group B (1/3) |
| Walnut | | | | group B (1/3) |
| Hazelnut | | | | group B (1/3) |
| Brazil | | | | group B (1/3) |
| Pecan | | | | group B (1/3) |

[a]Weal size is calculated as: (max width+perpendicular width)/2 minus size of negative control weal;
[b]removed from analysis due to problem identified with variable in dataset;
ISAAC=International Study of Asthma and Allergies in Childhood; WASP=World Asthma Phenotypes; SCAALA=Social Changes, Asthma and Allergy in Latin America; ALSPAC= Avon Longitudinal Study of Parents and Children;

**Table S3: Associations between missingness and demographics using logistic regression adjusted for study**

| Exclusion | Sex (male) | | | Age (years) | | | Asthma (Case v control) | | |
|---|---|---|---|---|---|---|---|---|---|
| | estimate | 95% CI | P value | estimate | 95% CI | P value | estimate | 95% CI | P value |
| Missing atopy data | 1.15 | (1.03, 1.29) | 0.01 | 1.07 | (1.02, 1.13) | 0.009 | 1.27 | (1.09, 1.47) | 0.002 |
| Missing symptom data[a] | 1.04 | (0.98, 1.11) | 0.23 | 0.63 | (0.60, 0.65) | <0.001 | NA | NA | NA |

CI = Confidence interval; NA = not applicable, data not available [a] only ISAAC and ALSPAC included as WASP and SCAALA had no missing data

**Table S4: Breakdown of participants by country with income classification from 2000 for ISAAC and 2018 for WASP and 2007 for SCAALA and 1998 for ALSPAC**

| Country | Participants | Income category |
|---|---|---|
| **WASP** | | |
| Brazil (br) | 109 | LMIC |
| Ecuador (ec) | 242 | LMIC |
| New Zealand (nz) | 331 | HIC |
| Uganda (ug) | 241 | LMIC |
| **ISAAC** | | |
| Albania (al) | 791 | LMIC |
| Brazil (br) | 925 | LMIC |
| China (cn) | 2001 | LMIC |
| Germany (de) | 3861 | HIC |
| Ecuador (ec) | 6 | LMIC |
| Estonia (ee) | 194 | LMIC |
| Spain (es) | 2923 | HIC |
| Georgia (ge) | 123 | LMIC |
| Ghana (gh) | 1162 | LMIC |
| Greece (gr) | 1656 | HIC |
| Hong Kong (hk) | 1225 | HIC |
| India (in) | 1511 | LMIC |
| Iceland (is) | 457 | HIC |
| Italy (it) | 1046 | HIC |
| Latvia (lv) | 235 | LMIC |
| Netherlands (nl) | 1142 | HIC |
| Norway (no) | 520 | HIC |
| New Zealand (nz) | 952 | HIC |
| Palestine (px) | 309 | LMIC |
| Sweden (se) | 1509 | HIC |
| Turkey (tr) | 345 | LMIC |
| United Kingdom (uk) | 763 | HIC |
| **SCAALA** | | |
| Brazil (br) | 692 | LMIC |
| Ecuador (ec) | 4630 | LMIC |
| **ALSPAC** | | |
| United Kingdom | 2840 | HIC |

ISAAC=International Study of Asthma and Allergies in Childhood; WASP=World Asthma Phenotypes; SCAALA=Social Changes, Asthma and Allergy in Latin America; ALSPAC= Avon Longitudinal Study of Parents and Children; LMIC=Low-middle-income country; HIC=High-income country;

**Table S5: Prevalence of potential risk factors by centre / study**

| Category | Covariable | n | ISAAC Multiple | WASP Brazil | WASP Ecuador | WASP New Zealand | WASP Uganda | WASP total | SCAALA Brazil | SCAALA Ecuador | SCAALA total | ALSPAC UK | Overall Total |
|---|---|---|---|---|---|---|---|---|---|---|---|---|---|
| Study sample size | | | 23656 | 109 | 242 | 331 | 241 | 923 | 692 | 4630 | 5322 | 2840 | 32741 |
| Confounder | Male sex | 32741 | 49% | 38% | 59% | 49% | 29% | 45% | 53% | 51% | 52% | 49% | 49% |
| House/ bedding | Feather bedding | 14156 | 18% | 4% | 18% | 37% | . | 25% | . | . | . | . | 18% |
| | Synthetic bedding | 14155 | 43% | 46% | 4% | 55% | . | 35% | . | . | . | . | 43% |
| | Blankets | 15560 | 44% | 14% | 56% | 47% | . | 45% | 22% | . | 22% | . | 43% |
| | Feather pillow | 14780 | 17% | 6% | 55% | 24% | . | 32% | . | . | . | . | 17% |
| | Synthetic pillow | 14060 | 39% | 71% | 1% | 72% | . | 47% | . | . | . | . | 40% |
| | Double glazing | 20831 | 55% | . | . | . | . | . | . | . | . | 81% | 58% |
| | Double glazing in 1st year | 20194 | 45% | . | . | . | . | . | . | . | . | 59% | 47% |
| | Urban | 23452 | 92% | . | . | . | . | . | 100% | 36% | 45% | 88% | 81% |
| | Urban in 1st year | 16953 | 89% | . | . | . | . | . | . | . | . | 93% | 89% |
| Heating/ cooking | Heating | 16979 | 68% | 0% | 0% | 100% | . | 48% | . | . | . | . | 67% |
| | Wood heating | 18386 | 14% | 0% | 0% | 40% | 0% | 14% | . | . | . | . | 14% |
| | Coal/coke heating | 17603 | 4% | 0% | 0% | 1% | 0% | 0% | . | . | . | . | 4% |
| | Gas heating | 16261 | 28% | . | . | 34% | . | 34% | . | . | . | . | 28% |
| | Heating in 1st year | 15523 | 68% | . | . | . | . | . | . | . | . | . | 68% |
| | Wood heating in 1st year | 16894 | 20% | . | . | . | . | . | . | . | . | . | 20% |
| | Coal/coke heating in 1st year | 16330 | 6% | . | . | . | . | . | . | . | . | . | 6% |
| | Cooking electric | 20728 | 45% | 1% | 15% | . | . | 11% | 0% | . | 0% | . | 43% |
| | Cooking gas | 29549 | 54% | 100% | 95% | 56% | . | 77% | 99% | 96% | 97% | 61% | 63% |
| | Cooking gas in 1st year | 18776 | 19% | . | . | . | . | . | 99% | . | 99% | 56% | 27% |
| Animals | Pets | 20342 | 43% | 51% | 63% | 77% | 30% | 58% | 37% | 60% | 57% | 70% | 51% |
| | Cats | 30397 | 15% | 19% | 36% | 48% | 10% | 31% | 10% | 37% | 33% | 30% | 20% |
| | Dogs | 30379 | 14% | 32% | 43% | 41% | 22% | 36% | 25% | 44% | 42% | 20% | 20% |
| | Birds | 24130 | 14% | . | . | . | . | . | . | . | . | 6% | 13% |
| | Cat in 1st year | 21091 | 9% | . | . | . | . | . | 7% | . | 7% | 29% | 12% |
| | Dog in 1st year | 21091 | 8% | . | . | . | . | . | 18% | . | 18% | 19% | 10% |
| | Bird in 1st year | 20399 | 7% | . | . | . | . | . | . | . | . | 5% | 7% |
| | Farm animals | 22052 | 7% | . | 77% | . | 42% | 60% | . | 22% | 22% | . | 11% |

| | | | | | | | | | | | | | |
|---|---|---|---|---|---|---|---|---|---|---|---|---|---|
| | Meat weekly | 26025 | 88% | . | 100% | 92% | . | 95% | 61% | 88% | 85% | . | 87% |
| | Seafood weekly | 26507 | 57% | . | 90% | 54% | . | 69% | 10% | 92% | 81% | . | 62% |
| | Green vegetables weekly | 24540 | 83% | . | . | . | . | . | 31% | 73% | 68% | 78% | 79% |
| Diet/ weight | Fruit weekly | 25019 | 90% | 69% | 96% | 98% | . | 93% | 50% | 93% | 87% | 96% | 90% |
| | Fast food weekly | 17878 | 25% | 94% | 11% | 47% | . | 42% | 10% | 6% | 7% | . | 20% |
| | Fruit juice weekly | 18553 | 69% | . | . | . | . | . | 55% | . | 55% | 69% | 69% |
| | Overweight | 14615 | 25% | 21% | 26% | 23% | 18% | 22% | 27% | 8% | 10% | 14% | 18% |
| | Obese | 14615 | 7% | 7% | 6% | 5% | 3% | 5% | 13% | 1% | 3% | 3% | 4% |
| | Older siblings | 29536 | 52% | . | . | . | . | . | 62% | 76% | 75% | 52% | 56% |
| | Younger siblings | 29470 | 49% | . | . | . | . | . | 42% | 77% | 72% | 52% | 53% |
| | More than 1 sibling | 29528 | 40% | . | . | . | . | . | 47% | 87% | 82% | 34% | 47% |
| | Ever breastfed | 29588 | 78% | . | . | . | . | . | 96% | 98% | 97% | 80% | 82% |
| Family/ birth | Low birthweight | 20176 | 18% | . | . | . | . | . | . | . | . | 4% | 16% |
| | Premature | 16875 | 16% | . | . | . | . | . | 8% | . | 8% | 9% | 14% |
| | Parental allergic diseases | 31175 | 36% | . | . | . | . | . | 31% | 41% | 40% | 74% | 39% |
| | Maternal education | 18189 | 79% | . | . | . | . | . | 44% | 32% | 34% | 100% | 70% |
| | Paternal education | 18389 | 80% | . | . | . | . | . | 100% | 38% | 46% | 100% | 74% |
| | Any smokers | 29371 | 45% | . | . | . | . | . | 21% | 44% | 41% | 29% | 43% |
| | Maternal smoking | 30869 | 25% | . | . | . | . | . | 11% | 13% | 13% | 16% | 22% |
| | Maternal smoking in 1st year | 27364 | 19% | . | . | . | . | . | 10% | 11% | 11% | 17% | 17% |
| | Maternal smoking during pregnancy | 28545 | 12% | . | . | . | . | . | 10% | 10% | 10% | 16% | 12% |
| Air-quality | Damp | 17670 | 14% | 38% | 38% | 15% | 30% | 28% | . | . | . | 90% | 20% |
| | Mould | 15021 | 9% | 40% | 22% | 54% | 35% | 39% | . | . | . | 52% | 15% |
| | Damp or mould | 15512 | 18% | 50% | 42% | 55% | 48% | 49% | 60% | . | 60% | 92% | 28% |
| | Damp in 1st year | 17763 | 15% | . | . | . | . | . | . | . | . | 49% | 20% |
| | Mould in 1st year | 14599 | 10% | . | . | . | . | . | . | . | . | 25% | 13% |
| | Air conditioning | 14910 | 29% | 2% | . | 3% | . | 3% | 1% | . | 1% | . | 27% |

ISAAC=International Study of Asthma and Allergies in Childhood; WASP=World Asthma Phenotypes; SCAALA=Social Changes, Asthma and Allergy in Latin America; ALSPAC= Avon Longitudinal Study of Parents and Children

**Table S6: Results from mixed effects logistic regression with random intercept for centre, adjusted for age, sex, study and country income group**

| Category | Risk factor | Pathway 1 X to NAA | | Pathway 2 X to Y | | Pathway 3 Y to AA | | Pathway 4 X to AA | | | | Pathway 5 NAA to AA | |
|---|---|---|---|---|---|---|---|---|---|---|---|---|---|
| | | *n* | OR (95% CI) | *n* | OR (95% CI) | *n* | OR (95% CI) | *n* | OR (95% CI) | predicted OR[a] | % of predicted | *n* | OR (95% CI) |
| House/ bedding | Feather bedding | 10567 | 0.63 (0.52, 0.76) | 11018 | 0.87 (0.76, 0.99) | 3589 | 0.64 (0.52, 0.79) | 10371 | 0.51 (0.42, 0.60) | 0.56 | 0.9 | 3138 | 0.75 (0.59, 0.95) |
| | Synthetic bedding | 10566 | 1.33 (1.16, 1.53) | 11017 | 1.18 (1.05, 1.31) | 3589 | 1.51 (1.28, 1.79) | 10370 | 1.87 (1.62, 2.16) | 1.78 | 1.1 | 3138 | 1.36 (1.14, 1.63) |
| | Blankets | 11909 | 1.08 (0.94, 1.23) | 12260 | 0.88 (0.79, 0.98) | 3651 | 0.97 (0.82, 1.15) | 11519 | 0.81 (0.70, 0.93) | 0.85 | 0.9 | 3300 | 0.74 (0.62, 0.88) |
| | Feather pillow | 11206 | 0.80 (0.66, 0.96) | 11618 | 0.88 (0.77, 1.01) | 3574 | 0.64 (0.50, 0.81) | 10946 | 0.52 (0.42, 0.63) | 0.56 | 0.9 | 3162 | 0.72 (0.57, 0.92) |
| | Synthetic pillow | 10500 | 1.04 (0.90, 1.20) | 10950 | 1.00 (0.89, 1.11) | 3560 | 1.34 (1.13, 1.58) | 10286 | 1.43 (1.24, 1.65) | 1.34 | 1.1 | 3110 | 1.45 (1.21, 1.74) |
| | Double glazing | 15773 | 0.84 (0.74, 0.96) | 17402 | 0.95 (0.86, 1.04) | 5058 | 0.98 (0.83, 1.15) | 15498 | 0.93 (0.81, 1.07) | 0.93 | 1.0 | 3429 | 1.15 (0.97, 1.37) |
| | Double glazing in 1st year | 15344 | 1.00 (0.88, 1.13) | 16874 | 0.98 (0.89, 1.07) | 4850 | 0.97 (0.83, 1.13) | 15068 | 0.95 (0.83, 1.08) | 0.94 | 1.0 | 3320 | 0.99 (0.84, 1.17) |
| | Urban | 18519 | 0.84 (0.73, 0.96) | 19188 | 1.02 (0.90, 1.17) | 4933 | 0.87 (0.70, 1.09) | 17609 | 0.95 (0.79, 1.13) | 0.89 | 1.1 | 4264 | 1.05 (0.85, 1.30) |
| | Urban in 1st year | 13120 | 0.86 (0.71, 1.03) | 13973 | 1.13 (0.97, 1.31) | 3833 | 0.94 (0.73, 1.21) | 12730 | 1.02 (0.83, 1.25) | 1.06 | 1.0 | 2980 | 1.12 (0.87, 1.44) |
| Heating/ cooking | Heating | 12772 | 1.02 (0.83, 1.25) | 14360 | 0.87 (0.75, 1.01) | 4207 | 1.18 (0.93, 1.51) | 12761 | 0.98 (0.79, 1.22) | 1.03 | 1.0 | 2619 | 0.96 (0.72, 1.29) |
| | Wood heating | 13677 | 0.95 (0.79, 1.15) | 14939 | 0.90 (0.77, 1.06) | 4709 | 1.04 (0.82, 1.32) | 13538 | 0.90 (0.73, 1.09) | 0.93 | 1.0 | 3447 | 1.02 (0.81, 1.28) |
| | Coal/coke heating | 13110 | 1.09 (0.74, 1.61) | 14253 | 1.01 (0.77, 1.31) | 4493 | 1.75 (1.11, 2.78) | 12948 | 1.86 (1.26, 2.75) | 1.76 | 1.1 | 3350 | 1.00 (0.69, 1.44) |
| | Gas heating | 12152 | 0.94 (0.79, 1.12) | 13439 | 1.06 (0.94, 1.19) | 4109 | 0.91 (0.75, 1.11) | 12030 | 0.99 (0.83, 1.17) | 0.96 | 1.0 | 2822 | 1.11 (0.90, 1.37) |
| | Heating in 1st year | 11900 | 1.01 (0.82, 1.25) | 13492 | 0.86 (0.74, 1.00) | 3623 | 1.20 (0.94, 1.53) | 11747 | 0.99 (0.80, 1.24) | 1.04 | 1.0 | 2031 | 0.93 (0.68, 1.26) |
| | Wood heating in 1st year | 12799 | 0.97 (0.80, 1.18) | 14263 | 0.88 (0.75, 1.03) | 4095 | 1.24 (0.95, 1.63) | 12546 | 1.00 (0.81, 1.24) | 1.09 | 0.9 | 2631 | 1.06 (0.82, 1.35) |
| | Coal/coke heating in 1st year | 12385 | 1.16 (0.81, 1.66) | 13766 | 0.92 (0.73, 1.15) | 3945 | 1.39 (0.90, 2.13) | 12117 | 1.37 (0.95, 1.98) | 1.27 | 1.1 | 2564 | 0.92 (0.62, 1.37) |
| | Cooking electric | 15748 | 1.09 (0.92, 1.30) | 17160 | 1.03 (0.92, 1.15) | 4980 | 0.93 (0.77, 1.11) | 15372 | 0.98 (0.84, 1.16) | 0.96 | 1.0 | 3568 | 0.93 (0.76, 1.15) |
| | Cooking gas | 22738 | 0.91 (0.80, 1.03) | 24210 | 0.96 (0.88, 1.06) | 6811 | 1.09 (0.94, 1.27) | 22129 | 1.04 (0.91, 1.18) | 1.05 | 1.0 | 5339 | 1.07 (0.91, 1.27) |
| | Cooking gas in 1st year | 14074 | 0.90 (0.78, 1.05) | 15223 | 0.93 (0.82, 1.05) | 4702 | 0.98 (0.81, 1.17) | 13801 | 0.92 (0.79, 1.07) | 0.90 | 1.0 | 3553 | 1.02 (0.84, 1.23) |
| Animals | Pets | 16101 | 1.07 (0.97, 1.18) | 16255 | 0.87 (0.79, 0.95) | 4241 | 1.06 (0.92, 1.22) | 15574 | 0.85 (0.75, 0.95) | 0.91 | 0.9 | 4087 | 0.87 (0.76, 1.00) |
| | Cats | 23390 | 1.01 (0.91, 1.12) | 25076 | 0.81 (0.74, 0.89) | 7007 | 1.02 (0.88, 1.19) | 22939 | 0.80 (0.71, 0.91) | 0.83 | 1.0 | 5321 | 0.83 (0.71, 0.98) |
| | Dogs | 23379 | 1.20 (1.08, 1.33) | 25058 | 0.90 (0.82, 0.99) | 7000 | 1.08 (0.93, 1.25) | 22926 | 0.91 (0.80, 1.03) | 0.97 | 0.9 | 5321 | 0.81 (0.70, 0.94) |
| | Birds | 18376 | 1.00 (0.85, 1.18) | 20396 | 0.85 (0.75, 0.96) | 5754 | 1.07 (0.87, 1.30) | 18266 | 0.90 (0.76, 1.07) | 0.90 | 1.0 | 3734 | 0.92 (0.74, 1.15) |
| | Cat in 1st year | 15808 | 1.02 (0.87, 1.19) | 17447 | 0.83 (0.72, 0.95) | 5283 | 1.08 (0.89, 1.32) | 15618 | 0.90 (0.77, 1.05) | 0.90 | 1.0 | 3644 | 0.91 (0.74, 1.12) |
| | Dog in 1st year | 15808 | 1.10 (0.94, 1.29) | 17447 | 0.73 (0.63, 0.84) | 5283 | 1.41 (1.15, 1.72) | 15618 | 0.94 (0.80, 1.10) | 1.02 | 0.9 | 3644 | 0.90 (0.73, 1.11) |
| | Bird in 1st year | 15330 | 1.38 (1.13, 1.68) | 17034 | 0.92 (0.78, 1.08) | 5069 | 1.47 (1.15, 1.87) | 15215 | 1.16 (0.94, 1.42) | 1.35 | 0.9 | 3365 | 0.84 (0.65, 1.09) |
| | Farm animals | 17447 | 1.06 (0.91, 1.23) | 18267 | 1.02 (0.88, 1.19) | 4605 | 1.00 (0.77, 1.30) | 16580 | 0.86 (0.69, 1.06) | 1.02 | 0.8 | 3785 | 0.86 (0.68, 1.08) |
| Diet/ weight | Meat weekly | 20325 | 1.03 (0.88, 1.20) | 21744 | 0.93 (0.81, 1.05) | 5700 | 0.96 (0.76, 1.21) | 19588 | 0.97 (0.79, 1.19) | 0.89 | 1.1 | 4281 | 0.99 (0.78, 1.24) |
| | Seafood weekly | 20653 | 0.99 (0.88, 1.11) | 22073 | 1.04 (0.95, 1.13) | 5854 | 0.87 (0.75, 1.00) | 19941 | 0.96 (0.84, 1.08) | 0.90 | 1.1 | 4434 | 0.99 (0.84, 1.16) |
| | Green vegetables weekly | 19289 | 1.01 (0.91, 1.13) | 20204 | 1.00 (0.91, 1.11) | 5251 | 1.06 (0.90, 1.25) | 18441 | 1.05 (0.91, 1.20) | 1.06 | 1.0 | 4336 | 1.02 (0.86, 1.21) |
| | Fruit weekly | 19436 | 0.80 (0.70, 0.93) | 20215 | 1.14 (0.99, 1.31) | 5583 | 0.78 (0.63, 0.97) | 18730 | 0.91 (0.76, 1.09) | 0.89 | 1.0 | 4804 | 1.01 (0.82, 1.25) |
| | Fast food weekly | 14118 | 1.16 (1.02, 1.33) | 14261 | 1.09 (0.97, 1.23) | 3760 | 0.96 (0.80, 1.15) | 13385 | 1.08 (0.92, 1.26) | 1.05 | 1.0 | 3617 | 0.88 (0.73, 1.07) |
| | Fruit juice weekly | 13926 | 1.15 (1.02, 1.30) | 14940 | 1.08 (0.98, 1.19) | 4627 | 0.82 (0.71, 0.96) | 13577 | 0.92 (0.81, 1.04) | 0.89 | 1.0 | 3613 | 0.80 (0.68, 0.94) |
| | Overweight | 11087 | 1.31 (1.14, 1.50) | 10709 | 1.08 (0.94, 1.24) | 3528 | 1.24 (1.03, 1.51) | 10635 | 1.24 (1.07, 1.44) | 1.34 | 0.9 | 3906 | 0.90 (0.76, 1.07) |
| | Obese | 11087 | 1.60 (1.28, 2.01) | 10709 | 0.93 (0.70, 1.22) | 3528 | 1.55 (1.11, 2.16) | 10635 | 1.35 (1.05, 1.74) | 1.43 | 0.9 | 3906 | 0.87 (0.66, 1.13) |
| Family/ birth | Older siblings | 23027 | 1.02 (0.93, 1.11) | 24657 | 0.93 (0.87, 1.00) | 6509 | 1.05 (0.93, 1.18) | 22216 | 0.89 (0.81, 0.99) | 0.97 | 0.9 | 4879 | 0.87 (0.76, 0.99) |
| | Younger siblings | 22996 | 1.03 (0.94, 1.13) | 24602 | 0.96 (0.89, 1.03) | 6474 | 0.95 (0.84, 1.07) | 22182 | 0.95 (0.85, 1.05) | 0.91 | 1.0 | 4868 | 0.91 (0.80, 1.03) |
| | More than 1 sibling | 23030 | 1.08 (0.98, 1.19) | 24646 | 0.90 (0.83, 0.97) | 6498 | 1.08 (0.95, 1.23) | 22216 | 0.90 (0.80, 1.00) | 0.97 | 0.9 | 4882 | 0.84 (0.73, 0.97) |
| | Ever breastfed | 23039 | 0.75 (0.67, 0.85) | 24632 | 0.95 (0.87, 1.04) | 6549 | 0.91 (0.79, 1.06) | 22239 | 0.92 (0.81, 1.04) | 0.87 | 1.1 | 4956 | 1.17 (0.99, 1.38) |
| | Low birthweight | 15397 | 1.25 (1.05, 1.49) | 16528 | 0.94 (0.81, 1.07) | 4779 | 1.09 (0.86, 1.38) | 15081 | 1.03 (0.85, 1.26) | 1.02 | 1.0 | 3648 | 0.80 (0.62, 1.03) |
| | Premature | 12984 | 1.42 (1.19, 1.69) | 13638 | 0.85 (0.73, 0.99) | 3891 | 1.28 (1.00, 1.64) | 12599 | 1.05 (0.86, 1.28) | 1.09 | 1.0 | 3237 | 0.71 (0.56, 0.91) |
| | Parental allergic diseases | 24285 | 2.45 (2.24, 2.69) | 26107 | 1.38 (1.29, 1.49) | 6890 | 2.05 (1.81, 2.31) | 23503 | 2.81 (2.53, 3.12) | 2.83 | 1.0 | 5068 | 1.28 (1.12, 1.47) |
| | Maternal education | 14692 | 0.90 (0.79, 1.04) | 15076 | 1.02 (0.90, 1.15) | 3497 | 1.09 (0.88, 1.36) | 13907 | 1.21 (0.99, 1.48) | 1.11 | 1.1 | 3113 | 1.40 (1.11, 1.76) |
| | Paternal education | 14865 | 0.93 (0.80, 1.07) | 15243 | 1.05 (0.93, 1.20) | 3524 | 0.79 (0.63, 1.00) | 14061 | 0.87 (0.71, 1.07) | 0.84 | 1.0 | 3146 | 0.98 (0.76, 1.26) |

| | | | | | | | | | | | | | |
|---|---|---|---|---|---|---|---|---|---|---|---|---|---|
| Air-quality | Any smokers | 22895 | 1.24 (1.14, 1.36) | 24500 | 0.87 (0.81, 0.93) | 6476 | 1.04 (0.93, 1.17) | 22118 | 0.90 (0.81, 0.99) | 0.90 | 1.0 | 4871 | 0.74 (0.65, 0.84) |
| | Maternal smoking | 24064 | 1.46 (1.32, 1.62) | 25823 | 0.84 (0.78, 0.92) | 6805 | 1.22 (1.07, 1.40) | 23302 | 1.04 (0.93, 1.17) | 1.03 | 1.0 | 5046 | 0.76 (0.65, 0.87) |
| | Maternal smoking in 1st year | 21441 | 1.56 (1.40, 1.74) | 22776 | 0.87 (0.78, 0.96) | 5923 | 1.37 (1.18, 1.60) | 20641 | 1.16 (1.02, 1.31) | 1.19 | 1.0 | 4588 | 0.74 (0.63, 0.87) |
| | Maternal smoking during pregnancy | 22270 | 1.56 (1.39, 1.75) | 23766 | 0.89 (0.80, 1.00) | 6275 | 1.40 (1.18, 1.65) | 21483 | 1.22 (1.06, 1.40) | 1.24 | 1.0 | 4779 | 0.83 (0.70, 0.98) |
| | Damp | 13028 | 1.64 (1.41, 1.90) | 13924 | 0.87 (0.75, 1.00) | 4642 | 1.51 (1.22, 1.86) | 12896 | 1.26 (1.06, 1.50) | 1.31 | 1.0 | 3746 | 0.92 (0.76, 1.12) |
| | Mould | 11302 | 1.51 (1.29, 1.75) | 11346 | 0.83 (0.69, 1.00) | 3719 | 1.51 (1.20, 1.91) | 11037 | 1.27 (1.07, 1.51) | 1.26 | 1.0 | 3675 | 0.90 (0.74, 1.09) |
| | Damp or mould | 11623 | 1.56 (1.36, 1.79) | 11712 | 0.92 (0.79, 1.06) | 3889 | 1.47 (1.20, 1.80) | 11275 | 1.31 (1.11, 1.54) | 1.35 | 1.0 | 3800 | 0.99 (0.83, 1.19) |
| | Damp in 1st year | 13363 | 1.51 (1.32, 1.72) | 14423 | 0.93 (0.82, 1.05) | 4400 | 1.65 (1.39, 1.96) | 13065 | 1.55 (1.35, 1.79) | 1.53 | 1.0 | 3340 | 1.06 (0.89, 1.26) |
| | Mould in 1st year | 11274 | 1.49 (1.29, 1.73) | 11514 | 0.92 (0.78, 1.10) | 3325 | 1.69 (1.35, 2.11) | 10839 | 1.57 (1.33, 1.86) | 1.56 | 1.0 | 3085 | 1.06 (0.87, 1.30) |
| | Air conditioning | 11116 | 0.92 (0.76, 1.12) | 12060 | 1.10 (0.98, 1.25) | 3794 | 0.86 (0.70, 1.06) | 10888 | 1.01 (0.84, 1.22) | 0.95 | 1.1 | 2850 | 1.03 (0.79, 1.34) |

OR=Odds Ratio; CI=Confidence Interval; X: non-atopic, non-asthmatic; NAA: non-atopic asthma; Y:atopic, non-asthmatic; AA: atopic asthma; [a]the OR for pathway 2 multiplied by the OR pathway 3;

**Table S7: Mixed effects logistic regression models stratified by country income group, with random intercept for centre, adjusted for age, sex and study**

| Category | Risk factor | Pathway 1 X to NAA | | | | Pathway 2 X to Y | | | | Pathway 3 Y to AA | | | |
|---|---|---|---|---|---|---|---|---|---|---|---|---|---|
| | | *n* | LMIC OR (95% CI) | HIC OR (95% CI) | Inter-action *P*-value | *n* | LMIC OR (95% CI) | HIC OR (95% CI) | Inter-action *P*-value | *n* | LMIC OR (95% CI) | HIC OR (95% CI) | Inter-action *P*-value |
| House/ bedding | Feather bedding | 10567 | 0.83 (0.52, 1.31) | 0.59 (0.48, 0.73) | 0.20 | 11018 | 0.82 (0.57, 1.19) | 0.88 (0.76, 1.01) | 0.76 | 3589 | 0.85 (0.43, 1.66) | 0.62 (0.50, 0.78) | 0.40 |
| | Synthetic bedding | 10566 | 0.92 (0.72, 1.18) | 1.59 (1.34, 1.88) | 0.00 | 11017 | 1.25 (0.96, 1.64) | 1.16 (1.03, 1.31) | 0.60 | 3589 | 1.43 (0.92, 2.23) | 1.53 (1.27, 1.83) | 0.79 |
| | Blankets | 11909 | 0.90 (0.74, 1.10) | 1.27 (1.05, 1.53) | 0.01 | 12260 | 0.80 (0.64, 1.00) | 0.91 (0.80, 1.03) | 0.32 | 3651 | 1.38 (0.98, 1.94) | 0.86 (0.71, 1.05) | 0.02 |
| | Feather pillow | 11206 | 1.12 (0.76, 1.64) | 0.72 (0.58, 0.90) | 0.05 | 11618 | 0.92 (0.67, 1.26) | 0.87 (0.75, 1.02) | 0.78 | 3574 | 1.23 (0.69, 2.22) | 0.56 (0.43, 0.73) | 0.02 |
| | Synthetic pillow | 10500 | 0.79 (0.55, 1.14) | 1.10 (0.94, 1.29) | 0.11 | 10950 | 1.18 (0.89, 1.56) | 0.97 (0.86, 1.09) | 0.22 | 3560 | 1.06 (0.64, 1.76) | 1.38 (1.16, 1.65) | 0.34 |
| | Double glazing | 15773 | 0.81 (0.63, 1.05) | 0.85 (0.73, 0.99) | 0.76 | 17402 | 0.87 (0.71, 1.06) | 0.97 (0.87, 1.08) | 0.34 | 5058 | 1.21 (0.81, 1.81) | 0.94 (0.79, 1.12) | 0.25 |
| | Double glazing in 1st year | 15344 | 0.85 (0.66, 1.11) | 1.04 (0.90, 1.20) | 0.19 | 16874 | 0.95 (0.75, 1.20) | 0.98 (0.89, 1.09) | 0.82 | 4850 | 0.92 (0.58, 1.47) | 0.97 (0.82, 1.15) | 0.83 |
| | Urban | 18519 | 0.79 (0.66, 0.93) | 0.92 (0.74, 1.14) | 0.26 | 19188 | 0.84 (0.70, 1.01) | 1.29 (1.06, 1.58) | 0.00 | 4933 | 1.38 (0.94, 2.03) | 0.70 (0.53, 0.91) | 0.00 |
| | Urban in 1st year | 13120 | 0.71 (0.51, 0.99) | 0.94 (0.74, 1.17) | 0.18 | 13973 | 1.27 (0.94, 1.70) | 1.08 (0.90, 1.29) | 0.36 | 3833 | 0.85 (0.47, 1.54) | 0.96 (0.73, 1.26) | 0.72 |
| Heating/ cooking | Heating | 12772 | 1.02 (0.77, 1.33) | 1.02 (0.75, 1.39) | 0.99 | 14360 | 0.99 (0.76, 1.29) | 0.82 (0.69, 0.98) | 0.25 | 4207 | 0.95 (0.57, 1.60) | 1.26 (0.96, 1.67) | 0.35 |
| | Wood heating | 13677 | 1.07 (0.82, 1.40) | 0.85 (0.65, 1.10) | 0.21 | 14939 | 1.18 (0.82, 1.70) | 0.85 (0.71, 1.01) | 0.10 | 4709 | 1.10 (0.63, 1.93) | 1.02 (0.79, 1.33) | 0.83 |
| | Coal/coke heating | 13110 | 1.35 (0.83, 2.18) | 0.72 (0.35, 1.51) | 0.17 | 14253 | 0.92 (0.67, 1.27) | 1.20 (0.77, 1.85) | 0.34 | 4493 | 1.44 (0.68, 3.05) | 1.98 (1.10, 3.54) | 0.51 |
| | Gas heating | 12152 | 0.82 (0.56, 1.19) | 0.98 (0.81, 1.20) | 0.39 | 13439 | 1.04 (0.80, 1.35) | 1.06 (0.93, 1.21) | 0.90 | 4109 | 0.93 (0.53, 1.60) | 0.91 (0.73, 1.12) | 0.95 |
| | Heating in 1st year | 11900 | 1.16 (0.87, 1.55) | 0.87 (0.65, 1.18) | 0.18 | 13492 | 0.81 (0.63, 1.04) | 0.89 (0.74, 1.07) | 0.54 | 3623 | 0.96 (0.59, 1.57) | 1.30 (0.98, 1.72) | 0.30 |
| | Wood heating in 1st year | 12799 | 1.24 (0.95, 1.63) | 0.76 (0.57, 1.00) | 0.01 | 14263 | 1.13 (0.83, 1.53) | 0.80 (0.67, 0.96) | 0.06 | 4095 | 1.08 (0.61, 1.92) | 1.29 (0.95, 1.75) | 0.59 |
| | Coal/coke heating in 1st year | 12385 | 1.47 (0.91, 2.38) | 0.85 (0.47, 1.53) | 0.15 | 13766 | 0.86 (0.65, 1.14) | 1.03 (0.70, 1.51) | 0.47 | 3945 | 1.75 (0.90, 3.41) | 1.17 (0.66, 2.07) | 0.37 |
| | Cooking electric | 15748 | 1.32 (0.90, 1.95) | 1.04 (0.86, 1.27) | 0.28 | 17160 | 1.14 (0.89, 1.45) | 1.00 (0.89, 1.13) | 0.36 | 4980 | 0.73 (0.49, 1.08) | 0.99 (0.81, 1.21) | 0.17 |
| | Cooking gas | 22738 | 0.68 (0.53, 0.87) | 1.01 (0.87, 1.16) | 0.01 | 24210 | 1.02 (0.84, 1.23) | 0.95 (0.85, 1.06) | 0.53 | 6811 | 1.85 (1.24, 2.77) | 0.99 (0.85, 1.17) | 0.00 |
| | Cooking gas in 1st year | 14074 | 0.76 (0.50, 1.16) | 0.93 (0.79, 1.09) | 0.39 | 15223 | 0.83 (0.60, 1.16) | 0.94 (0.83, 1.08) | 0.50 | 4702 | 1.16 (0.62, 2.15) | 0.96 (0.79, 1.16) | 0.58 |
| Animals | Pets | 16101 | 1.16 (1.00, 1.34) | 0.99 (0.86, 1.14) | 0.12 | 16255 | 0.89 (0.76, 1.04) | 0.85 (0.76, 0.96) | 0.69 | 4241 | 1.40 (1.05, 1.87) | 0.96 (0.82, 1.14) | 0.03 |
| | Cats | 23390 | 0.92 (0.79, 1.08) | 1.09 (0.94, 1.25) | 0.13 | 25076 | 0.80 (0.67, 0.94) | 0.81 (0.73, 0.92) | 0.81 | 7007 | 1.54 (1.13, 2.09) | 0.90 (0.76, 1.07) | 0.00 |
| | Dogs | 23379 | 1.21 (1.05, 1.40) | 1.19 (1.03, 1.37) | 0.85 | 25058 | 0.94 (0.80, 1.11) | 0.88 (0.78, 0.98) | 0.48 | 7000 | 1.41 (1.04, 1.91) | 0.99 (0.83, 1.17) | 0.05 |
| | Birds | 18376 | 1.24 (0.87, 1.78) | 0.95 (0.79, 1.14) | 0.19 | 20396 | 0.91 (0.66, 1.26) | 0.84 (0.74, 0.95) | 0.62 | 5754 | 1.39 (0.77, 2.53) | 1.03 (0.83, 1.27) | 0.35 |
| | Cat in 1st year | 15808 | 1.99 (1.36, 2.93) | 0.91 (0.77, 1.08) | 0.00 | 17447 | 0.93 (0.69, 1.26) | 0.81 (0.69, 0.94) | 0.41 | 5283 | 1.70 (1.03, 2.80) | 1.00 (0.80, 1.24) | 0.05 |
| | Dog in 1st year | 15808 | 1.82 (1.27, 2.62) | 0.99 (0.83, 1.18) | 0.00 | 17447 | 0.93 (0.64, 1.35) | 0.70 (0.60, 0.82) | 0.16 | 5283 | 3.21 (1.91, 5.37) | 1.20 (0.96, 1.50) | 0.00 |
| | Bird in 1st year | 15330 | 1.68 (0.94, 3.01) | 1.34 (1.09, 1.66) | 0.48 | 17034 | 1.07 (0.68, 1.69) | 0.90 (0.76, 1.07) | 0.47 | 5069 | 3.26 (1.62, 6.58) | 1.32 (1.02, 1.71) | 0.02 |
| | Farm animals | 17447 | 0.99 (0.84, 1.18) | 1.28 (0.97, 1.70) | 0.13 | 18267 | 1.24 (1.02, 1.50) | 0.76 (0.59, 0.97) | 0.00 | 4605 | 0.98 (0.67, 1.43) | 1.01 (0.70, 1.47) | 0.90 |
| Diet/ weight | Meat weekly | 20325 | 1.11 (0.93, 1.32) | 0.81 (0.60, 1.08) | 0.07 | 21744 | 0.94 (0.79, 1.11) | 0.91 (0.75, 1.11) | 0.86 | 5700 | 1.06 (0.76, 1.47) | 0.87 (0.63, 1.20) | 0.41 |
| | Seafood weekly | 20653 | 1.05 (0.88, 1.25) | 0.94 (0.81, 1.09) | 0.34 | 22073 | 0.94 (0.80, 1.09) | 1.08 (0.98, 1.20) | 0.13 | 5854 | 0.82 (0.61, 1.11) | 0.88 (0.75, 1.04) | 0.69 |
| | Fruit weekly | 19436 | 0.79 (0.66, 0.94) | 0.83 (0.66, 1.06) | 0.71 | 20215 | 1.24 (1.00, 1.52) | 1.07 (0.89, 1.29) | 0.31 | 5583 | 0.77 (0.55, 1.08) | 0.79 (0.60, 1.03) | 0.92 |
| | Green vegetables weekly | 19289 | 1.06 (0.91, 1.23) | 0.96 (0.81, 1.13) | 0.38 | 20204 | 1.01 (0.86, 1.19) | 1.00 (0.88, 1.14) | 0.94 | 5251 | 1.03 (0.77, 1.39) | 1.07 (0.88, 1.30) | 0.85 |
| | Fast food weekly | 14118 | 1.06 (0.87, 1.29) | 1.27 (1.05, 1.53) | 0.20 | 14261 | 1.20 (0.97, 1.49) | 1.05 (0.91, 1.21) | 0.33 | 3760 | 0.63 (0.42, 0.95) | 1.08 (0.87, 1.32) | 0.02 |
| | Fruit juice weekly | 13926 | 1.18 (0.97, 1.43) | 1.13 (0.96, 1.33) | 0.75 | 14940 | 1.21 (1.03, 1.42) | 1.01 (0.89, 1.14) | 0.08 | 4627 | 0.57 (0.42, 0.76) | 0.94 (0.79, 1.12) | 0.00 |
| | Overweight | 11087 | 0.90 (0.72, 1.12) | 1.66 (1.40, 1.98) | 0.00 | 10709 | 1.24 (0.96, 1.61) | 1.02 (0.87, 1.21) | 0.22 | 3528 | 1.12 (0.75, 1.67) | 1.29 (1.03, 1.60) | 0.54 |
| | Obese | 11087 | 0.98 (0.65, 1.48) | 2.02 (1.54, 2.65) | 0.00 | 10709 | 1.01 (0.60, 1.71) | 0.90 (0.65, 1.24) | 0.71 | 3528 | 1.56 (0.81, 3.01) | 1.54 (1.04, 2.28) | 0.98 |
| Family/ birth | Older siblings | 23027 | 1.00 (0.87, 1.15) | 1.03 (0.91, 1.16) | 0.75 | 24657 | 1.04 (0.90, 1.21) | 0.90 (0.83, 0.97) | 0.07 | 6509 | 0.94 (0.72, 1.23) | 1.07 (0.94, 1.22) | 0.39 |
| | Younger siblings | 22996 | 1.12 (0.98, 1.29) | 0.97 (0.86, 1.09) | 0.12 | 24602 | 0.86 (0.75, 1.00) | 0.99 (0.91, 1.07) | 0.11 | 6474 | 0.91 (0.70, 1.19) | 0.96 (0.84, 1.09) | 0.76 |
| | More than 1 sibling | 23030 | 1.17 (1.00, 1.37) | 1.03 (0.90, 1.17) | 0.21 | 24646 | 1.01 (0.86, 1.19) | 0.87 (0.79, 0.95) | 0.12 | 6498 | 0.91 (0.67, 1.24) | 1.12 (0.98, 1.29) | 0.23 |
| | Ever breastfed | 23039 | 0.72 (0.56, 0.92) | 0.76 (0.67, 0.88) | 0.65 | 24632 | 0.91 (0.75, 1.11) | 0.96 (0.87, 1.06) | 0.64 | 6549 | 0.95 (0.65, 1.37) | 0.91 (0.77, 1.07) | 0.85 |
| | Low birthweight | 15397 | 1.07 (0.81, 1.39) | 1.40 (1.12, 1.76) | 0.13 | 16528 | 0.83 (0.63, 1.08) | 0.98 (0.83, 1.15) | 0.30 | 4779 | 0.84 (0.48, 1.49) | 1.15 (0.89, 1.48) | 0.33 |
| | Premature | 12984 | 1.28 (0.96, 1.72) | 1.50 (1.20, 1.88) | 0.40 | 13638 | 0.84 (0.64, 1.10) | 0.85 (0.70, 1.03) | 0.92 | 3891 | 1.40 (0.88, 2.22) | 1.24 (0.92, 1.66) | 0.66 |
| | Parental allergic diseases | 24285 | 3.51 (3.06, 4.02) | 1.80 (1.59, 2.04) | 0.00 | 26107 | 1.15 (0.99, 1.34) | 1.47 (1.35, 1.59) | 0.01 | 6890 | 2.45 (1.89, 3.18) | 1.94 (1.69, 2.23) | 0.12 |
| | Maternal education | 14692 | 0.95 (0.80, 1.12) | 0.82 (0.64, 1.05) | 0.34 | 15076 | 1.12 (0.92, 1.35) | 0.95 (0.81, 1.12) | 0.21 | 3497 | 0.95 (0.68, 1.32) | 1.23 (0.91, 1.66) | 0.26 |

| | | | | | | | | | | | | | |
|---|---|---|---|---|---|---|---|---|---|---|---|---|---|
| | Paternal education | 14865 | 0.96 (0.81, 1.14) | 0.86 (0.67, 1.10) | 0.47 | 15243 | 1.03 (0.84, 1.25) | 1.07 (0.91, 1.26) | 0.72 | 3524 | 0.68 (0.46, 1.02) | 0.86 (0.64, 1.14) | 0.36 |
| Air-quality | Any smokers | 22895 | 1.12 (0.98, 1.27) | 1.38 (1.21, 1.56) | 0.02 | 24500 | 0.96 (0.85, 1.09) | 0.82 (0.76, 0.90) | 0.05 | 6476 | 1.02 (0.81, 1.30) | 1.05 (0.91, 1.20) | 0.87 |
| | Maternal smoking | 24064 | 1.27 (1.07, 1.51) | 1.58 (1.39, 1.79) | 0.05 | 25823 | 0.92 (0.74, 1.14) | 0.83 (0.76, 0.91) | 0.39 | 6805 | 1.37 (0.95, 1.96) | 1.20 (1.04, 1.39) | 0.52 |
| | Maternal smoking in 1st year | 21441 | 1.48 (1.23, 1.79) | 1.60 (1.40, 1.84) | 0.50 | 22776 | 1.04 (0.81, 1.32) | 0.84 (0.75, 0.93) | 0.12 | 5923 | 1.45 (0.97, 2.16) | 1.36 (1.15, 1.61) | 0.78 |
| | Maternal smoking during pregnancy | 22270 | 1.55 (1.27, 1.88) | 1.57 (1.35, 1.81) | 0.93 | 23766 | 1.03 (0.80, 1.33) | 0.86 (0.76, 0.98) | 0.23 | 6275 | 1.43 (0.94, 2.18) | 1.39 (1.16, 1.67) | 0.91 |
| | Damp | 13028 | 1.63 (1.34, 1.98) | 1.65 (1.31, 2.08) | 0.94 | 13924 | 0.84 (0.66, 1.06) | 0.89 (0.74, 1.06) | 0.70 | 4642 | 1.86 (1.27, 2.72) | 1.38 (1.07, 1.77) | 0.20 |
| | Mould | 11302 | 1.41 (1.14, 1.75) | 1.61 (1.30, 2.00) | 0.39 | 11346 | 0.70 (0.50, 1.00) | 0.89 (0.72, 1.11) | 0.25 | 3719 | 1.87 (1.12, 3.10) | 1.43 (1.11, 1.86) | 0.37 |
| | Damp or mould | 11623 | 1.50 (1.26, 1.79) | 1.66 (1.33, 2.08) | 0.49 | 11712 | 0.86 (0.68, 1.09) | 0.96 (0.79, 1.16) | 0.49 | 3889 | 1.77 (1.25, 2.49) | 1.33 (1.03, 1.72) | 0.19 |
| | Damp in 1st year | 13363 | 1.81 (1.47, 2.25) | 1.35 (1.15, 1.60) | 0.03 | 14423 | 0.83 (0.66, 1.03) | 0.98 (0.85, 1.14) | 0.20 | 4400 | 1.89 (1.30, 2.75) | 1.59 (1.31, 1.93) | 0.43 |
| | Mould in 1st year | 11274 | 1.61 (1.27, 2.04) | 1.42 (1.17, 1.73) | 0.44 | 11514 | 0.75 (0.52, 1.08) | 0.98 (0.81, 1.19) | 0.20 | 3325 | 2.03 (1.16, 3.57) | 1.63 (1.28, 2.08) | 0.48 |
| | Air conditioning | 11116 | 0.92 (0.64, 1.33) | 0.92 (0.73, 1.15) | 0.99 | 12060 | 1.34 (1.08, 1.66) | 1.01 (0.86, 1.17) | 0.03 | 3794 | 0.77 (0.52, 1.15) | 0.90 (0.70, 1.15) | 0.52 |

X: non-atopic, non-asthmatic; NAA: non-atopic asthma; Y:atopic, non-asthmatic; AA: atopic asthma ; LMIC: low-middle-income country; HIC: high-income country; OR=Odds Ratio; CI=Confidence Interval; X: non-atopic, non-asthmatic; NAA: non-atopic asthma; Y:atopic, non-asthmatic; AA: atopic asthma; [a]the OR for pathway 2 multiplied by the OR pathway 3;

**Table S7 continued**

| Category | Risk factor | Pathway 4 X to AA | | | | | | Pathway 5 NAA to AA | | | |
|---|---|---|---|---|---|---|---|---|---|---|---|
| | | *n* | LMIC OR (95% CI) | HIC OR (95% CI) | Inter-action *P*-value | LMIC predicted OR[a] | HIC predicted OR[a] | *n* | LMIC OR (95% CI) | HIC OR (95% CI) | Inter-action *P*-value |
| House/ bedding | Feather bedding | 10371 | 0.49 (0.29, 0.83) | 0.51 (0.42, 0.61) | 0.88 | 0.70 | 0.55 | 3138 | 0.66 (0.36, 1.21) | 0.77 (0.59, 1.00) | 0.66 |
| | Synthetic bedding | 10370 | 1.79 (1.26, 2.55) | 1.88 (1.61, 2.21) | 0.80 | 1.79 | 1.77 | 3138 | 1.50 (1.09, 2.07) | 1.30 (1.05, 1.61) | 0.46 |
| | Blankets | 11519 | 0.87 (0.67, 1.15) | 0.78 (0.66, 0.93) | 0.51 | 1.10 | 0.78 | 3300 | 0.91 (0.70, 1.18) | 0.63 (0.50, 0.80) | 0.05 |
| | Feather pillow | 10946 | 1.12 (0.70, 1.77) | 0.43 (0.34, 0.54) | 0.00 | 1.13 | 0.49 | 3162 | 1.15 (0.77, 1.71) | 0.57 (0.43, 0.76) | 0.01 |
| | Synthetic pillow | 10286 | 1.74 (1.12, 2.72) | 1.40 (1.20, 1.63) | 0.36 | 1.25 | 1.34 | 3110 | 1.66 (1.14, 2.42) | 1.40 (1.14, 1.71) | 0.44 |
| | Double glazing | 15498 | 0.90 (0.62, 1.30) | 0.94 (0.81, 1.09) | 0.84 | 1.05 | 0.91 | 3429 | 1.25 (0.89, 1.75) | 1.12 (0.91, 1.36) | 0.57 |
| | Double glazing in 1st year | 15068 | 0.76 (0.50, 1.15) | 0.97 (0.84, 1.11) | 0.28 | 0.88 | 0.96 | 3320 | 1.31 (0.90, 1.90) | 0.92 (0.77, 1.11) | 0.10 |
| | Urban | 17609 | 1.09 (0.78, 1.52) | 0.90 (0.73, 1.11) | 0.34 | 1.16 | 0.90 | 4264 | 1.24 (0.88, 1.75) | 0.95 (0.72, 1.24) | 0.22 |
| | Urban in 1st year | 12730 | 1.14 (0.69, 1.87) | 0.99 (0.79, 1.25) | 0.63 | 1.08 | 1.04 | 2980 | 1.43 (0.91, 2.24) | 1.00 (0.74, 1.36) | 0.21 |
| Heating/ cooking | Heating | 12761 | 0.94 (0.62, 1.42) | 1.00 (0.77, 1.29) | 0.82 | 0.94 | 1.04 | 2619 | 1.03 (0.65, 1.62) | 0.92 (0.62, 1.35) | 0.71 |
| | Wood heating | 13538 | 1.23 (0.80, 1.87) | 0.82 (0.66, 1.03) | 0.10 | 1.30 | 0.87 | 3447 | 1.00 (0.71, 1.41) | 1.04 (0.76, 1.41) | 0.87 |
| | Coal/coke heating | 12948 | 1.47 (0.77, 2.82) | 2.14 (1.32, 3.47) | 0.37 | 1.32 | 2.37 | 3350 | 0.70 (0.45, 1.08) | 3.02 (1.31, 6.93) | 0.00 |
| | Gas heating | 12030 | 0.95 (0.60, 1.51) | 1.00 (0.83, 1.20) | 0.85 | 0.97 | 0.96 | 2822 | 1.31 (0.89, 1.91) | 1.03 (0.81, 1.33) | 0.31 |
| | Heating in 1st year | 11747 | 0.76 (0.50, 1.17) | 1.10 (0.85, 1.43) | 0.16 | 0.78 | 1.16 | 2031 | 0.66 (0.39, 1.10) | 1.14 (0.77, 1.69) | 0.09 |
| | Wood heating in 1st year | 12546 | 1.06 (0.69, 1.64) | 0.98 (0.77, 1.26) | 0.76 | 1.22 | 1.04 | 2631 | 0.77 (0.54, 1.11) | 1.44 (1.01, 2.04) | 0.02 |
| | Coal/coke heating in 1st year | 12117 | 1.45 (0.82, 2.57) | 1.32 (0.82, 2.13) | 0.80 | 1.51 | 1.20 | 2564 | 0.73 (0.45, 1.18) | 1.50 (0.73, 3.09) | 0.10 |
| | Cooking electric | 15372 | 0.92 (0.64, 1.33) | 1.00 (0.83, 1.20) | 0.71 | 0.83 | 0.99 | 3568 | 0.91 (0.63, 1.31) | 0.94 (0.73, 1.22) | 0.86 |
| | Cooking gas | 22129 | 1.85 (1.27, 2.70) | 0.95 (0.83, 1.09) | 0.00 | 1.88 | 0.94 | 5339 | 1.90 (1.23, 2.94) | 0.96 (0.80, 1.15) | 0.00 |
| | Cooking gas in 1st year | 13801 | 1.05 (0.62, 1.78) | 0.91 (0.78, 1.07) | 0.62 | 0.96 | 0.90 | 3553 | 1.40 (0.86, 2.28) | 0.96 (0.79, 1.18) | 0.17 |
| Animals | Pets | 15574 | 1.15 (0.91, 1.46) | 0.77 (0.67, 0.88) | 0.00 | 1.24 | 0.82 | 4087 | 1.06 (0.84, 1.34) | 0.78 (0.65, 0.93) | 0.04 |
| | Cats | 22939 | 1.12 (0.87, 1.44) | 0.73 (0.63, 0.84) | 0.00 | 1.22 | 0.73 | 5321 | 1.35 (1.02, 1.77) | 0.67 (0.55, 0.81) | 0.00 |
| | Dogs | 22926 | 1.19 (0.93, 1.53) | 0.83 (0.72, 0.96) | 0.01 | 1.33 | 0.87 | 5321 | 1.03 (0.81, 1.31) | 0.70 (0.58, 0.85) | 0.02 |
| | Birds | 18266 | 1.32 (0.77, 2.27) | 0.87 (0.72, 1.04) | 0.15 | 1.27 | 0.86 | 3734 | 0.97 (0.61, 1.54) | 0.90 (0.70, 1.16) | 0.81 |
| | Cat in 1st year | 15618 | 1.56 (1.03, 2.37) | 0.83 (0.70, 0.99) | 0.01 | 1.58 | 0.80 | 3644 | 0.97 (0.58, 1.61) | 0.90 (0.72, 1.12) | 0.79 |
| | Dog in 1st year | 15618 | 2.55 (1.70, 3.82) | 0.80 (0.67, 0.95) | 0.00 | 2.99 | 0.84 | 3644 | 1.27 (0.82, 1.97) | 0.82 (0.65, 1.03) | 0.08 |
| | Bird in 1st year | 15215 | 2.37 (1.35, 4.16) | 1.06 (0.85, 1.32) | 0.01 | 3.51 | 1.19 | 3365 | 1.29 (0.74, 2.27) | 0.76 (0.57, 1.01) | 0.10 |
| | Farm animals | 16580 | 1.07 (0.79, 1.44) | 0.69 (0.51, 0.94) | 0.05 | 1.22 | 0.77 | 3785 | 1.12 (0.85, 1.47) | 0.51 (0.35, 0.76) | 0.00 |
| Diet/ weight | Meat weekly | 19588 | 1.10 (0.82, 1.47) | 0.84 (0.63, 1.12) | 0.20 | 0.99 | 0.79 | 4281 | 0.94 (0.71, 1.25) | 1.08 (0.73, 1.59) | 0.59 |
| | Seafood weekly | 19941 | 0.83 (0.64, 1.08) | 1.00 (0.87, 1.15) | 0.23 | 0.77 | 0.96 | 4434 | 0.80 (0.60, 1.07) | 1.09 (0.90, 1.33) | 0.08 |
| | Fruit weekly | 18730 | 0.97 (0.73, 1.29) | 0.88 (0.70, 1.11) | 0.60 | 0.95 | 0.84 | 4804 | 1.06 (0.79, 1.42) | 0.97 (0.71, 1.32) | 0.67 |
| | Green vegetables weekly | 18441 | 1.07 (0.83, 1.38) | 1.04 (0.88, 1.22) | 0.85 | 1.04 | 1.07 | 4336 | 0.95 (0.73, 1.23) | 1.07 (0.86, 1.33) | 0.49 |
| | Fast food weekly | 13385 | 0.92 (0.66, 1.28) | 1.13 (0.94, 1.35) | 0.29 | 0.76 | 1.13 | 3617 | 0.80 (0.58, 1.10) | 0.93 (0.73, 1.19) | 0.47 |
| | Fruit juice weekly | 13577 | 0.86 (0.67, 1.09) | 0.94 (0.82, 1.09) | 0.50 | 0.68 | 0.95 | 3613 | 0.77 (0.58, 1.02) | 0.81 (0.66, 0.99) | 0.76 |
| | Overweight | 10635 | 1.11 (0.81, 1.51) | 1.29 (1.08, 1.53) | 0.41 | 1.39 | 1.32 | 3906 | 1.29 (0.97, 1.72) | 0.75 (0.61, 0.93) | 0.00 |
| | Obese | 10635 | 1.52 (0.92, 2.49) | 1.30 (0.96, 1.74) | 0.59 | 1.57 | 1.39 | 3906 | 1.76 (1.10, 2.83) | 0.64 (0.47, 0.88) | 0.00 |
| Family/ birth | Older siblings | 22216 | 0.79 (0.63, 0.99) | 0.92 (0.82, 1.03) | 0.23 | 0.98 | 0.96 | 4879 | 0.76 (0.61, 0.96) | 0.92 (0.79, 1.08) | 0.18 |
| | Younger siblings | 22182 | 0.78 (0.62, 0.98) | 0.99 (0.88, 1.11) | 0.07 | 0.79 | 0.95 | 4868 | 0.71 (0.56, 0.89) | 1.02 (0.87, 1.19) | 0.01 |
| | More than 1 sibling | 22216 | 0.71 (0.55, 0.91) | 0.94 (0.84, 1.07) | 0.04 | 0.92 | 0.98 | 4882 | 0.64 (0.50, 0.82) | 0.95 (0.81, 1.12) | 0.01 |
| | Ever breastfed | 22239 | 0.95 (0.69, 1.31) | 0.91 (0.80, 1.05) | 0.81 | 0.86 | 0.88 | 4956 | 0.95 (0.65, 1.40) | 1.22 (1.02, 1.47) | 0.25 |
| | Low birthweight | 15081 | 0.75 (0.46, 1.22) | 1.11 (0.89, 1.39) | 0.14 | 0.70 | 1.13 | 3648 | 0.73 (0.45, 1.20) | 0.82 (0.61, 1.11) | 0.69 |
| | Premature | 12599 | 1.00 (0.68, 1.47) | 1.06 (0.84, 1.35) | 0.80 | 1.17 | 1.05 | 3237 | 0.80 (0.51, 1.24) | 0.68 (0.50, 0.91) | 0.55 |
| | Parental allergic diseases | 23503 | 2.93 (2.36, 3.64) | 2.78 (2.46, 3.13) | 0.67 | 2.82 | 2.84 | 5068 | 0.90 (0.72, 1.12) | 1.57 (1.33, 1.86) | 0.00 |

| | | | | | | | | | | | |
|---|---|---|---|---|---|---|---|---|---|---|---|
| | Maternal education | 13907 | 1.31 (0.98, 1.74) | 1.13 (0.86, 1.49) | 0.47 | 1.06 | 1.17 | 3113 | 1.40 (1.03, 1.91) | 1.39 (0.98, 1.99) | 0.98 |
| | Paternal education | 14061 | 0.84 (0.59, 1.19) | 0.89 (0.69, 1.15) | 0.76 | 0.70 | 0.92 | 3146 | 0.94 (0.65, 1.36) | 1.01 (0.72, 1.43) | 0.76 |
| Air-quality | Any smokers | 22118 | 0.91 (0.74, 1.12) | 0.89 (0.79, 1.01) | 0.87 | 0.98 | 0.86 | 4871 | 0.89 (0.71, 1.12) | 0.67 (0.57, 0.79) | 0.05 |
| | Maternal smoking | 23302 | 1.15 (0.85, 1.54) | 1.03 (0.91, 1.16) | 0.50 | 1.26 | 1.00 | 5046 | 1.03 (0.78, 1.36) | 0.68 (0.57, 0.80) | 0.01 |
| | Maternal smoking in 1st year | 20641 | 1.26 (0.91, 1.74) | 1.14 (0.99, 1.31) | 0.58 | 1.50 | 1.14 | 4588 | 0.83 (0.61, 1.12) | 0.71 (0.59, 0.86) | 0.42 |
| | Maternal smoking during pregnancy | 21483 | 1.35 (0.96, 1.90) | 1.20 (1.03, 1.39) | 0.53 | 1.47 | 1.20 | 4779 | 0.99 (0.72, 1.37) | 0.77 (0.64, 0.94) | 0.19 |
| | Damp | 12896 | 1.33 (1.01, 1.76) | 1.22 (0.98, 1.52) | 0.64 | 1.55 | 1.22 | 3746 | 1.01 (0.77, 1.33) | 0.83 (0.63, 1.10) | 0.32 |
| | Mould | 11037 | 1.30 (0.94, 1.81) | 1.26 (1.03, 1.54) | 0.87 | 1.31 | 1.28 | 3675 | 1.03 (0.77, 1.38) | 0.81 (0.63, 1.04) | 0.22 |
| | Damp or mould | 11275 | 1.37 (1.06, 1.76) | 1.27 (1.02, 1.57) | 0.65 | 1.52 | 1.27 | 3800 | 1.11 (0.88, 1.41) | 0.86 (0.66, 1.12) | 0.16 |
| | Damp in 1st year | 13065 | 1.74 (1.29, 2.35) | 1.51 (1.29, 1.76) | 0.40 | 1.56 | 1.56 | 3340 | 0.94 (0.69, 1.30) | 1.11 (0.91, 1.36) | 0.40 |
| | Mould in 1st year | 10839 | 1.67 (1.11, 2.49) | 1.55 (1.29, 1.87) | 0.76 | 1.52 | 1.60 | 3085 | 1.06 (0.74, 1.52) | 1.06 (0.84, 1.35) | 1.00 |
| | Air conditioning | 10888 | 1.20 (0.85, 1.69) | 0.95 (0.76, 1.18) | 0.26 | 1.03 | 0.90 | 2850 | 1.19 (0.72, 1.96) | 0.97 (0.72, 1.32) | 0.51 |

**Table S8: Results from mixed effects logistic regression models for ISAAC study only, with random intercept for centre, adjusted for age, sex, and country income group**

| Category (for figures) | Risk factor | Pathway 1 X to NAA | | Pathway 2 X to Y | | Pathway 3 Y to AA | | Pathway 4 X to AA | | |
|---|---|---|---|---|---|---|---|---|---|---|
| | | *n* | OR (95% CI) | *n* | OR (95% CI) | *n* | OR (95% CI) | *n* | OR (95% CI) | predicted OR[a] |
| House/ bedding | Feather bedding | 10266 | 0.61 (0.50, 0.75) | 10828 | 0.88 (0.77, 1.00) | 3214 | 0.61 (0.49, 0.76) | 9926 | 0.50 (0.41, 0.60) | 0.54 |
| | Synthetic bedding | 10266 | 1.35 (1.17, 1.56) | 10828 | 1.16 (1.04, 1.30) | 3214 | 1.59 (1.34, 1.90) | 9926 | 1.93 (1.66, 2.25) | 1.85 |
| | Blankets | 11130 | 1.11 (0.96, 1.29) | 11657 | 0.89 (0.80, 1.00) | 3062 | 0.95 (0.79, 1.14) | 10671 | 0.82 (0.70, 0.96) | 0.84 |
| | Feather pillow | 10905 | 0.74 (0.61, 0.91) | 11428 | 0.90 (0.78, 1.03) | 3199 | 0.57 (0.44, 0.74) | 10501 | 0.44 (0.35, 0.55) | 0.51 |
| | Synthetic pillow | 10199 | 1.06 (0.91, 1.23) | 10760 | 1.00 (0.89, 1.11) | 3185 | 1.35 (1.14, 1.61) | 9841 | 1.46 (1.26, 1.69) | 1.35 |
| | Double glazing | 13765 | 0.89 (0.77, 1.03) | 15477 | 0.93 (0.84, 1.03) | 4342 | 1.00 (0.84, 1.19) | 13477 | 0.92 (0.78, 1.08) | 0.93 |
| | Double glazing in 1st year | 13345 | 0.98 (0.84, 1.14) | 14945 | 0.95 (0.86, 1.05) | 4134 | 0.97 (0.81, 1.17) | 13059 | 0.87 (0.74, 1.02) | 0.92 |
| | Urban | 11926 | 0.80 (0.64, 0.99) | 12831 | 1.27 (1.05, 1.54) | 3433 | 0.71 (0.53, 0.95) | 11510 | 0.90 (0.70, 1.14) | 0.90 |
| | Urban in 1st year | 11262 | 0.86 (0.70, 1.05) | 12173 | 1.11 (0.95, 1.30) | 3144 | 1.02 (0.77, 1.35) | 10853 | 1.09 (0.86, 1.38) | 1.14 |
| Heating/ cooking | Heating | 12471 | 1.03 (0.83, 1.26) | 14170 | 0.87 (0.75, 1.01) | 3831 | 1.20 (0.94, 1.53) | 12315 | 0.99 (0.79, 1.23) | 1.04 |
| | Wood heating | 13242 | 1.00 (0.83, 1.21) | 14702 | 0.92 (0.78, 1.08) | 4221 | 1.06 (0.82, 1.37) | 12945 | 0.95 (0.77, 1.18) | 0.97 |
| | Coal/coke heating | 12675 | 1.09 (0.73, 1.61) | 14016 | 1.01 (0.77, 1.30) | 4005 | 1.72 (1.08, 2.74) | 12355 | 1.82 (1.23, 2.71) | 1.73 |
| | Gas heating | 12041 | 0.96 (0.80, 1.15) | 13325 | 1.05 (0.93, 1.18) | 3897 | 0.89 (0.73, 1.10) | 11792 | 0.96 (0.80, 1.14) | 0.94 |
| | Heating in 1st year | 11900 | 1.01 (0.82, 1.25) | 13492 | 0.86 (0.74, 1.00) | 3623 | 1.20 (0.94, 1.53) | 11747 | 0.99 (0.80, 1.24) | 1.04 |
| | Wood heating in 1st year | 12799 | 0.97 (0.80, 1.18) | 14263 | 0.88 (0.75, 1.03) | 4095 | 1.24 (0.95, 1.63) | 12546 | 1.00 (0.81, 1.24) | 1.09 |
| | Coal/coke heating in 1st year | 12385 | 1.16 (0.81, 1.66) | 13766 | 0.92 (0.73, 1.15) | 3945 | 1.39 (0.90, 2.13) | 12117 | 1.37 (0.95, 1.98) | 1.27 |
| | Cooking electric | 15082 | 1.07 (0.90, 1.28) | 16674 | 1.02 (0.92, 1.14) | 4608 | 0.94 (0.78, 1.12) | 14766 | 0.98 (0.83, 1.15) | 0.96 |
| | Cooking gas | 15868 | 0.96 (0.82, 1.13) | 17672 | 0.99 (0.89, 1.10) | 4950 | 1.14 (0.95, 1.37) | 15596 | 1.10 (0.93, 1.29) | 1.13 |
| | Cooking gas in 1st year | 11579 | 1.00 (0.82, 1.22) | 12869 | 0.89 (0.77, 1.02) | 3768 | 1.11 (0.87, 1.40) | 11373 | 0.99 (0.81, 1.21) | 0.98 |
| Animals | Pets | 9118 | 1.02 (0.88, 1.19) | 9697 | 0.89 (0.79, 1.01) | 2290 | 1.12 (0.93, 1.36) | 8923 | 0.95 (0.80, 1.12) | 1.00 |
| | Cats | 16426 | 1.07 (0.91, 1.26) | 18523 | 0.82 (0.73, 0.92) | 5049 | 1.04 (0.85, 1.26) | 16296 | 0.78 (0.66, 0.92) | 0.85 |
| | Dogs | 16424 | 1.14 (0.97, 1.34) | 18522 | 0.91 (0.81, 1.03) | 5049 | 1.01 (0.83, 1.22) | 16297 | 0.93 (0.78, 1.10) | 0.92 |
| | Birds | 16428 | 0.98 (0.83, 1.17) | 18525 | 0.84 (0.74, 0.95) | 5050 | 1.11 (0.90, 1.36) | 16299 | 0.93 (0.77, 1.12) | 0.93 |
| | Cat in 1st year | 13323 | 0.93 (0.75, 1.16) | 15102 | 0.81 (0.69, 0.94) | 4349 | 1.13 (0.88, 1.45) | 13199 | 0.91 (0.74, 1.12) | 0.91 |
| | Dog in 1st year | 13323 | 0.97 (0.78, 1.20) | 15102 | 0.76 (0.64, 0.89) | 4349 | 1.32 (1.03, 1.69) | 13199 | 0.94 (0.76, 1.15) | 1.00 |
| | Bird in 1st year | 13323 | 1.39 (1.12, 1.74) | 15102 | 0.95 (0.80, 1.12) | 4349 | 1.55 (1.21, 2.00) | 13199 | 1.34 (1.08, 1.67) | 1.47 |
| | Farm animals | 13067 | 1.15 (0.94, 1.42) | 14153 | 0.73 (0.59, 0.91) | 3877 | 1.23 (0.89, 1.69) | 12662 | 0.83 (0.64, 1.08) | 0.90 |
| Diet/ weight | Meat weekly | 15494 | 0.98 (0.80, 1.20) | 17152 | 0.99 (0.84, 1.15) | 4658 | 0.88 (0.66, 1.17) | 15178 | 0.96 (0.75, 1.24) | 0.87 |
| | Seafood weekly | 15822 | 0.99 (0.87, 1.12) | 17482 | 1.04 (0.95, 1.13) | 4812 | 0.87 (0.75, 1.02) | 15532 | 0.95 (0.83, 1.09) | 0.90 |
| | Fruit weekly | 12560 | 0.74 (0.61, 0.88) | 13667 | 1.03 (0.88, 1.21) | 3740 | 0.80 (0.62, 1.04) | 12211 | 0.84 (0.67, 1.04) | 0.83 |
| | Green vegetables weekly | 12706 | 0.96 (0.82, 1.13) | 13840 | 1.02 (0.90, 1.16) | 3781 | 1.05 (0.85, 1.30) | 12359 | 1.03 (0.85, 1.23) | 1.07 |
| | Fast food weekly | 9275 | 1.23 (1.06, 1.44) | 9667 | 1.10 (0.97, 1.25) | 2626 | 0.96 (0.78, 1.17) | 8893 | 1.10 (0.93, 1.31) | 1.05 |
| | Fruit juice weekly | 11417 | 1.20 (1.03, 1.41) | 12576 | 1.08 (0.97, 1.20) | 3703 | 0.80 (0.67, 0.96) | 11149 | 0.88 (0.76, 1.03) | 0.86 |
| | Overweight | 4120 | 1.31 (1.07, 1.61) | 4193 | 1.02 (0.85, 1.22) | 1574 | 1.29 (1.00, 1.66) | 3999 | 1.25 (1.02, 1.54) | 1.31 |
| | Obese | 4120 | 1.69 (1.24, 2.30) | 4193 | 0.93 (0.66, 1.30) | 1574 | 1.80 (1.18, 2.72) | 3999 | 1.43 (1.04, 1.97) | 1.67 |
| Family/ birth | Older siblings | 16422 | 1.02 (0.91, 1.15) | 18269 | 0.93 (0.86, 1.01) | 5030 | 1.00 (0.87, 1.14) | 16109 | 0.85 (0.75, 0.96) | 0.93 |
| | Younger siblings | 16391 | 1.14 (1.01, 1.28) | 18214 | 0.95 (0.88, 1.03) | 4995 | 1.02 (0.89, 1.17) | 16075 | 0.97 (0.86, 1.10) | 0.97 |
| | More than 1 sibling | 16425 | 1.14 (1.01, 1.29) | 18258 | 0.90 (0.83, 0.98) | 5019 | 1.12 (0.97, 1.30) | 16109 | 0.92 (0.81, 1.05) | 1.01 |
| | Ever breastfed | 16456 | 0.82 (0.71, 0.95) | 18291 | 0.94 (0.85, 1.03) | 5039 | 0.92 (0.78, 1.08) | 16146 | 0.90 (0.78, 1.03) | 0.86 |

| | | | | | | | | | | |
|---|---|---|---|---|---|---|---|---|---|---|
| | Low birthweight | 13346 | 1.16 (0.96, 1.40) | 14584 | 0.94 (0.82, 1.08) | 4034 | 1.03 (0.81, 1.32) | 13028 | 0.96 (0.77, 1.20) | 0.97 |
| | Premature | 10423 | 1.37 (1.10, 1.71) | 11254 | 0.86 (0.73, 1.02) | 2926 | 1.29 (0.97, 1.72) | 10118 | 1.08 (0.84, 1.39) | 1.11 |
| | Parental allergic diseases | 17983 | 2.17 (1.94, 2.44) | 20012 | 1.41 (1.30, 1.53) | 5479 | 2.01 (1.75, 2.31) | 17679 | 2.78 (2.46, 3.13) | 2.84 |
| | Maternal education | 8546 | 0.76 (0.61, 0.93) | 9156 | 0.97 (0.83, 1.14) | 2067 | 1.15 (0.87, 1.52) | 8219 | 1.17 (0.91, 1.51) | 1.12 |
| | Paternal education | 8570 | 0.89 (0.73, 1.09) | 9179 | 1.10 (0.94, 1.29) | 2070 | 0.78 (0.60, 1.02) | 8241 | 0.89 (0.71, 1.13) | 0.86 |
| Air-quality | Any smokers | 16393 | 1.28 (1.14, 1.44) | 18222 | 0.84 (0.78, 0.91) | 5018 | 1.03 (0.90, 1.17) | 16092 | 0.85 (0.76, 0.96) | 0.87 |
| | Maternal smoking | 17579 | 1.51 (1.33, 1.70) | 19555 | 0.82 (0.75, 0.90) | 5333 | 1.24 (1.07, 1.44) | 17292 | 1.02 (0.90, 1.16) | 1.02 |
| | Maternal smoking in 1st year | 14968 | 1.65 (1.44, 1.90) | 16513 | 0.86 (0.77, 0.96) | 4448 | 1.35 (1.13, 1.61) | 14643 | 1.16 (1.00, 1.34) | 1.16 |
| | Maternal smoking during pregnancy | 15755 | 1.59 (1.37, 1.85) | 17487 | 0.88 (0.77, 1.00) | 4776 | 1.33 (1.09, 1.61) | 15455 | 1.16 (0.99, 1.37) | 1.16 |
| | Damp | 11657 | 1.91 (1.63, 2.25) | 12817 | 0.87 (0.75, 1.01) | 3791 | 1.75 (1.38, 2.20) | 11346 | 1.62 (1.34, 1.95) | 1.52 |
| | Mould | 9908 | 1.64 (1.36, 1.97) | 10210 | 0.86 (0.69, 1.08) | 2873 | 1.76 (1.29, 2.41) | 9467 | 1.63 (1.28, 2.08) | 1.52 |
| | Damp or mould | 9785 | 1.82 (1.55, 2.14) | 10201 | 0.91 (0.77, 1.08) | 2849 | 1.79 (1.39, 2.31) | 9346 | 1.71 (1.40, 2.09) | 1.63 |
| | Damp in 1st year | 11366 | 1.76 (1.51, 2.06) | 12498 | 0.88 (0.76, 1.02) | 3684 | 2.04 (1.65, 2.52) | 11056 | 1.88 (1.58, 2.23) | 1.80 |
| | Mould in 1st year | 9280 | 1.62 (1.34, 1.95) | 9593 | 0.90 (0.72, 1.11) | 2612 | 1.92 (1.44, 2.58) | 8835 | 1.79 (1.42, 2.25) | 1.73 |
| | Air conditioning | 10545 | 0.91 (0.75, 1.10) | 11557 | 1.08 (0.96, 1.23) | 3298 | 0.89 (0.72, 1.10) | 10193 | 1.00 (0.83, 1.20) | 0.96 |

ISAAC=International Study of Asthma and Allergies in Childhood; OR=Odds Ratio; CI=Confidence Interval; X: non-atopic, non-asthmatic; NAA: non-atopic asthma; Y:atopic, non-asthmatic; AA: atopic asthma; [a]the OR for pathway 2 multiplied by the OR pathway 3;

**Table S9: Results from mixed effects logistic regression models for WASP study only, with random intercept for centre, adjusted for age, sex, and country income group**

| Category | Risk factor | Pathway 1 X to NAA | | Pathway 2 X to Y | | Pathway 3 Y to AA | | Pathway 4 X to AA | | |
|---|---|---|---|---|---|---|---|---|---|---|
| | | *n* | OR (95% CI) | *n* | OR (95% CI) | *n* | OR (95% CI) | *n* | OR (95% CI) | predicted OR[a] |
| House/ bedding | Feather bedding | 301 | 0.73 (0.42, 1.26) | 190 | 0.67 (0.33, 1.34) | 375 | 0.93 (0.49, 1.79) | 445 | 0.57 (0.35, 0.93) | 0.62 |
| | Synthetic bedding | 300 | 1.05 (0.55, 2.02) | 189 | 1.93 (0.93, 4.02) | 375 | 0.90 (0.50, 1.62) | 444 | 1.27 (0.76, 2.14) | 1.74 |
| | Blankets | 301 | 0.62 (0.38, 1.02) | 190 | 0.67 (0.35, 1.29) | 375 | 0.58 (0.33, 1.03) | 445 | 0.44 (0.28, 0.68) | 0.39 |
| | Feather pillow | 301 | 1.11 (0.68, 1.84) | 190 | 0.52 (0.25, 1.09) | 375 | 1.01 (0.51, 2.00) | 445 | 1.05 (0.64, 1.74) | 0.52 |
| | Synthetic pillow | 301 | 0.92 (0.45, 1.85) | 190 | 1.14 (0.52, 2.53) | 375 | 1.46 (0.79, 2.67) | 445 | 1.11 (0.60, 2.06) | 1.67 |
| Heating/ cooking | Heating | 301 | 0.25 (0.14, 0.44) | 190 | 2.39 (1.13, 5.08) | 376 | N/A | 446 | N/A | N/A |
| | Wood heating | 435 | 0.36 (0.16, 0.83) | 237 | 0.52 (0.24, 1.14) | 488 | 0.87 (0.45, 1.68) | 593 | 0.55 (0.32, 0.97) | 0.46 |
| | Coal/coke heating | 435 | N/A | 237 | N/A | 488 | N/A | 593 | N/A | N/A |
| | Gas heating | 111 | 0.62 (0.25, 1.54) | 114 | 1.44 (0.64, 3.26) | 212 | 1.10 (0.55, 2.22) | 238 | 1.46 (0.80, 2.67) | 1.59 |
| | Cooking electric | 188 | 1.91 (0.71, 5.16) | 73 | 4.32 (0.83, 22.42) | 158 | 0.21 (0.04, 1.04) | 203 | 1.19 (0.37, 3.87) | 0.90 |
| | Cooking gas | 300 | 0.42 (0.20, 0.87) | 189 | 1.32 (0.60, 2.92) | 376 | 0.68 (0.34, 1.34) | 445 | 0.83 (0.47, 1.44) | 0.90 |
| Animals | Pets | 432 | 0.72 (0.46, 1.12) | 234 | 0.85 (0.42, 1.72) | 483 | 0.64 (0.35, 1.16) | 587 | 0.56 (0.36, 0.85) | 0.54 |
| | Cats | 432 | 0.79 (0.50, 1.24) | 234 | 1.05 (0.56, 1.95) | 483 | 0.52 (0.30, 0.90) | 587 | 0.65 (0.43, 0.99) | 0.54 |
| | Dogs | 432 | 0.86 (0.56, 1.32) | 234 | 0.71 (0.37, 1.34) | 483 | 1.26 (0.71, 2.24) | 587 | 0.82 (0.56, 1.21) | 0.89 |
| | Farm animals | 304 | 0.72 (0.42, 1.25) | 111 | 5.54 (0.66, 46.59) | 179 | 0.11 (0.01, 0.85) | 262 | 0.58 (0.33, 1.00) | 0.59 |
| Diet/ weight | Meat weekly | 277 | 1.01 (0.22, 4.62) | 177 | 0.78 (0.19, 3.15) | 280 | 1.15 (0.35, 3.76) | 351 | 0.74 (0.25, 2.16) | 0.90 |
| | Seafood weekly | 277 | 1.42 (0.74, 2.72) | 176 | 1.45 (0.69, 3.04) | 280 | 1.09 (0.57, 2.07) | 350 | 1.41 (0.84, 2.35) | 1.58 |
| | Fruit weekly | 296 | 0.52 (0.13, 2.03) | 186 | 0.28 (0.06, 1.46) | 371 | 1.10 (0.34, 3.58) | 439 | 0.76 (0.19, 3.04) | 0.31 |
| | Fastfood weekly | 297 | 1.13 (0.64, 2.00) | 187 | 1.07 (0.54, 2.12) | 372 | 1.41 (0.78, 2.52) | 441 | 0.94 (0.56, 1.59) | 1.50 |
| | Overweight | 435 | 1.32 (0.82, 2.14) | 237 | 1.02 (0.48, 2.18) | 486 | 1.45 (0.74, 2.84) | 591 | 1.19 (0.76, 1.87) | 1.49 |
| | Obese | 435 | 1.94 (0.70, 5.35) | 237 | 0.58 (0.10, 3.30) | 486 | 2.64 (0.61, 11.54) | 591 | 2.22 (0.88, 5.59) | 1.54 |
| Air-quality | Damp | 435 | 0.57 (0.36, 0.89) | 237 | 0.69 (0.34, 1.41) | 488 | 0.87 (0.45, 1.69) | 593 | 0.58 (0.38, 0.89) | 0.60 |
| | Mould | 422 | 0.89 (0.58, 1.39) | 231 | 0.97 (0.51, 1.81) | 465 | 0.93 (0.53, 1.63) | 572 | 0.87 (0.58, 1.29) | 0.89 |
| | Air conditioning | 128 | N/A | 124 | N/A | 302 | 0.49 (0.12, 2.01) | 324 | N/A | N/A |

WASP=World Asthma Phenotypes; OR=Odds Ratio; CI=Confidence Interval; X: non-atopic, non-asthmatic; NAA: non-atopic asthma; Y:atopic, non-asthmatic; AA: atopic asthma; [a]the OR for pathway 2 multiplied by the OR pathway 3;

**Table S10: Results from mixed effects logistic regression models for SCAALA study only, with random intercept for centre, adjusted for age and sex**

| Category | Risk factor | Pathway 1 X to NAA | | Pathway 2 X to Y | | Pathway 3 Y to AA | | Pathway 4 X to AA | | |
|---|---|---|---|---|---|---|---|---|---|---|
| | | *n* | OR (95% CI) | *n* | OR (95% CI) | *n* | OR (95% CI) | *n* | OR (95% CI) | predicted OR[a] |
| House/ bedding | Blankets | 478 | 1.14 (0.72, 1.79) | 413 | 0.72 (0.40, 1.30) | 214 | 2.28 (1.17, 4.47) | 403 | 1.67 (0.99, 2.81) | 1.64 |
| Heating/ cooking | Cooking electric | 478 | N/A | 413 | N/A | 214 | N/A | 403 | N/A | N/A |
| | Cooking gas | 4555 | 0.51 (0.34, 0.76) | 4416 | 0.49 (0.33, 0.75) | 763 | 2.77 (0.82, 9.41) | 4059 | 1.31 (0.40, 4.27) | 1.37 |
| | Cooking gas in 1st year | 478 | N/A | 413 | N/A | 214 | 0.46 (0.04, 5.16) | 403 | N/A | N/A |
| Animals | Pets | 4559 | 1.21 (1.02, 1.43) | 4420 | 0.85 (0.71, 1.02) | 763 | 1.52 (1.07, 2.17) | 4063 | 1.27 (0.93, 1.73) | 1.30 |
| | Cats | 4558 | 0.93 (0.77, 1.11) | 4419 | 0.70 (0.57, 0.86) | 762 | 1.99 (1.33, 2.99) | 4063 | 1.35 (0.95, 1.90) | 1.39 |
| | Dogs | 4557 | 1.34 (1.13, 1.58) | 4419 | 0.95 (0.79, 1.15) | 763 | 1.42 (0.99, 2.04) | 4062 | 1.29 (0.95, 1.77) | 1.35 |
| | Cat in 1st year | 478 | 2.81 (1.36, 5.78) | 413 | 0.71 (0.23, 2.23) | 214 | 3.59 (1.11, 11.60) | 403 | 2.35 (1.03, 5.35) | 2.56 |
| | Dog in 1st year | 478 | 2.13 (1.32, 3.45) | 413 | 0.57 (0.26, 1.21) | 214 | 4.70 (2.10, 10.51) | 403 | 2.50 (1.44, 4.33) | 2.66 |
| | Farm animals | 4076 | 1.07 (0.85, 1.34) | 4003 | 1.47 (1.18, 1.83) | 549 | 0.85 (0.51, 1.39) | 3656 | 1.25 (0.79, 1.97) | 1.25 |
| Diet/ weight | Meat weekly | 4554 | 1.09 (0.86, 1.38) | 4415 | 0.80 (0.62, 1.01) | 762 | 1.07 (0.71, 1.63) | 4059 | 0.99 (0.68, 1.43) | 0.85 |
| | Seafood weekly | 4554 | 0.91 (0.68, 1.22) | 4415 | 0.99 (0.72, 1.37) | 762 | 0.60 (0.32, 1.11) | 4059 | 0.71 (0.44, 1.17) | 0.59 |
| | Fruit weekly | 4549 | 0.81 (0.63, 1.04) | 4410 | 1.54 (1.12, 2.11) | 762 | 0.70 (0.43, 1.13) | 4054 | 1.16 (0.78, 1.74) | 1.07 |
| | Green vegetables weekly | 4548 | 1.05 (0.87, 1.26) | 4410 | 1.01 (0.82, 1.23) | 761 | 1.01 (0.70, 1.48) | 4055 | 1.13 (0.81, 1.58) | 1.02 |
| | Fast food weekly | 4546 | 0.94 (0.67, 1.31) | 4407 | 1.08 (0.77, 1.51) | 762 | 0.84 (0.45, 1.57) | 4051 | 0.99 (0.57, 1.73) | 0.91 |
| | Fruit juice weekly | 478 | 1.16 (0.79, 1.71) | 413 | 1.36 (0.87, 2.12) | 214 | 0.66 (0.38, 1.14) | 403 | 0.98 (0.61, 1.56) | 0.89 |
| | Overweight | 4461 | 0.89 (0.67, 1.18) | 4322 | 1.24 (0.93, 1.64) | 720 | 1.20 (0.74, 1.94) | 3976 | 1.34 (0.88, 2.03) | 1.48 |
| | Obese | 4461 | 0.81 (0.49, 1.34) | 4322 | 1.37 (0.83, 2.28) | 720 | 1.02 (0.48, 2.17) | 3976 | 1.10 (0.57, 2.11) | 1.39 |
| Family/ birth | Older siblings | 4554 | 0.92 (0.76, 1.11) | 4416 | 1.00 (0.82, 1.24) | 761 | 1.15 (0.78, 1.68) | 4061 | 0.95 (0.68, 1.32) | 1.15 |
| | Younger siblings | 4554 | 1.03 (0.85, 1.24) | 4416 | 0.79 (0.64, 0.97) | 761 | 0.83 (0.58, 1.19) | 4061 | 0.69 (0.50, 0.95) | 0.65 |
| | More than 1 sibling | 4554 | 1.01 (0.81, 1.26) | 4416 | 0.90 (0.70, 1.15) | 761 | 0.86 (0.56, 1.31) | 4061 | 0.66 (0.47, 0.93) | 0.77 |
| | Ever breastfed | 4504 | 0.75 (0.47, 1.20) | 4369 | 0.95 (0.54, 1.67) | 759 | 0.74 (0.30, 1.80) | 4014 | 0.70 (0.33, 1.50) | 0.70 |
| | Premature | 477 | 1.03 (0.50, 2.13) | 412 | 0.86 (0.36, 2.08) | 214 | 1.52 (0.53, 4.32) | 402 | 1.31 (0.57, 2.98) | 1.31 |
| | Parental allergic diseases | 4553 | 3.61 (3.02, 4.31) | 4415 | 1.19 (0.99, 1.44) | 762 | 2.26 (1.59, 3.22) | 4057 | 2.67 (1.96, 3.64) | 2.70 |
| | Maternal education | 4091 | 1.04 (0.86, 1.25) | 3960 | 1.09 (0.89, 1.34) | 695 | 1.01 (0.70, 1.46) | 3637 | 1.28 (0.93, 1.77) | 1.10 |
| | Paternal education | 4240 | 0.97 (0.79, 1.18) | 4104 | 0.97 (0.78, 1.20) | 719 | 0.84 (0.51, 1.37) | 3769 | 0.79 (0.51, 1.24) | 0.81 |
| Air-quality | Any smokers | 4546 | 1.01 (0.85, 1.20) | 4408 | 0.98 (0.82, 1.19) | 762 | 0.84 (0.58, 1.23) | 4052 | 0.82 (0.58, 1.14) | 0.83 |
| | Maternal smoking | 4483 | 1.05 (0.82, 1.35) | 4349 | 0.95 (0.73, 1.25) | 757 | 1.15 (0.70, 1.91) | 3998 | 1.06 (0.68, 1.66) | 1.10 |
| | Maternal smoking in 1st year | 4463 | 1.16 (0.89, 1.52) | 4330 | 1.05 (0.79, 1.40) | 756 | 1.24 (0.74, 2.08) | 3978 | 1.19 (0.75, 1.89) | 1.30 |
| | Maternal smoking during pregnancy | 4454 | 1.20 (0.91, 1.58) | 4320 | 1.12 (0.84, 1.50) | 755 | 1.31 (0.78, 2.22) | 3969 | 1.33 (0.84, 2.11) | 1.47 |
| | Damp or mould | 478 | 1.60 (1.07, 2.37) | 413 | 0.92 (0.59, 1.42) | 214 | 1.54 (0.89, 2.69) | 403 | 1.41 (0.88, 2.26) | 1.41 |
| | Air conditioning | 443 | 1.77 (0.11, 28.71) | 379 | 12.63 (1.37, 116.40) | 194 | N/A | 371 | N/A | N/A |

SCAALA=Social Changes, Asthma and Allergy in Latin America; OR=Odds Ratio; CI=Confidence Interval; X: non-atopic, non-asthmatic; NAA: non-atopic asthma; Y:atopic, non-asthmatic; AA: atopic asthma;
[a]the OR for pathway 2 multiplied by the OR pathway 3;

**Table S11: Results from logistic regression models for ALSPAC study only, adjusted for age and sex**

| Category | Risk factor | Pathway 1 X to NAA | | Pathway 2 X to Y | | Pathway 3 Y to AA | | Pathway 4 X to AA | | |
|---|---|---|---|---|---|---|---|---|---|---|
| | | *n* | OR (95% CI) | *n* | OR (95% CI) | *n* | OR (95% CI) | *n* | OR (95% CI) | predicted OR[a] |
| House/ bedding | Double glazing | 2008 | 0.70 (0.53, 0.91) | 1925 | 1.11 (0.80, 1.54) | 716 | 0.90 (0.61, 1.33) | 2021 | 1.00 (0.75, 1.33) | 1.00 |
| | Double glazing in 1st year | 1999 | 1.03 (0.82, 1.30) | 1929 | 1.18 (0.92, 1.51) | 716 | 0.94 (0.69, 1.27) | 2009 | 1.11 (0.89, 1.40) | 1.11 |
| | Urban | 2034 | 1.05 (0.75, 1.48) | 1937 | 1.24 (0.83, 1.86) | 737 | 0.72 (0.45, 1.15) | 2036 | 0.90 (0.65, 1.25) | 0.90 |
| | Urban in 1st year | 1858 | 0.88 (0.57, 1.37) | 1800 | 1.33 (0.77, 2.29) | 689 | 0.63 (0.34, 1.18) | 1877 | 0.82 (0.53, 1.26) | 0.84 |
| Heating/ cooking | Cooking gas | 2015 | 1.03 (0.82, 1.29) | 1933 | 1.00 (0.78, 1.28) | 722 | 0.98 (0.72, 1.33) | 2029 | 0.96 (0.76, 1.20) | 0.98 |
| | Cooking gas in 1st year | 2017 | 0.80 (0.64, 1.00) | 1941 | 1.06 (0.83, 1.35) | 720 | 0.82 (0.61, 1.10) | 2025 | 0.86 (0.69, 1.07) | 0.86 |
| Animals | Pets | 1992 | 1.11 (0.86, 1.42) | 1904 | 0.81 (0.62, 1.05) | 705 | 0.77 (0.56, 1.05) | 2001 | 0.62 (0.49, 0.78) | 0.62 |
| | Cats | 1974 | 1.08 (0.85, 1.37) | 1900 | 0.94 (0.72, 1.23) | 713 | 0.74 (0.53, 1.04) | 1993 | 0.70 (0.54, 0.90) | 0.70 |
| | Dogs | 1966 | 1.27 (0.97, 1.65) | 1883 | 0.74 (0.54, 1.03) | 705 | 0.89 (0.59, 1.34) | 1980 | 0.67 (0.49, 0.90) | 0.66 |
| | Birds | 1948 | 1.14 (0.73, 1.77) | 1871 | 1.01 (0.61, 1.68) | 704 | 0.71 (0.37, 1.39) | 1967 | 0.74 (0.44, 1.25) | 0.72 |
| | Cat in 1st year | 2007 | 1.02 (0.80, 1.30) | 1932 | 0.91 (0.70, 1.19) | 720 | 0.89 (0.64, 1.24) | 2016 | 0.81 (0.63, 1.04) | 0.81 |
| | Dog in 1st year | 2007 | 1.09 (0.83, 1.43) | 1932 | 0.64 (0.45, 0.90) | 720 | 1.14 (0.75, 1.73) | 2016 | 0.72 (0.53, 0.97) | 0.73 |
| | Bird in 1st year | 2007 | 1.31 (0.84, 2.03) | 1932 | 0.62 (0.33, 1.19) | 720 | 0.72 (0.30, 1.72) | 2016 | 0.45 (0.23, 0.87) | 0.45 |
| Diet/ weight | Green vegetables weekly | 2035 | 1.11 (0.84, 1.45) | 1954 | 0.93 (0.70, 1.23) | 709 | 1.10 (0.77, 1.57) | 2027 | 1.03 (0.79, 1.36) | 1.02 |
| | Fruit weekly | 2031 | 1.61 (0.86, 3.00) | 1952 | 1.77 (0.88, 3.59) | 710 | 0.56 (0.25, 1.25) | 2026 | 0.98 (0.59, 1.64) | 0.99 |
| | Fruit juice weekly | 2031 | 1.03 (0.81, 1.30) | 1951 | 1.00 (0.77, 1.30) | 710 | 0.98 (0.71, 1.35) | 2025 | 1.00 (0.78, 1.27) | 0.98 |
| | Overweight | 2071 | 2.02 (1.52, 2.67) | 1957 | 0.98 (0.67, 1.44) | 748 | 1.25 (0.79, 1.97) | 2069 | 1.24 (0.90, 1.71) | 1.22 |
| | Obese | 2071 | 2.65 (1.60, 4.40) | 1957 | 1.05 (0.48, 2.26) | 748 | 1.10 (0.44, 2.79) | 2069 | 1.14 (0.57, 2.24) | 1.15 |
| Family/ birth | Older siblings | 2051 | 1.13 (0.91, 1.41) | 1972 | 0.82 (0.65, 1.05) | 718 | 1.22 (0.91, 1.65) | 2046 | 1.01 (0.81, 1.26) | 1.01 |
| | Younger siblings | 2051 | 0.74 (0.59, 0.92) | 1972 | 1.32 (1.03, 1.68) | 718 | 0.76 (0.56, 1.02) | 2046 | 1.00 (0.80, 1.25) | 1.00 |
| | More than 1 sibling | 2051 | 0.96 (0.76, 1.21) | 1972 | 0.87 (0.68, 1.13) | 718 | 1.05 (0.77, 1.44) | 2046 | 0.93 (0.74, 1.18) | 0.92 |
| | Ever breastfed | 2079 | 0.60 (0.47, 0.76) | 1972 | 1.11 (0.81, 1.52) | 751 | 0.95 (0.65, 1.40) | 2079 | 1.05 (0.80, 1.39) | 1.05 |
| | Low birthweight | 2051 | 1.98 (1.26, 3.13) | 1944 | 0.87 (0.44, 1.72) | 745 | 1.81 (0.85, 3.86) | 2053 | 1.55 (0.94, 2.56) | 1.56 |
| | Premature | 2084 | 1.60 (1.16, 2.21) | 1972 | 0.76 (0.48, 1.20) | 751 | 1.18 (0.69, 2.04) | 2079 | 0.92 (0.63, 1.36) | 0.90 |
| | Maternal education | 2055 | 1.00 (0.00, 0.00) | 1960 | 1.00 (0.00, 0.00) | 735 | 1.00 (0.00, 0.00) | 2051 | 1.00 (0.00, 0.00) | 1.00 |
| | Paternal education | 2055 | 1.00 (0.00, 0.00) | 1960 | 1.00 (0.00, 0.00) | 735 | 1.00 (0.00, 0.00) | 2051 | 1.00 (0.00, 0.00) | 1.00 |
| Air-quality | Any smokers | 1956 | 1.62 (1.28, 2.06) | 1870 | 0.91 (0.68, 1.22) | 696 | 1.32 (0.94, 1.85) | 1974 | 1.17 (0.92, 1.50) | 1.20 |
| | Maternal smoking | 2002 | 1.99 (1.52, 2.61) | 1919 | 1.06 (0.74, 1.50) | 715 | 1.12 (0.73, 1.70) | 2012 | 1.15 (0.84, 1.56) | 1.18 |
| | Maternal smoking in 1st year | 2010 | 1.73 (1.32, 2.26) | 1933 | 0.70 (0.48, 1.01) | 719 | 1.61 (1.05, 2.49) | 2020 | 1.12 (0.84, 1.51) | 1.13 |
| | Maternal smoking during pregnancy | 2061 | 1.88 (1.45, 2.44) | 1959 | 0.73 (0.50, 1.06) | 744 | 1.85 (1.21, 2.82) | 2059 | 1.36 (1.02, 1.80) | 1.34 |
| | Damp | 936 | 1.39 (0.76, 2.53) | 870 | 0.98 (0.52, 1.83) | 363 | 0.74 (0.37, 1.49) | 957 | 0.69 (0.43, 1.12) | 0.73 |
| | Mould | 972 | 1.52 (1.11, 2.09) | 905 | 0.72 (0.50, 1.03) | 381 | 1.51 (1.00, 2.30) | 998 | 1.08 (0.80, 1.45) | 1.08 |
| | Damp or mould | 934 | 1.21 (0.65, 2.27) | 867 | 0.97 (0.49, 1.90) | 360 | 0.70 (0.33, 1.47) | 953 | 0.64 (0.39, 1.06) | 0.68 |
| | Damp in 1st year | 1997 | 1.11 (0.88, 1.38) | 1925 | 1.08 (0.85, 1.38) | 716 | 1.09 (0.81, 1.46) | 2009 | 1.16 (0.93, 1.44) | 1.18 |
| | Mould in 1st year | 1994 | 1.29 (1.01, 1.66) | 1921 | 0.98 (0.73, 1.30) | 713 | 1.40 (1.00, 1.97) | 2004 | 1.36 (1.06, 1.74) | 1.37 |

ALSPAC= Avon Longitudinal Study of Parents and Children; OR=Odds Ratio; CI=Confidence Interval; X: non-atopic, non-asthmatic; NAA: non-atopic asthma; Y:atopic, non-asthmatic; AA: atopic asthma; [a]the OR for pathway 2 multiplied by the OR pathway 3;

**Table S12: Sensitivity analysis of atopy definition (ISAAC and ALSPAC only)**

| Atopy definition (non-atopy remains as no reaction) | Study | Non-Atopic Non-Asthmatic (X) (%) | Atopic Non-Asthmatic (Y) (%) | Atopic Asthmatic (AA) (%) | Non-Atopic Asthmatic (NAA) (%) | Total | Correlation between pathways 1 and 3 | Correlation between pathways 2 and 3 |
|---|---|---|---|---|---|---|---|---|
| Standard[a] | ISAAC | 16239 (69%) | 3922 (17%) | 1584 (7%) | 1911 (8%) | 23656 | 0.80 (0.67, 0.93) | -0.12 (-0.33, 0.09) |
| | ALSPAC | 1650 (58%) | 322 (11%) | 433 (15%) | 435 (15%) | 2840 | | |
| | TOTAL | 17889 (68%) | 4244 (16%) | 2017 (8%) | 2346 (9%) | 26496 | | |
| Wheal size >=5mm for any core allergen[b] | ISAAC | 16239 (78%) | 1777 (9%) | 979 (5%) | 1911 (9%) | 20906 | 0.63 (0.45, 0.81) | -0.13 (-0.36, 0.10) |
| | ALSPAC | 1650 (72%) | 56 (2%) | 141 (6%) | 435 (19%) | 2282 | | |
| | TOTAL | 17889 (77%) | 1833 (8%) | 1120 (5%) | 2346 (10%) | 23188 | | |
| Wheal size >=4mm for at least 2 core allergens[b] | ISAAC | 16239 (81%) | 1164 (6%) | 745 (4%) | 1911 (10%) | 20059 | 0.63 (0.45, 0.81) | -0.17 (-0.41, 0.07) |
| | ALSPAC | 1650 (76%) | 19 (1%) | 80 (4%) | 435 (20%) | 2184 | | |
| | TOTAL | 17889 (80%) | 1183 (5%) | 825 (4%) | 2346 (11%) | 22243 | | |

ISAAC=International Study of Asthma and Allergies in Childhood; ALSPAC= Avon Longitudinal Study of Parents and Children; [a] wheal size >=3mm for ISAAC or >=2mm for ALSPAC for any allergen; [b] those who meet the standard definition but not the new definition are excluded from the sensitivity analysis.

**Table S13: Sensitivity analyses by centre**

| Sensitivity | Non-Atopic Non-Asthmatic (X) (%) | Atopic Non-Asthmatic (Y) (%) | Atopic Asthmatic (AA) (%) | Non-Atopic Asthmatic (NAA) (%) | Total | Correlation between pathways 1 and 3 | Correlation between pathways 2 and 3 |
|---|---|---|---|---|---|---|---|
| Original | 21920 (67%) | 4870 (15%) | 2642 (8%) | 3309 (10%) | 32741 | 0.81 (0.68, 0.94) | -0.06 (-0.29, 0.17) |
| Omitting WASP | 21749 (68%) | 4804 (15%) | 2220 (7%) | 3045 (10%) | 31818 | 0.82 (0.69, 0.95) | -0.08 (-0.30, 0.14) |
| Omitting ISAAC[a] | 5681 (63%) | 948 (10%) | 1058 (12%) | 1398 (15%) | 9085 | 0.62 (0.36, 0.88) | -0.16 (-0.42, 0.10) |
| Omitting SCAALA | 18060 (66%) | 4310 (16%) | 2439 (9%) | 2610 (10%) | 27419 | 0.79 (0.66, 0.92) | -0.09 (-0.32, 0.14) |
| Omitting ALSPAC | 20270 (68%) | 4548 (15%) | 2209 (7%) | 2874 (10%) | 29901 | 0.80 (0.67, 0.93) | -0.10 (-0.33, 0.13) |
| Controls more lenient on missing data for ISAAC and ALSPAC[b] | 24482 (68%) | 5616 (16%) | 2642 (7%) | 3309 (9%) | 36409 | 0.85 (0.76, 0.94) | -0.15 (-0.35, 0.05) |

ISAAC=International Study of Asthma and Allergies in Childhood; ALSPAC= Avon Longitudinal Study of Parents and Children; WASP=World Asthma Phenotypes; SCAALA=Social Changes, Asthma and Allergy in Latin America; [a] excludes the early life risk factors of any heating, wood heating and coal/coke heating as they were only in the ISAAC study and excludes current coal/coke heating, air conditioning, electric cooking, and any heating due to low sample size; [b] allows for the question "has the child ever been told they have asthma?" to be missing from ISAAC or for one timepoint of data on symptoms to be missing from ALSPAC.
